# Unveiling the Porphyromonadaceae-*TFF1* Interaction and *ITGAM* as Critical Factors in Post-operative Recurrence of Crohn’s Disease

**DOI:** 10.64898/2026.01.16.26344277

**Authors:** Roger Suau, Mireia Lopez-Siles, Margalida Cabrer, Marina Rovira, Laura Clua, Yamile Zabana, Nallely Bueno-Hernández, Robert Benaiges-Fernández, Gisela Piñero, Violeta Lorén, Diandra Monfort-Ferré, Iris Ginés, José Francisco Sánchez-Herrero, Margarita Martínez-Medina, Carolina Serena, Lauro Sumoy, Eugeni Domènech, Míriam Mañosa, Josep Manyé

**Affiliations:** Inflammatory Bowel Diseases Research Group, Germans Trias i Pujol Research Institute (IGTP), Badalona, Spain; Research Group in Microbiology of intestinal disease, Universitat de Girona, Girona, Spain; Digestive System Service, Hospital Universitari Mútua de Terrassa, Terrassa, Spain; Centre for Biomedical Research in Network Hepatic and Digestive Diseases CIBEREHD, Inflammatory Bowel Diseases, Badalona, Spain; Proteomics and Metabolomics Laboratory, Research Directorate, General Hospital of Mexico, “Dr. Eduardo Liceaga”, Mexico City, Mexico; High Content Genomics and Bioinformatics Unit, IGTP, Badalona, Spain; Hospital Universitari de Tarragona Joan XXIII. Institut d′Investigació Sanitària Pere Virgili, Universitat Rovira i Virgili, Tarragona, Spain; Germans Trias i Pujol University Hospital, Badalona, Spain

**Keywords:** Crohn’s disease, Post-operative recurrence, Microbiota, Multi-omic, Porphyromonadaceae, Epithelial barrier

## Abstract

**Background:** Crohn’s disease (CD) is a chronic inflammatory disorder of the gastrointestinal tract characterized by high post-operative recurrence (POR) rates, reaching up to 90% within one year. Current clinical and endoscopic predictors show limited accuracy.

**Objective:** This study aimed to identify molecular mechanisms associated with POR at the time of surgery through integrated transcriptomic and bacteriomic analyses of ileal tissue.

**Design:** Ileal samples were obtained during surgery from 20 patients with CD and 10 inflammatory bowel disease–free controls, with an independent validation cohort of 49 patients with CD. POR was evaluated every six months using ileocolonoscopy and defined by Rutgeerts score. Host gene expression and tissue-associated microbiome profiles were integrated using correlation and pathway enrichment analyses to uncover host–microbe interactions linked to POR.

**Results:** In the inflamed mucosa of patients who developed endoscopic POR, we identified a novel immune interaction involving the Porphyromonadaceae family, mainly *Parabacteroides gordonii*, which was slightly depleted. This depletion was associated with downregulation of epithelial barrier and tissue repair genes, including *TFF1* and *LSR*, findings confirmed in the validation cohort. Porphyromonadaceae abundance positively correlated with short-chain fatty acid levels, particularly propionate. Additionally, omics integration revealed an association between Xanthomonadaceae and increased expression of *ITGAM*, a gene involved in neutrophil activation.

**Conclusion:** These results highlight microbial–host gene interactions associated with POR. The pathogenic ITGAM-driven immune signature and the protective Porphyromonadaceae–*TFF1*–propionate axis supporting epithelial integrity may enable microbiome-informed prognostic tools and therapeutic strategies for CD POR.

- **What is already known on this topic:** Post-operative recurrence in Crohn’s disease is linked to microbial dysbiosis, particularly reduced diversity and expansion of Enterobacteriaceae. However, how microbial changes translate into host molecular mechanisms driving POR remains unclear.
- **What this study adds:** This prospective multi-omic study identifies a disrupted Porphyromonadaceae–SCFA–epithelial barrier axis and the participation of neutrophil responses in patients who develop POR at surgery time.
- **How this study might affect research, practice or policy:** The findings provide mechanistic targets for microbiome-informed risk stratification and prevention of POR. They support development of microbial or metabolite-based interventions aimed at restoring epithelial barrier function after surgery.

## Introduction

Crohn’s disease (CD) is a chronic inflammatory disorder of the gastrointestinal tract characterized by relapsing inflammation and periods of remission^1^. Despite major advances in medical therapy, including biologics and small molecules, up to two-thirds of patients ultimately require intestinal resection due to refractory or complicated disease^2^. Surgery, however, is not curative, and up to 90% of patients develop post-operative recurrence (POR) within the first year^3^. POR represents a significant clinical challenge, as it may require repeated surgeries and worsens the patient’s long-term prognosis and quality of life^4^.

Current clinical predictors of POR, such as smoking, penetrating disease, perianal involvement, and prior resections, have limited accuracy, and the molecular mechanisms driving POR remain poorly understood^5^. Although thiopurines and anti-TNF agents reduce POR risk, a substantial proportion of patients develop POR despite prophylaxis^3^. Thus, there is an unmet need to better understand the molecular drivers of POR, as prerequisite for improving the patients’ management.

Emerging evidence suggests that POR arises from a complex interplay between host genetics, immune responses, and the intestinal microbiota. GWAS have identified genetic variants associated with CD susceptibility and POR, including *NOD2, ATG16L1, CXCR3* and *IL23R*, implicating pathways related to microbial sensing, autophagy, and immune regulation^6–8^. In addition, transcriptomic studies have further identified altered expression of genes involved in epithelial barrier function, autophagy, and cytokine signaling—including TNF-α, IFN-γ, and IL-17A—in patients who develop early POR ^6,9–11^

However, genetic susceptibility and expression also affect host–microbiota interactions, contributing to dysbiosis. CD-associated dysbiosis is characterized by depletion of beneficial commensals, such as *Faecalibacterium prausnitzii*, and expansion of pathobionts, including adherent-invasive *Escherichia coli* (AIEC), which have been linked to disease severity and POR^12–14^. Beyond changes in microbial composition, the microbiota not only interacts with the host’s immune system but also influences gene expression through mechanisms such as epigenetic modification^15^.

Integrating host transcriptomics with bacteriome profiling offers a powerful approach to dissect host–microbiota interactions at the mucosal interface to explore the molecular mechanisms underlying POR. Previous multi-omic studies have demonstrated that microbial communities influence mucosal gene expression and inflammatory pathways. However, such integrative approaches have not been systematically applied to the study of POR in CD^15,16^.

Given the complexity of host–microbiome interactions in CD, we hypothesized that integrating mucosal transcriptomic and bacteriome data would identify biologically coherent host–microbiota interactions associated with susceptibility to POR. Using a prospective, mechanistic discovery framework, we analyzed inflamed and microscopically uninflamed ileocolonic resection specimens from patients with CD, integrating gene expression, mucosal microbiota composition, and SCFA profiles. Our aim was to uncover microbiota-host biological function interactions relevant to POR and to identify candidate targets for personalized prevention of this complication.

## Material and methods

### Study design and patient cohorts

Inflamed and microscopically uninflamed ileal tissue samples were prospectively collected from 20 adults with CD undergoing ileocolonic resection and from 10 IBD-free controls undergoing right hemicolectomy for colon cancer with normal ileal histology, as previously described^6^. CD diagnosis followed Lennard-Jones criteria^17^, and disease phenotype was defined using the Montreal classification^18^. Patients with terminal ileostomy were excluded. Demographic, clinical, and treatment data were systematically recorded. Patients included in the study were not involved in the production of this research.

The inception cohort was designed for mechanistic discovery rather than predictive modeling. To validate prioritized findings, an independent cohort of 49 additional CD patients was included using identical selection criteria.

All patients underwent ileocolonoscopy 6–12 months after surgery and were classified according to the Rutgeerts score^19^ as non-POR (i0–i1) or POR (i2–i4). In the validation cohort, POR was further stratified as mild (i2) or severe (i3–i4).

Samples were stored in RNAlater for nucleic acid analyses or frozen at −80°C for SCFA quantification and housed at the IGTP Biobank following EQUATOR Network and BRISQ guidelines^20^. Procedures adhered to the Declaration of Helsinki and the Belmont Report^21^. The study was approved by the institutional ethics committee and all participants provided informed consent.

### Bioinformatic analysis of gene expression profiling

Gene expression profiling was performed in 40 mucosal samples from the inception cohort (inflamed and uninflamed), 10 control samples, and four technical replicates. Procedures are detailed in Supplementary Methods S1–S2. Differential expression analyses comparing controls, non-POR, and POR groups were performed using Kruskal–Wallis tests with Dunn’s post hoc correction (FDR ≤ 0.05; |log₂FC| ≥ 0.2). A significance score was calculated by multiplying the −log_10_(FDR) to the log_2_FC. A permissive FC threshold was applied to capture coordinated but moderate transcriptional shifts typical of heterogeneous intestinal tissue. Volcano plots were performed. Analyses were done in R (v4.4.2).

### Mucosal bacteriome sequencing and analysis

Detailed version of this methodology can be found in Methods S3-S4. Briefly, DNA was extracted from intestinal tissue using the Nucleospin® Tissue Kit. The V4 region of bacterial 16S rRNA was sequenced using a modified Illumina protocol^22^ on the Illumina MiSeq platform. Quality control and taxonomic assignment in operational taxonomic units (OTUs) were performed using QIIME2, DADA2, and the SILVA 138-99 database^23^.

Alpha and beta diversity metrics were computed using Shannon, Simpson, Bray–Curtis, and Jaccard indices. Differential abundance analyses were conducted with DESeq2 (adjusted p ≤ 0.05; |log₂FC| ≥ 1.5). Batch effects for the two cohorts were assessed^24^. The correlation analysis between sequencing depth (raw read pairs) and observed species richness ruled out potential technical biases (Figure S1). Sequencing data are available at NCBI SRA (PRJNA1241201).

### Correlation analyses between microbiome and transcriptome

Correlation and regression analyses were performed between normalized transcriptomic data and relative selected OTU/families abundances. Linear regressions were restricted to |Kendall’s τ| > 0.3 and > 3 non-missing samples.

### Transcriptomic pathway enrichment analysis

Pathway enrichment analysis (PEA) was conducted using clusterProfiler in R (v4.2.2), based on over-representation analysis (ORA) drawing from GO, KEGG, and Reactome databases^25^. ORA was used on significant transcripts (separated by inflamed and uninflamed zones) in comparisons against IBD-free controls, excluding POR vs non-POR due to low significant genes. Exclusive genes (non-coincident between POR groups) were analyzed, both globally and separated by direction of regulation. ORA was also used in the bacteria-transcript correlations. Significant pathways (adjusted p ≤ 0.05, 10–300 genes, ≥ 3 input genes) were grouped by functional similarity. Results were visualized using barplots.

### Graphical representation of the integrative networks in the inception cohort

Cytoscape software (U.S., v3.10.0) was used to graphically build up integrative networks in each phenotype and intestinal zone. Nodes (transcripts, taxa, pathways) were color-coded and shaped by type. Gene symbols are given for each transcript and intensity of the color (red: under-expressed, blue: over-expressed) is a value calculated using both the FC and the FDR. Inner and border colors show POR/non-POR vs controls and inter-group comparisons, respectively. Edge color represented correlation direction and strength. PEA aggrupation nodes border width corresponds to the highest −log_2_(q-value) of each pathway aggrupation in biological processes.

### Candidate gene prioritization for qRT-PCR replication in the validation cohort

Transcripts for qRT-PCR validation were selected based on: transcripts correlating to a bacterial group and enriched in PEAs that were: a) differentially expressed towards the controls and non-coincident and b) enriched in the same or similar pathways from the transcriptome analyses. The flowchart of the whole omic integrative study is represented in Figure 1. Full methodology can be found in Methods S1 for RNA extraction and S5 for qRT-PCR conditions.

**Figure 1.**
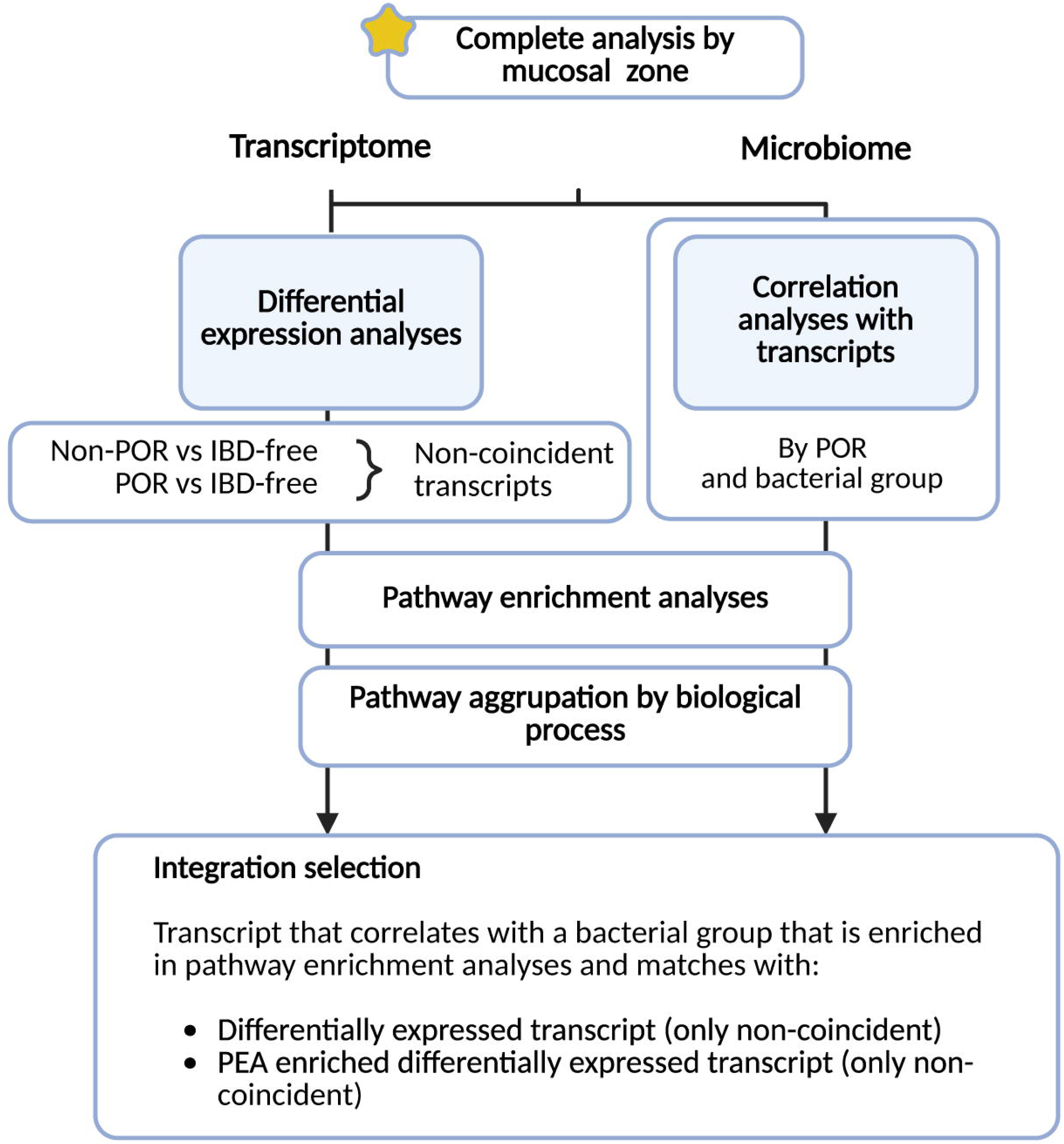
Flowchart of the integrative multi-omic analysis to elucidate the bacterial involvement in CD POR. Transcriptome data was analyzed for differential expression. Microbiome data was correlated to the expression level of transcripts by POR and mucosal inflammation. Transcripts were inputted in PEA using GO, KEGG and Reactome databases and the result pathways were grouped by biological processes. Finally, a score of transcript selection for the validation in a corroboration cohort was built and applied to the analysis. CD, Crohn’s disease; GO, gene ontology; KEGG, Kyoto Encyclopedia of Genes and Genomes; OTU, operational taxonomic unit; PEA, pathway enrichment analysis; POR, post-operative recurrence.

Selected transcripts were *ITGAM*, *LSR* and *TFF1*, and barrier-related genes (*CLDN1*, *CLDN2*, *MUC2*, *TJP1*, *OCLN*; find Taqman®Assays references in Table S1).

### SCFA quantification

SCFAs levels were quantified by gas chromatography-mass spectrometry (GC-MS)^26^. Briefly, using a Shimadzu GCMS-QP2010 Plus (Shimadzu, Kyoto, Japan) system and TRB-FFAP column (Teknokroma Analítica SA, Barcelona, Spain). Data was acquired by the GCMSsolution software. Reference mixture of SCFA (CRM46975, Merck Life Science S.L.U.) was used.

### Standard statistics

Demographic and clinical were analyzed using Student T test or Χ^2^ for numerical or categorical variables, respectively. Data normality was assessed using the Shapiro-Wilk test. Kruskal-Wallis/Dunn’s test and Mann-Whitney tests were used for independent variables, whereas Wilcoxon test for dependent ones. Correlation analyses were done by Pearson or Spearman tests for parametric and non-parametric data, respectively. All analyses were performed in R (v4.2.2).

## Results

### Clinical characterization of inception and validation cohorts

Patients with CD in the inception cohort were younger, had lower BMI, and smoked more frequently than controls (Table 1). When comparing the validation to the inception cohort, the proportion of non-smokers and POR rates were higher. No significant demographic differences were observed between POR groups in either cohort (Tables S2-S3). However, within the POR group in the validation cohort, immunomodulators were observed to be more frequently used in mild POR whereas anti-TNF therapy in severe POR (Table S4).

**Table 1.**
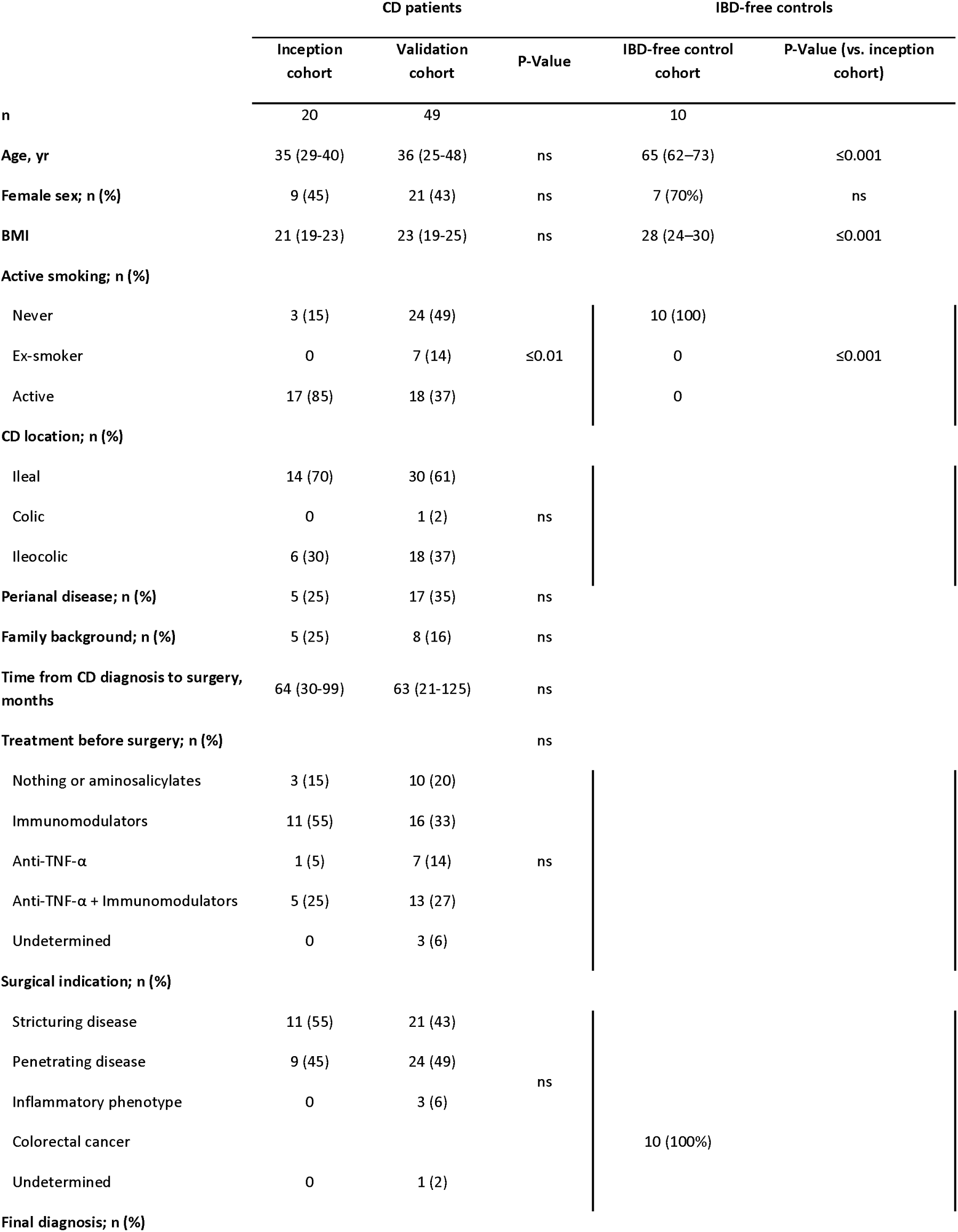
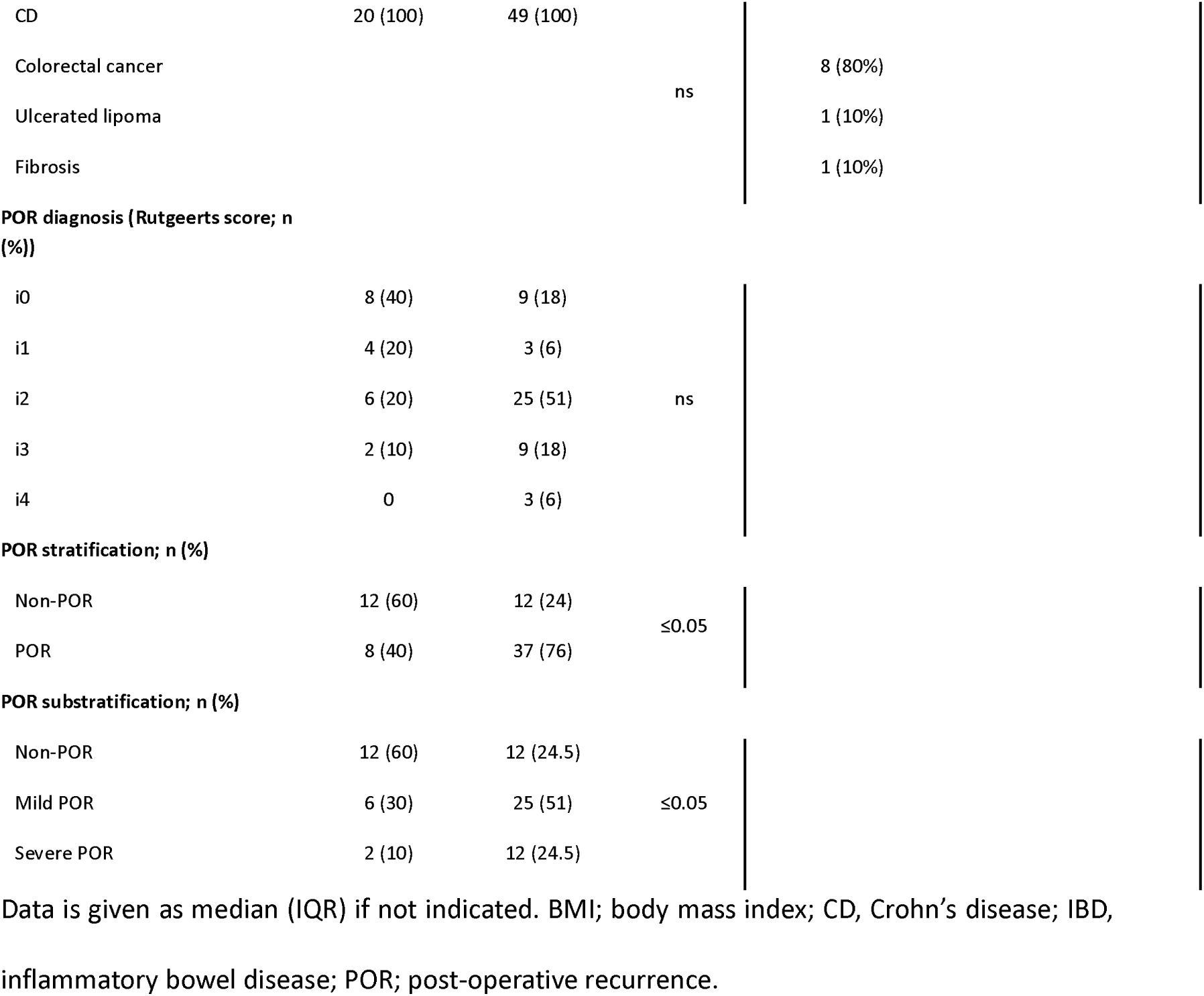
Demographic and epidemiological features of the inception cohort compared to the validation cohort and IBD-free controls.

### PEAs reveal epithelial barrier-related processes linked to POR

Direct transcriptomic comparisons between POR and non-POR patients identified differentially expressed transcripts (Table 2). Then, the comparisons towards controls pointed out distinct sets of non-coincident differentially expressed transcripts in both inflamed and uninflamed mucosa (Table 3 for the top scoring transcripts and Tables S5-S6 for whole results). Notably, POR was associated with a substantially higher number of altered transcripts, particularly under-expression in both uninflamed and inflamed mucosa. Results were also graphically represented in volcano plots (Figures S2-S3).

**Table 2.** Differentially expressed transcripts between POR and non-POR in both inflammatory statuses.

|  | GEO | Refseq/GenBank | Symbol | FDR | log<sub>2</sub>FC |
| --- | --- | --- | --- | --- | --- |
| Uninflamed | GE790713 | AK093292 | ATP11A-AS1 | 0.031 | -0.21 |
|  | GE60082 | NM_004059 | KYAT1 | 0.048 | -0.22 |
|  | GE529037 | AK124131 | PWRN2 | 0.011 | -0.95 |
|  | GE486546 | NM_002854 | PVALB | 0.029 | -1.63 |
| Inflamed | GE478868 | NM_001890 | CSN1S1 | 0.033 | 0.31 |
|  | GE79286 | NM_022837 | PITX1 | 0.004 | 0.31 |
|  | GE81172 | NM_002653 | NA | 0.003 | -0.23 |

FC, fold change; FDR, false discovery rate.

**Table 3.**
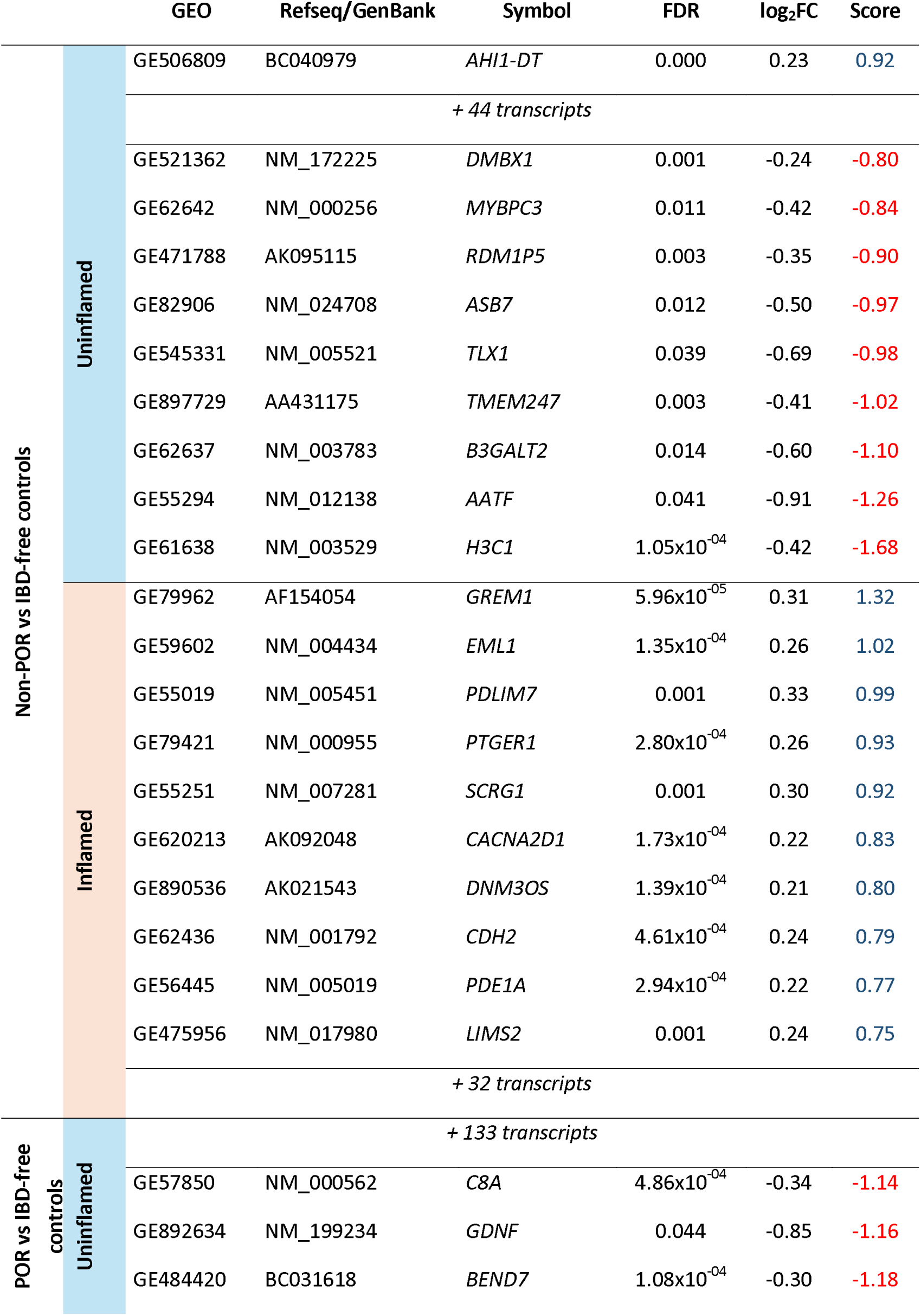
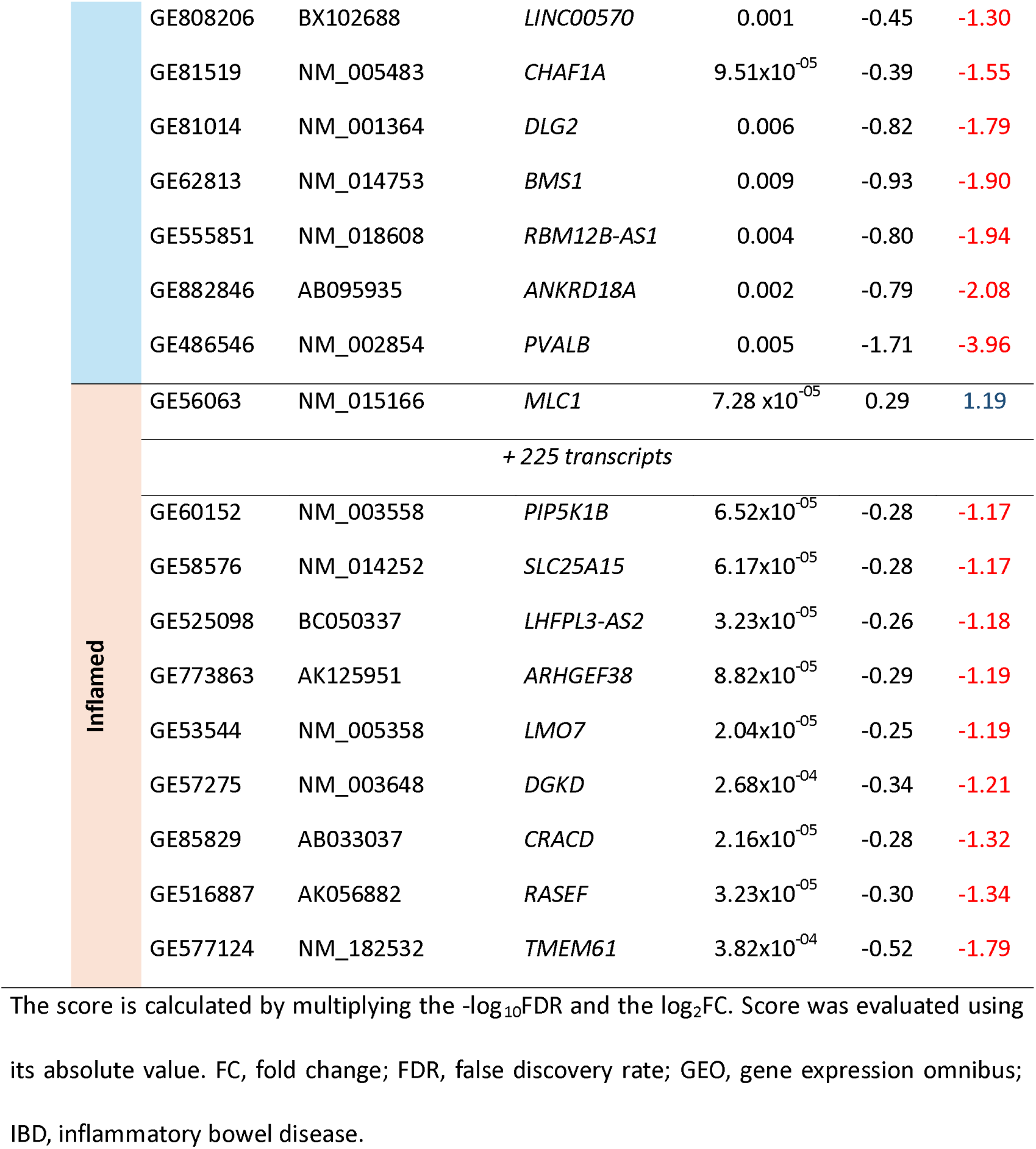
Top 10-scoring non-coincident differentially expressed transcripts between non-POR/POR and IBD-free controls in both inflammatory statuses.

PEAs revealed that in non-POR (Table 4), the uninflamed mucosa was enriched in extracellular matrix-related pathways, epithelial-mesenchymal transition and muscle formation. Of note, *MYBPC3* was linked to muscle genesis. In the inflamed mucosa, xenobiotic metabolism was predominant, along with unique enrichments in synapse function (*CACNA2D1*) and cell adhesion and tight junctions (*CDH2* and *LIMS2*).

**Table 4.**
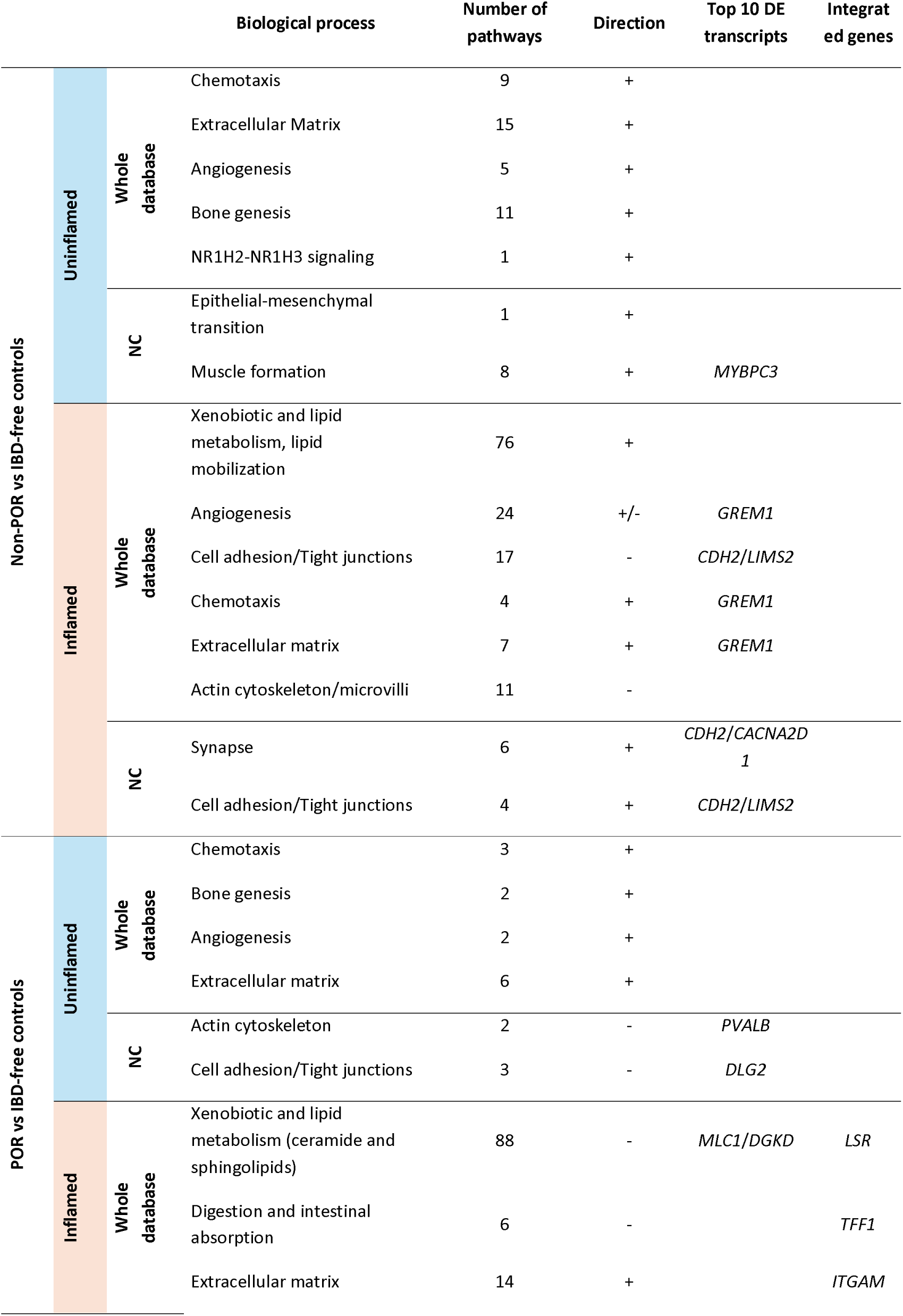
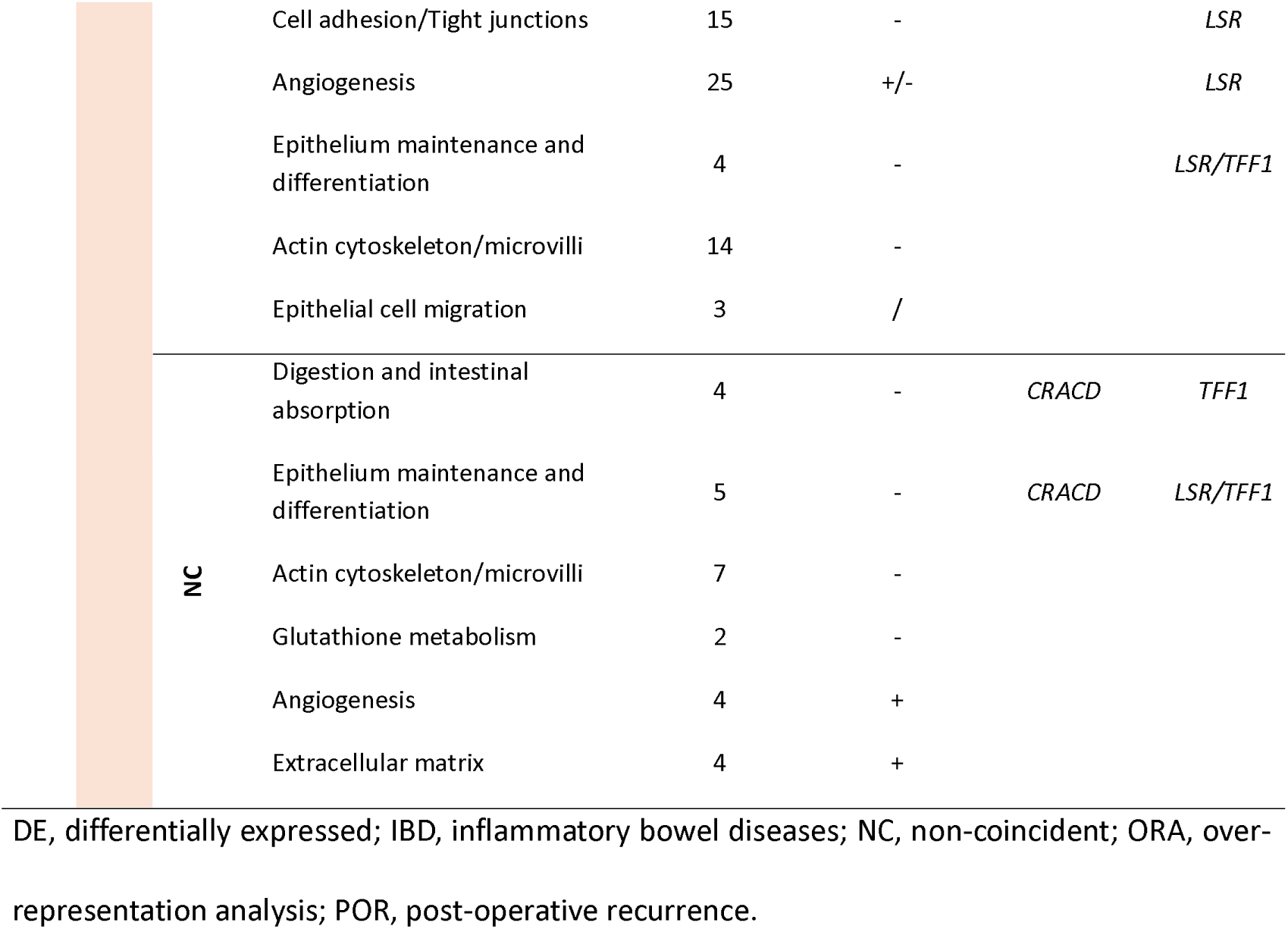
Significant biological processes enriched in the non-POR and POR versus the IBD-free controls in the ORA.

In POR, the uninflamed mucosa again showed enrichment of extracellular matrix related pathways, while Actin cytoskeleton (*PVALB*) and cell adhesion/tight junction pathways were exclusively enriched here. In the inflamed mucosa, xenobiotic metabolism was also the most represented topic -though negatively enriched-, followed by angiogenesis, tight junctions, epithelial maintenance and digestion-related functions. Key transcripts included *MLC1*, *DGKD*, and *CRACD*, the latter enriched in intestinal absorption and barrier.

### Microbial diversity is reduced in POR

Microbial alpha and beta diversity were analyzed to gain insights into the overall microbial diversity and community composition. Alpha diversity was reduced in patients with CD compared with controls and was lowest in POR (Figure S4), with no difference between mild and severe POR (Figure S5). Beta diversity analyses showed no clear clustering by demographic, clinical and batch variables (Figure S6). Altogether suggesting that POR is associated with a more unbalanced and less diverse microbial ecosystem, but the lack of clear clustering raises the need for other alternative or more integrative analytical approaches for better resolution.

Severe POR shows a marked increase of Clostridiacieae and Enterobacteriaceae together with a loss in Porphyromonadaceae and Ruminococcaceae bacterial families.

Although global microbial diversity did not differ markedly across groups, specific OTUs or families showed significant shifts in abundance associated with POR (Figures 2 and 3; see Figure S7 for frequence heatmaps showing the unclassified bacteria). These changes highlighted two opposing microbial patterns linked to POR: enrichment of taxa associated with inflammatory responses and depletion of bacterial families linked to epithelial homeostasis and SCFA production.

**Figure 2.**
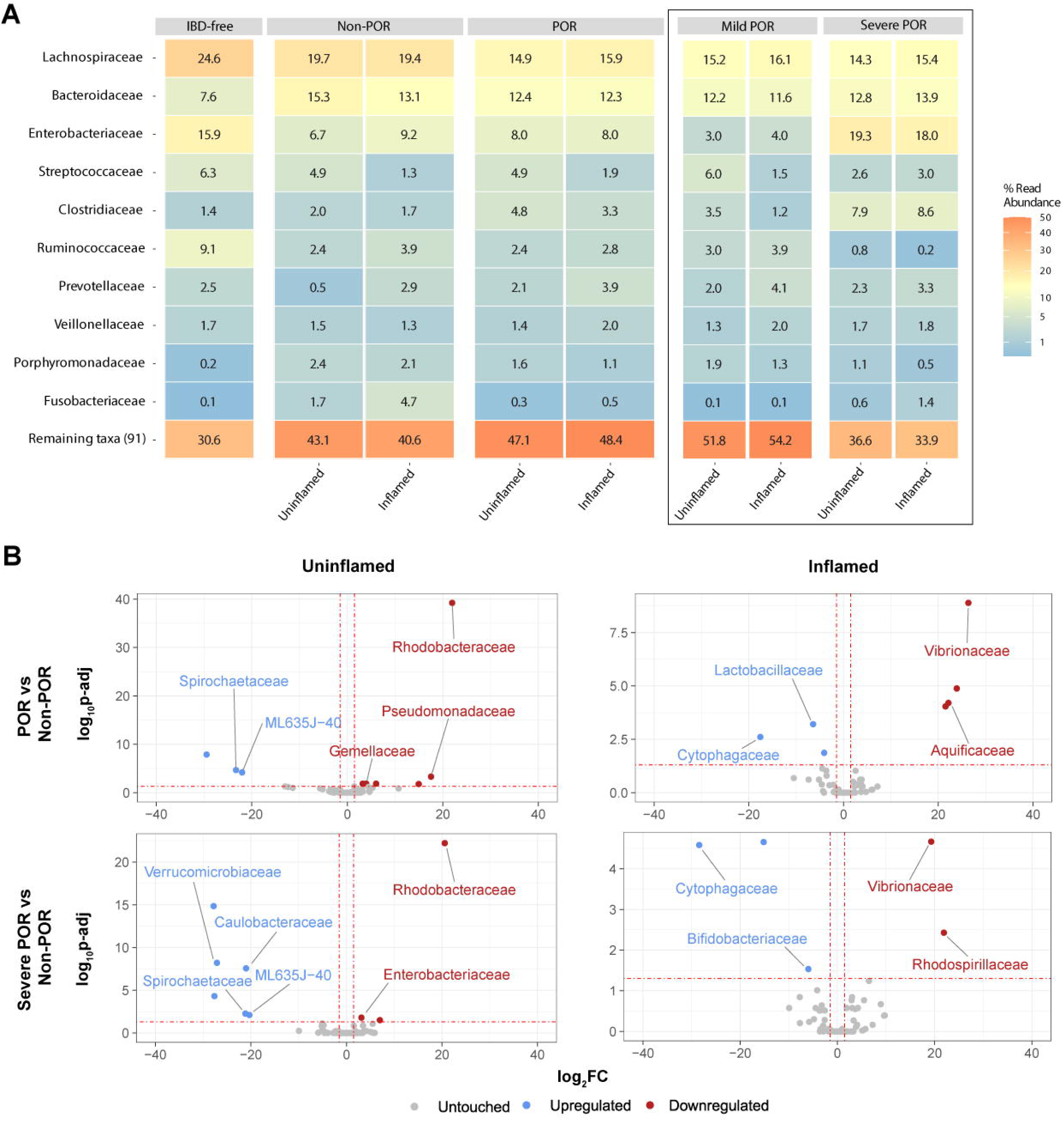
Alterations in bacterial family-level abundance associated with POR in CD. (A) Heatmap showing the relative abundances (%) of the top 10 most prevalent bacterial families across all samples, stratified by POR phenotype and mucosal inflammation status. (B) Volcano plots illustrate the DESeq2 differential abundance analysis of bacterial families comparing non-POR versus POR or severe POR, according to mucosal inflammation. Families with significantly higher relative abundance in POR are shown in red, whereas those enriched in non-POR are shown in blue. The y-axis represents the −log_10_(adjusted P-value) and the x-axis indicates the log_2_FC. Only taxonomically classified and statistically significant families are indicated in the plots. Rhodobacteriaceae, Pseudomonadaceae and Gemellaceae were significantly higher in POR compared to the non-POR patients in the uninflamed mucosa, while Vibrionaceae and Aquificaceae were so in the inflamed zone. On the other hand, Spirochaetaceae levels were higher in non-POR in the uninflamed mucosa, while Cytophagaceae and Lactobacillaceae were increased in the inflamed mucosa. Verrucomicrobiaceae and Bifidobacteriaceae families were notably depleted in the severe POR, Verrucomicrobiaceae in the uninflamed mucosa, whereas Bifidobacteriaceae in the inflamed one. Interestingly, Enterobacteriaceae was higher in severe POR in the uninflamed zone. CD, Crohn’s disease; FC, fold-change; POR, post-operative recurrence.

**Figure 3.**
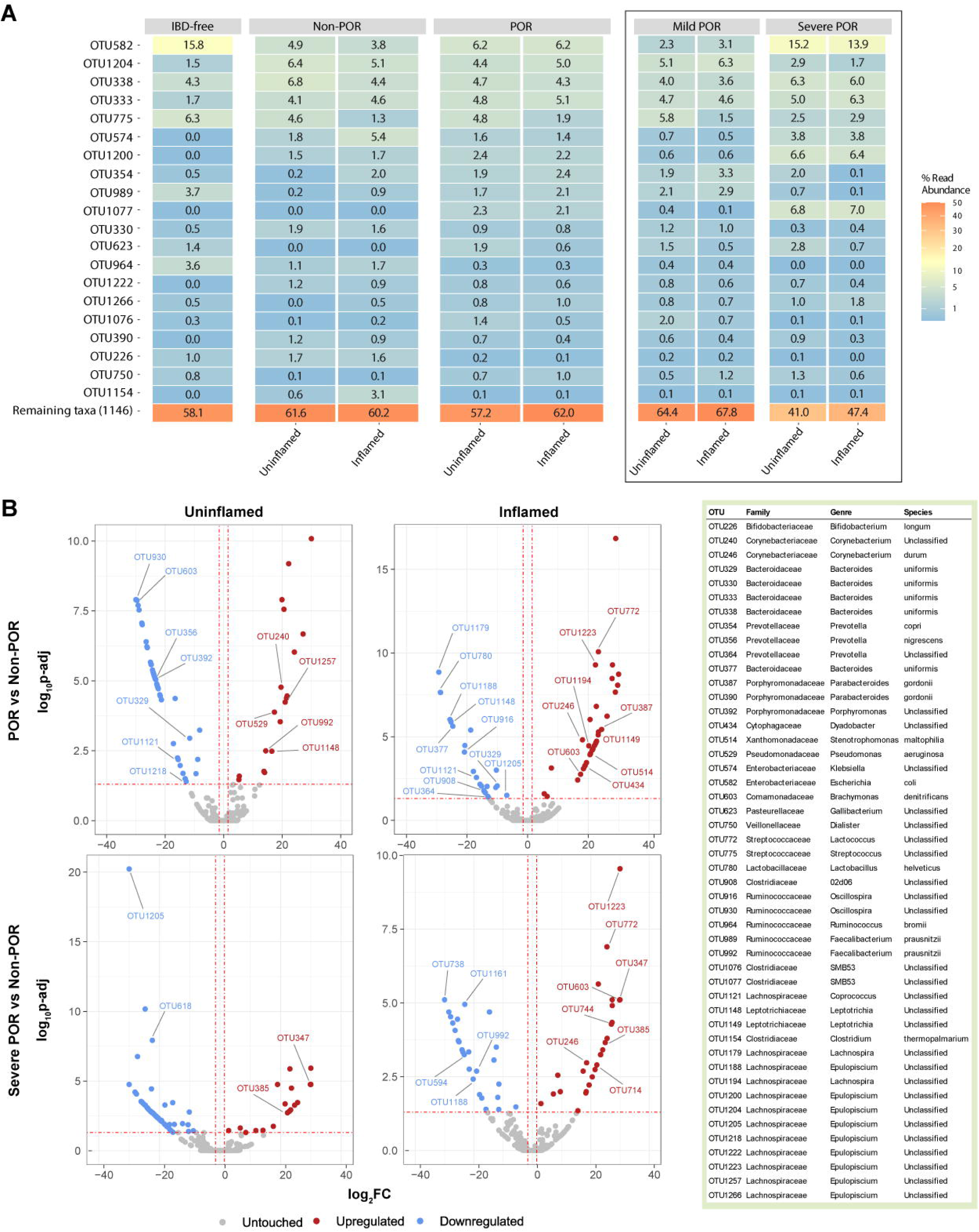
Alterations in the bacterial OTU abundance associated with POR in CD. (A) Heatmap showing the relative abundances (%) of the top 20 most prevalent OTUs across all samples, stratified by POR phenotype and mucosal inflammation status. (B) Volcano plots illustrate the DESeq2 differential abundance analysis of bacterial OTUs comparing non-POR versus POR or severe POR, according to mucosal inflammation. OTUs with significantly higher relative abundance in POR are shown in red, whereas those enriched in non-POR are shown in blue. The y-axis represents the −log_10_(adjusted P-value) and the x-axis indicates the log_2_FC. Only taxonomically classified and statistically significant OTUs are indicated in the plots. CD, Crohn’s disease; FC, fold-change; OTU, operational taxonomic unit; POR, post-operative recurrence.

Lachnospiraceae, Bacteroidaceae and Enterobacteriaceae were the most abundant families (Figure 2A). *Lachnospiraceae* abundance was lower in CD than in the IBD-free controls and showed a trend toward further reduction with POR severity. In contrast, *Bacteroidaceae* were more prevalent in CD samples compared to IBD-free controls. Strikingly, *Enterobacteriaceae* and *Clostridiaceae* were enriched in severe POR in both uninflamed (19.3% and 7.9%, respectively) and inflamed mucosa (18% and 8.6%). On the other hand, Ruminococcaceae and Porphyromonadaceae –particularly represented by *Parabacteroides gordonii* (mainly OTU386 and OTU390)- were reduced in severe POR. Although some families did not show great frequencies, they showed significant associations (Figure 2B). The whole results of the differentially expressed families towards the IBD-free controls and between POR groups can be found in Tables S7 and S8, respectively.

At the OTU level, Lachnospiraceae Epulopiscium (OTU1204) and *E. coli* (OTU582), were found to be the most abundant ones (Figure 3A). Moreover, the OTU1204 and OTU1205 (both L. Epulopiscium) were lower only in the uninflamed mucosa of the severe POR when compared to non-POR (adjusted p = 0.059 and 0.032, respectively). In contrast, OTU1200 (L. Epulopiscium) showed a trend to be increased in the severe POR compared to the non-POR (adjusted p = 0.089 in the uninflamed zone).

Notably, the *E. coli* OTU582 was found at low levels in non- and mild POR but significantly increased in severe POR and IBD-free controls (DEseq2 non-POR vs severe POR adjusted p = 0.036 and 0.026, and mild POR vs severe POR adjusted p = 0.008 and 0.002; for the uninflamed and inflamed zone, respectively; Figure 3B). Importantly, the OTU989 (*Faecalibacterium prausnitzii*) was 4-fold reduced in severe states of POR in inflammation when compared to the non-POR (adjusted p = 0.066), consistent with its known anti-inflammatory role. OTU1077 (Clostridiaceae SMB53) showed a marked increase (∼7%) in severe POR in both zones. The whole results of the differentially expressed OTUs towards the IBD-free controls and between POR groups can be found in Tables S9 and S10, respectively.

### Bacterial associations with host transcriptomic pathways in POR phenotypes

To move beyond taxonomic associations, bacterial groups were linked to host transcriptomic pathways using correlation-based enrichment analyses, enabling functional interpretation of host–microbiota interactions.

Bacterial families and OTUs showing > 80 significant correlations were selected for PEA, alongside OTU582 due to its prior relevance. In non-POR, Actinomycetales negatively correlated with mitochondrial respiration, and protein translation, whereas the Porphyromonadaceae family, and OTU1184 (L. Epulopiscium) showed positive correlations with mitochondrial functions. In the inflamed zone, the mitochondrial respiration remained prominent (21 pathways) negatively correlated with OTU1240 (L. Epulopiscium). Other bacterial groups were correlated to cytoskeleton, extracellular matrix or cell migration functions, among others, as summarized in Table 5.

**Table 5.**
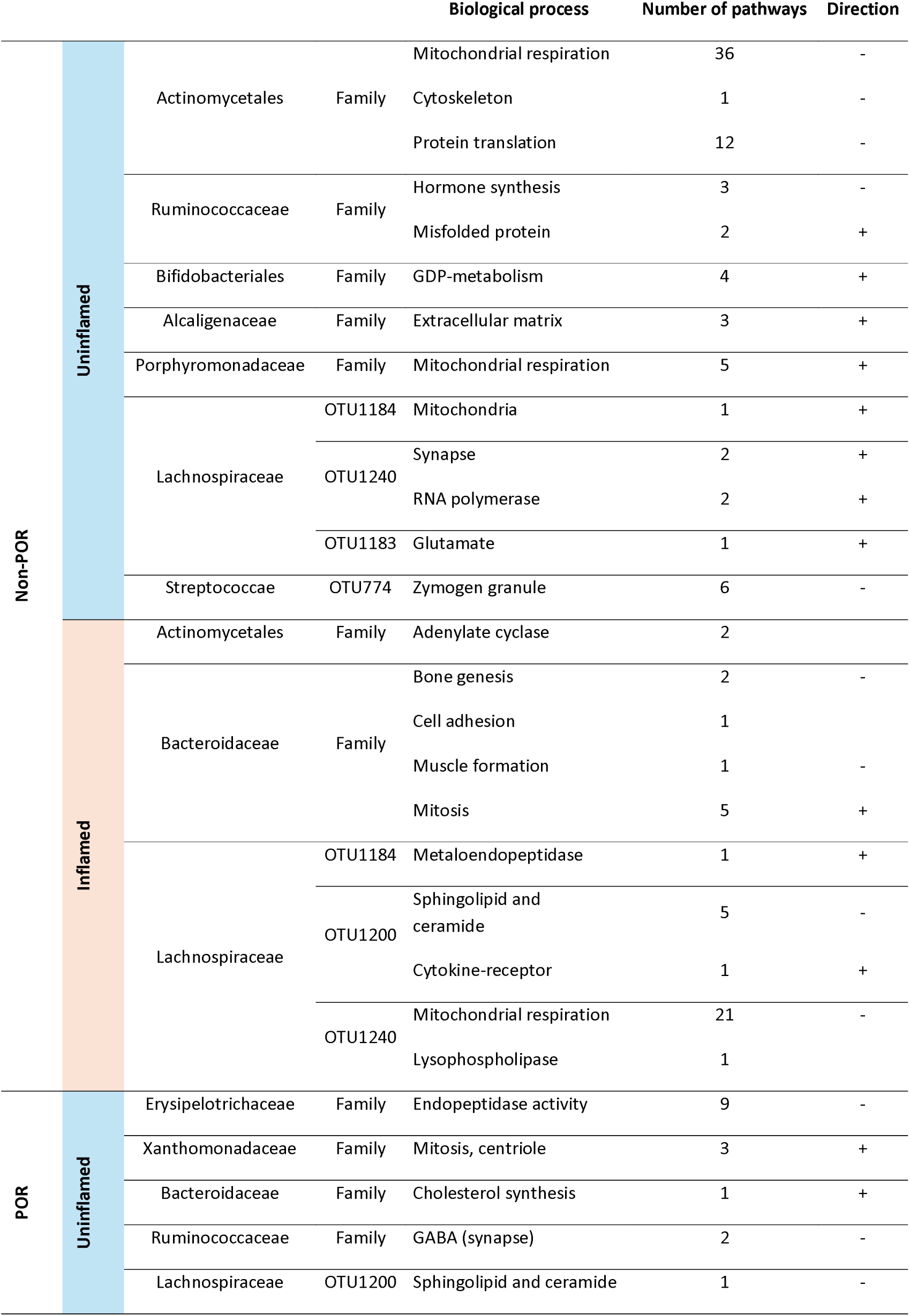
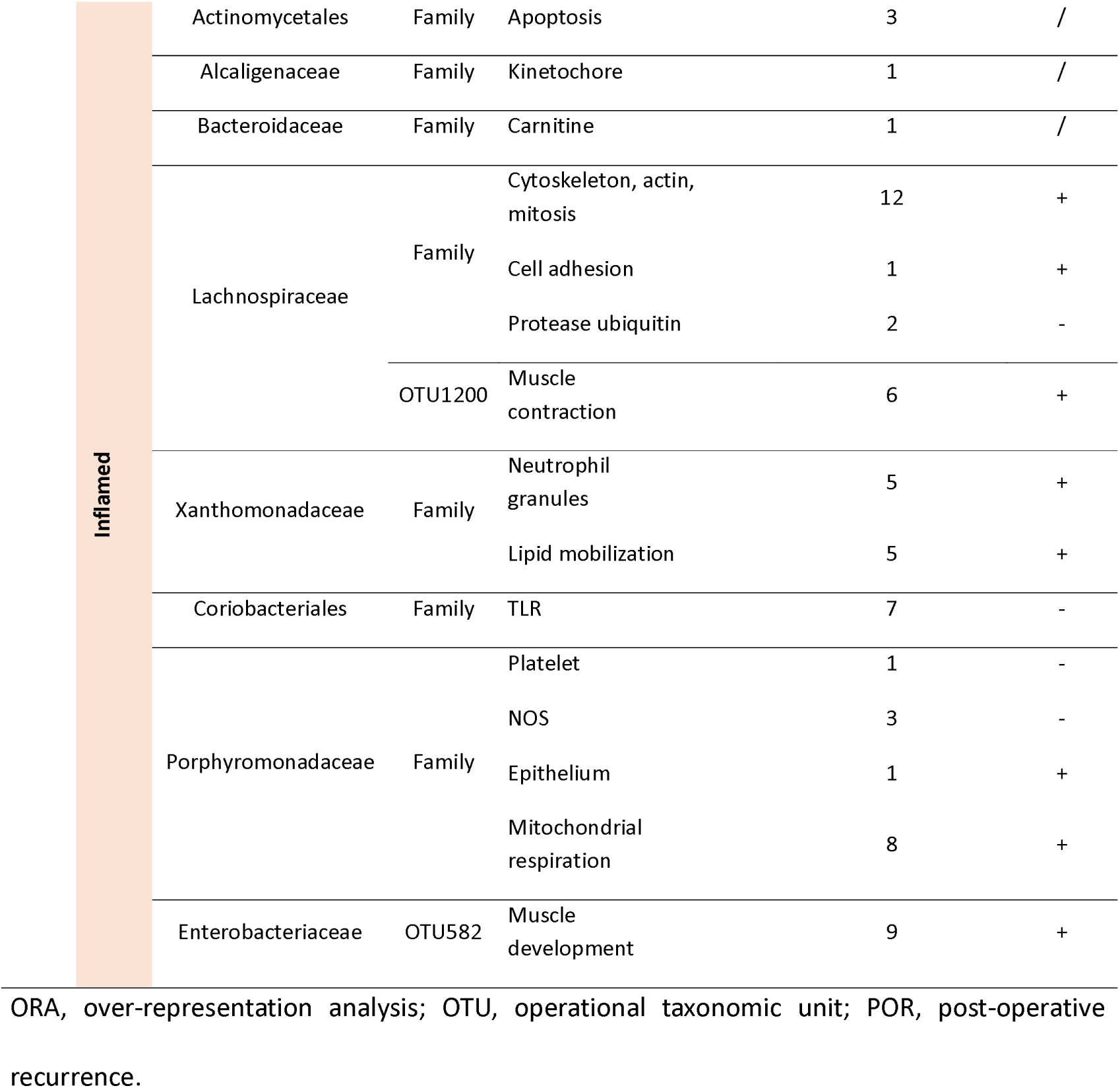
Enriched biological processes correlating with bacterial families or OTUs through ORA in each POR phenotype and mucosal inflammatory status.

|  |  |  |  | Biological process | Number of pathways | Direction |
| --- | --- | --- | --- | --- | --- | --- |
| Non-POR | Uninflamed | Actinomycetales | Family | Mitochondrial respiration | 36 | - |
|  |  |  |  | Cytoskeleton | 1 | - |
|  |  |  |  | Protein translation | 12 | - |
|  |  | Ruminococcaceae | Family | Hormone synthesis | 3 | - |
|  |  |  |  | Misfolded protein | 2 | + |
|  |  | Bifidobacteriales | Family | GDP-metabolism | 4 | + |
|  |  | Alcaligenaceae | Family | Extracellular matrix | 3 | + |
|  |  | Porphyromonadaceae | Family | Mitochondrial respiration | 5 | + |
|  |  | Lachnospiraceae | OTU1184 | Mitochondria | 1 | + |
|  |  |  | OTU1240 | Synapse | 2 | + |
|  |  |  |  | RNA polymerase | 2 | + |
|  |  |  | OTU1183 | Glutamate | 1 | + |
|  |  | Streptococcae | OTU774 | Zymogen granule | 6 | - |
|  | Inflamed | Actinomycetales | Family | Adenylate cyclase | 2 |  |
|  |  | Bacteroidaceae | Family | Bone genesis | 2 | - |
|  |  |  |  | Cell adhesion | 1 |  |
|  |  |  |  | Muscle formation | 1 | - |
|  |  |  |  | Mitosis | 5 | + |
|  |  | Lachnospiraceae | OTU1184 | Metaloendopeptidase | 1 | + |
|  |  |  | OTU1200 | Sphingolipid and ceramide | 5 | - |
|  |  |  |  | Cytokine-receptor | 1 | + |
|  |  |  | OTU1240 | Mitochondrial respiration | 21 | - |
|  |  |  |  | Lysophospholipase | 1 |  |
| POR | Uninflamed | Erysipelotrichaceae | Family | Endopeptidase activity | 9 | - |
|  |  | Xanthomonadaceae | Family | Mitosis, centriole | 3 | + |
|  |  | Bacteroidaceae | Family | Cholesterol synthesis | 1 | + |
|  |  | Ruminococcaceae | Family | GABA (synapse) | 2 | - |
|  |  | Lachnospiraceae | OTU1200 | Sphingolipid and ceramide | 1 | - |

|  |  |  |  |  |  |
| --- | --- | --- | --- | --- | --- |
| Inflamed | Actinomycetales | Family | Apoptosis | 3 | / |
|  | Alcaligenaceae | Family | Kinetochore | 1 | / |
|  | Bacteroidaceae | Family | Carnitine | 1 | / |
|  | Lachnospiraceae | Family | Cytoskeleton, actin, mitosis | 12 | + |
|  |  |  | Cell adhesion | 1 | + |
|  |  |  | Protease ubiquitin | 2 | - |
|  |  | OTU1200 | Muscle contraction | 6 | + |
|  | Xanthomonadaceae | Family | Neutrophil granules | 5 | + |
|  |  |  | Lipid mobilization | 5 | + |
|  | Coriobacteriales | Family | TLR | 7 | - |
|  | Porphyromonadaceae | Family | Platelet | 1 | - |
|  |  |  | NOS | 3 | - |
|  |  |  | Epithelium | 1 | + |
|  |  |  | Mitochondrial respiration | 8 | + |
|  | Enterobacteriaceae | OTU582 | Muscle development | 9 | + |

ORA, over-representation analysis; OTU, operational taxonomic unit; POR, post-operative recurrence.

In POR, the Erysipelotrichaceae family negatively correlated with endopeptidase activity (uninflamed mucosa). The Porphyromonadaceae were positively related again to mitochondrial respiration and epithelium maintenance; the OTU582 to muscle genesis; and Xanthomonadaceae family, mainly the *Stenotrophomonas maltophilia* species (OTU516), to neutrophil granules and lipid mobilization in the inflamed mucosa.

#### Host-microbiome interactions relevant to POR

Candidate host–microbiota interactions were prioritized through an integration strategy combining correlation strength, pathway co-enrichment and biological plausibility. The multi-omic integration identified key transcript-bacteria pairs for replication in the validation cohort (Table S11). Only those transcripts correlating to bacterial groups that enriched in pathways were considered. The selection was applied based on correlation strength and pathway co-enrichment. Only pairs from the inflamed zone of the POR phenotype fulfilled the conditions and were finally selected for further analysis.

A graphic interaction network highlighted these key pairs (full network in Figures S8). The *ITGAM* was positively correlated to Xanthomonadaceae, linked to ECM pathways and to neutrophil granules formation (Figure 4A & B). This family also correlates positively to the expression of genes as important for neutrophil activity as *NFKB1* and *MMP9*. This correlation was not corroborated in the validation cohort (Figure 4C). Nevertheless, the *ITGAM* expression was significantly higher in the POR group when compared with the non-POR one. Additionally, after the stratification of POR between mild and severe, the *ITGAM* was over-expressed in severe POR compared to non-POR.

**Figure 4.**
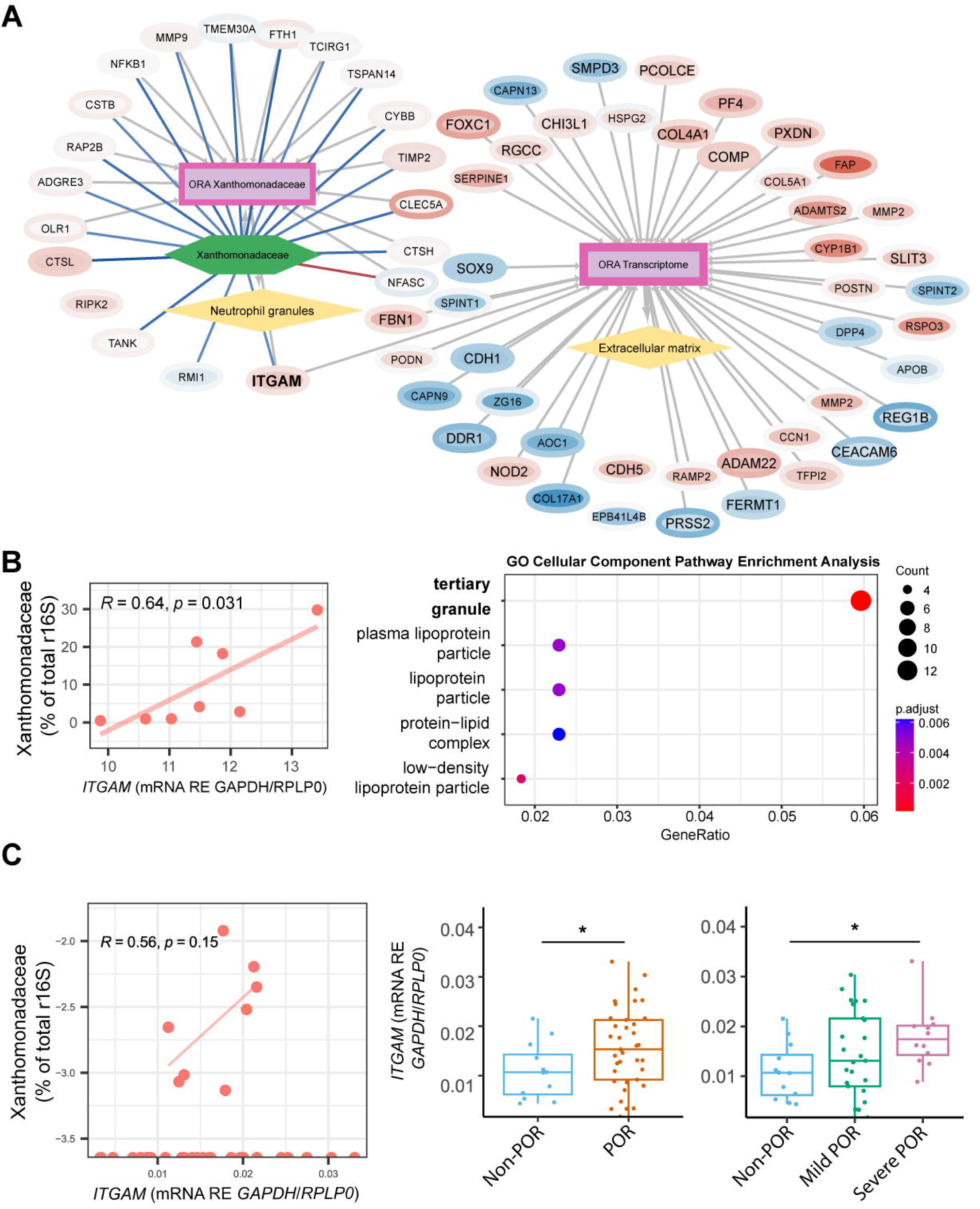
Xanthomonadaceae integrative transcriptome/microbiome network of the inflamed mucosa in patients with POR. (A) Graphical representation of the transcriptome/microbiome integration by Cytoscape. Gene symbols are given for each transcript and intensity of the color (red for under-expressed, blue for over-expressed) is a value calculated using both the FC and the adjusted P-value. Inner color refers to the comparison POR vs IBD-free controls, and border color refers to the comparison between POR vs non-POR. Font size is bigger for non-coincident transcripts. PEA aggrupation nodes border width corresponds to the highest −log_2_(q-value) of each aggrupation. Link line color between transcripts and bacteria indicate the intensity of the correlation (red for negative correlation, blue for positive). Line width between pathway and PEA aggrupation indicates number of enriched pathways grouped. Find website version in: https://doi.org/10.18119/N9390T. (B) Scatter plot depicting the *ITGAM* correlation to the Xanthomonadaceae family and the top 5 enriched pathways in the PEA using the GO cellular component database. For the scatter plot, the y-axis represents the bacterial family while the x-axis corresponds to gene expression. These contain a fitted line, and R^2^ and P-value are indicated in the plot. Correlation analyses were performed using Kendall statistical test. (C) Scatter plots depicting the correlation of the Xanthomonadaceae family with *ITGAM* gene expression and boxplots showing the *ITGAM* gene expression depending on the POR phenotype in the validation cohort. Boxplots y-axis corresponds to the gene expression, and the x-axis depicts each POR phenotype. Correlation test was performed using Pearson statistical analysis. Boxplots contain boxes indicating IQR, and bars, indicating maximum and minimum. The median point is depicted as a bar inside the box. Asterisks indicate significant differences between groups, as determined by the Kruskal-Wallis test with post-hoc Dunn’s test. Significance levels are: * P-value ≤ 0.05. GO, gene ontology; IBD, inflammatory bowel disease; IQR, interquartile range; ORA, over-representation analysis; PEA, pathway enrichment analysis; POR, post-operative recurrence; R, rho; r16S, ribosomal 16S, RE, relative expression.

On the other hand, Porphyromonadaceae correlated positively and strongly with *LSR* and *TFF1*, both involved in epithelial barrier and lipoprotein related functions (Figure 5A & B). These functions were in fact downregulated in POR in the inflamed zone. The validation in an independent cohort in the same kind of group of patients and mucosal inflammation confirmed the correlation between the *Porphyromonadaceae* and *TFF1* (R^2^ = 0.51, P-value = 0.003; Figure 5C). LSR also showed an interesting positive trend (R^2^ = 0.3, P-value = 0.09).

**Figure 5.**
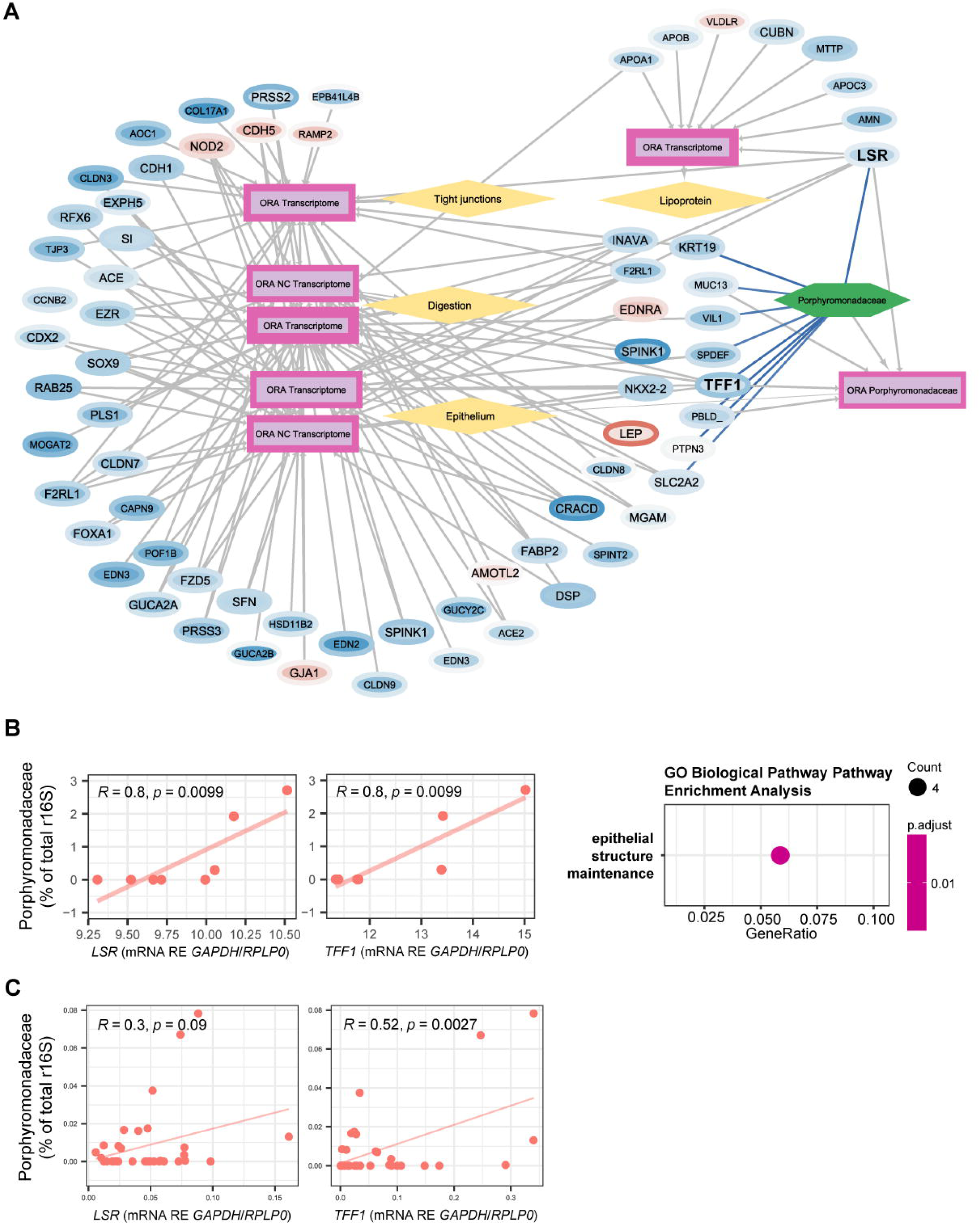
Porphyromonadaceae integrative transcriptome/microbiome network of the inflamed mucosa in patients with POR. (A) Graphical representation of the transcriptome/microbiome integration by Cytoscape. Gene symbols are given for each transcript and intensity of the color (red for under-expressed, blue for over-expressed) is a value calculated using both the FC and the adjusted P-value. Inner color refers to the comparison POR vs IBD-free controls, and border color refers to the comparison between POR vs non-POR. Font size is bigger for non-coincident transcripts. PEA aggrupation nodes border width corresponds to the highest −log_2_(q-value) of each aggrupation. Link line color between transcripts and bacteria indicate the intensity of the correlation (red for negative correlation, blue for positive). Line width between pathway and PEA aggrupation indicates number of enriched pathways grouped. Find website version in: https://doi.org/10.18119/N9ZK6M. (B) Scatter plot depicting the *TFF1* and *LSR* correlation to the Porphyromonadaceae family and the top 5 enriched pathways in the PEA using the GO cellular component database. Correlation analyses were performed using Kendall statistical test. For the scatter plots, the y-axis represents the bacterial family while the x-axis corresponds to the gene expression. Scatter plots contain a fitted line, and R^2^ and P-value are indicated in the plot. (C) Scatter plots representing the correlation of the Porphyromonadaceae family with *TFF1* and *LSR* gene expression in the validation cohort. Correlation test was performed using Pearson statistical analysis. GO, gene ontology; IBD, inflammatory bowel disease; ORA, over-representation analysis; PEA, pathway enrichment analysis; POR, post-operative recurrence; R, rho; r16S, ribosomal 16S, RE, relative expression.

### SCFA propionate correlates with Porphyromonadaceae and barrier function integrity

The SCFA analyses in the inflamed mucosa of CD patients revealed that propionate positively correlated with the abundance of *Porphyromonadaceae* (R^2^ = 0.49, P-value = 0.002; Figure 6A) and with the *TFF1* gene expression (R^2^ = 0.40, P-value = 0.013) and tendentially to LSR expression (Figure 6B). As of interest, only in POR, the propionate was also correlated to the intestinal barrier occludin gene expression (*OCLN*; R^2^ = 0.53, P-value = 0.027; see Figure 6C). Furthermore, *OCLN* expression was tendentially lower in POR and significantly lower in the severe POR phenotype compared to the non-POR. Together, these findings link microbial composition, mucosal propionate levels and epithelial barrier gene expression into a unified functional axis associated with POR.

**Figure 6.**
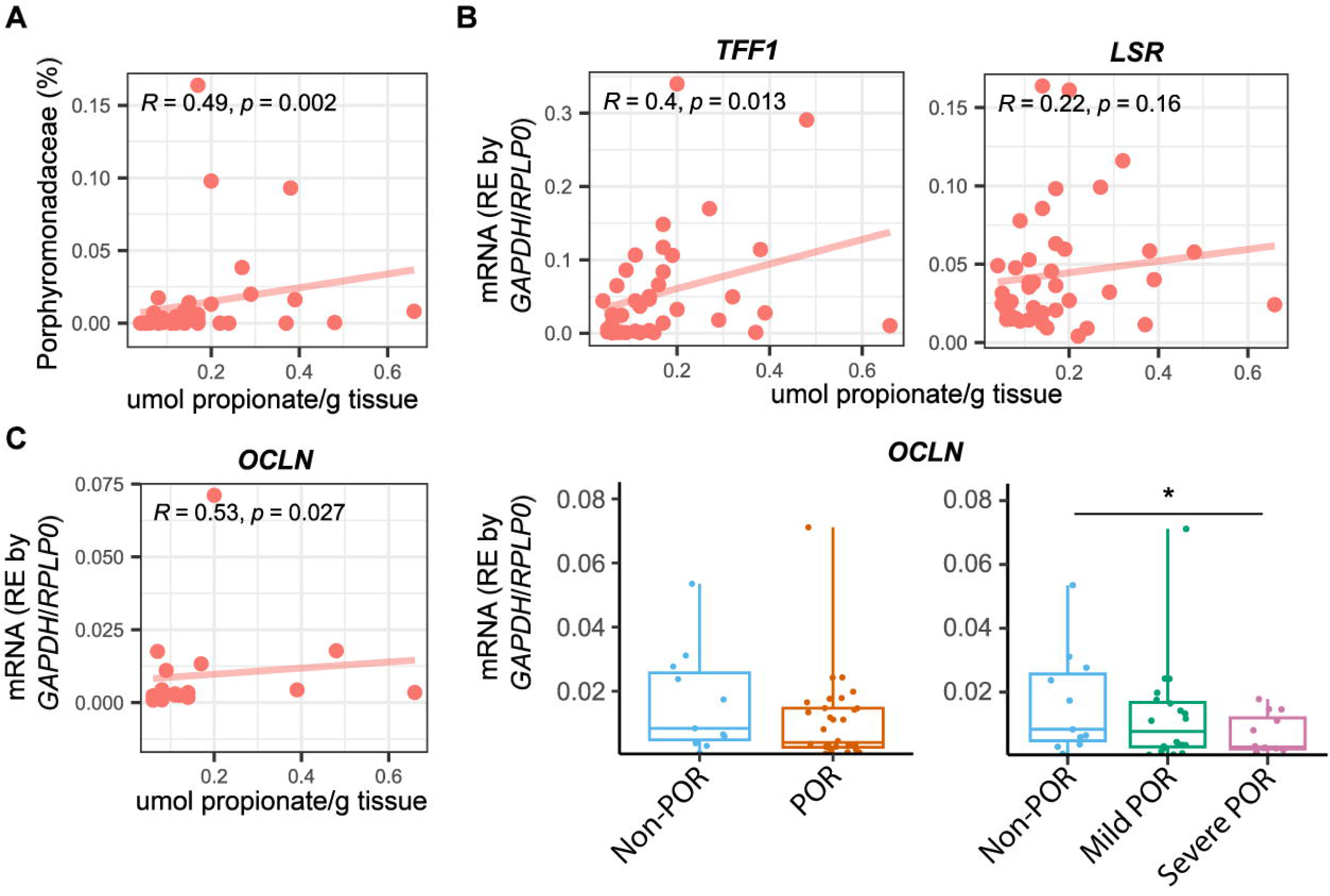
Propionate association to the Porphyromonadaceae-TFF1-barrier axis. (A) Scatter plot representing the correlation between the Porphyromonadaceae frequency in percentage (y-axis) and the propionate levels in µmol/g of tissue (x-axis). Scatter plot data are represented as points and the R^2^ and the P-value are given in the graph. Fitted line draws the y=mx+n formula. Correlation test was performed using Spearman statistical analysis. (B) Scatter plots depicting the correlation of the propionate levels (x-axis) with the mRNA expression of *TFF1* and *LSR* (y-axis) given as RE and normalized by the expression of *GAPDH* and *RPLP0* as housekeeping genes. (C) Propionate (x-axis) correlation to the *OCLN* mRNA expression (y-axis) in POR, and boxplots showing the *OCLN* expression (y-axis) depending on the POR phenotype. Boxplots contain boxes indicating IQR, and bars, indicating maximum and minimum. The median point is depicted as a bar inside the box. Asterisks indicate significant differences between groups, as determined by the Kruskal-Wallis test with post-hoc Dunn’s test. Significance levels are: * P-value ≤ 0.05. IQR, interquartile range; POR, post-operative recurrence; R, rho; RE, relative expression.

## Discussion

Microbial dysbiosis is increasingly recognized as a central contributor to POR in CD. In this study, we applied an integrative multi-omic strategy combining mucosal transcriptomics, bacteriome profiling, and metabolite analysis to explore the molecular mechanisms underlying POR. Importantly, this work was designed as a mechanistic discovery study, prioritizing biologically coherent host–microbiota interactions rather than immediate clinical prediction.

A key strength of our approach is the parallel analysis of inflamed and microscopically uninflamed mucosa. While inflamed tissue reflects active disease processes, uninflamed mucosa closely resembles the existing mucosa after surgery. Analysis of this tissue enabled the identification of transcriptional and microbial changes that precede overt inflammation and may contribute to POR susceptibility. Here, genes whose expression is relevant for the onset of POR are discovered and related to bacterial groups. PEAs further allowed functional interpretation of these alterations.

One of the first observations was a reduction in microbial alpha diversity in CD compared with IBD-free controls, which was most pronounced in patients who later developed POR. Reduced microbial diversity is a well-established feature of CD^27,28^, although similar reductions have also been reported in colorectal cancer^29^, indicating that comparisons with controls should be interpreted cautiously and primarily as contextual references rather than as a healthy baseline. Previous studies evaluating diversity at the time of surgery have yielded mixed results, with some reporting no clear differences between patients who remained in remission and those who developed POR^28^, while others observed reduced richness in patients who later relapsed^30,31^. Consistent with these reports, our data suggest that diversity metrics alone are insufficient to robustly discriminate POR risk but nonetheless reflect an underlying microbial imbalance associated with this complication.

When studying the results by bacterial groups, we observed a shift towards the Enterobacteriaceae family and specifically *E. coli* (OTU582) in severer states of POR. It is widely known that the Enterobacteriaceae family is related to the severity of CD^33^. In fact, it is not only increased in surgical specimens^14^ but it has also been related to POR. For instance, Hamilton et al, reported its association with a higher risk of POR that might be due to the increase in facultative anaerobe pathobionts, such as *E. coli*^34^. In fact, the presence of AIEC in the surgical specimen was able to predict the presence of any post-operative lesion after six months^35^ due to the capacity of this strain to translocate, invade and adhere intestinal epithelial. Nevertheless, our methodology was limited in the identification of AIEC and could only classify *E. coli* at the species level. Interestingly, the OTU582 was positively correlated to muscle development pathways in the inflamed mucosa of patients with POR, suggesting a potential link to *muscularis* hyperplasia and fibrosis, interaction already described in CD pathology^36,37^.

In regard to the Xanthomonadaceae family, mainly constituted by the *S. maltophilia* species in our samples, although not being altered among phenotypes, only in POR it correlated to neutrophil activity with the key participation of *ITGAM*. This result was not validated in the independent cohort. Nevertheless, the *ITGAM* expression was observed higher in POR and severe POR. The *S. maltophilia* is a gram-negative bacterial species present in approximately 20% of patients with CD^38^ that may cause diarrheal diseases in immune-compromised patients. Interestingly, a cell wall defective *S. maltophilia* has been suggested to be involved in the pathogenesis of IBD^39^. On the other hand, *ITGAM* is an integrin which allows interactions between leucocytes and endothelial cells, important for pro-inflammatory cell recruitment in IBD^40^. Interestingly, *ITGAM* has previously been associated with neutrophils due to its linkage with cell trafficking of immune cells during inflammation, activating cytotoxic processes. In fact, its deficiency is linked to a decreased inflammation^41^. *S. maltophilia* has been also associated to an activation of the neutrophil response and its chemotaxis^42,43^. These findings suggest that Xanthomonadaceae species may contribute to a neutrophil-drive inflammatory signature associated with POR, rather than representing a validated causal interaction. However, the most promising finding is the increase of the *ITGAM* expression, which might indicate a higher susceptibility to POR due to increased neutrophil activity.

Our data also revealed a reduction in the relative abundance of the Porphyromonadaceae family -mainly represented by *P. gordonii*- in patients who subsequently developed POR. Members of the Parabacteroides genus have previously been associated with remission in CD following surgical resection^14,44^. Porphyromonadaceae are obligate anaerobic bacteria known for their ability to produce SCFA^45^, metabolites known to regulate epithelial barrier integrity and immune tolerance, thereby limiting bacterial translocation^46,47^.

Notably, our analysis indicates a significant association between this bacterial family and epithelial barrier function, particularly in the inflamed mucosa of patients with POR. This association is supported by correlations with the expression levels of epithelial-associated proteins such as *LSR* and *TFF1*, the latter of which was validated in an independent replicate cohort. Consistently, transcriptomic PEAs revealed negative enrichment of digestion, absorption and epithelial maintenance pathways in POR. Going into detail, *LSR* is a critical component of tricellular tight junctions^49^, while *TFF1* plays a pivotal role in mucosal protection and repair by enhancing the viscosity and elasticity of the mucin layer. Due to its anti-inflammatory properties, *TFF1* has been implicated as a protective factor in IBD^50^. Importantly, Porphyromonadaceae abundance correlated with mucosal propionate levels, which in turn correlated with *TFF1* and *OCLN* expression. SCFAs have been shown to regulate the expression of tight junction proteins and mucins^47,51^, and Parabacteroides species have been demonstrated to upregulate occludin-1 expression, thereby reinforcing epithelial barrier integrity^52^. Taken together, these findings converge on a coherent Porphyromonadaceae-SCFA-epithelial barrier axis that appears compromised in patients who develop POR. This, in turn, may facilitate bacterial translocation and contribute to the pathogenesis of post-operative lesions in CD patients. Together, our data support a model in which depletion of Porphyromonadaceae leads to reduced SCFA availability, impaired epithelial barrier maintenance, and increased susceptibility to POR.

This study represents, to our knowledge, the first prospective integrative multi-omic analysis combining host transcriptomics, mucosal bacteriome profiles and SCFA measurements to investigate POR in CD. Despite its novel approach, the study has several limitations. First, the inception cohort was relatively small, limiting stratification by POR severity. However, it was primarily intended for mechanistic discovery, with validation of prioritized findings performed in an independent cohort. Another limitation is the use of samples from patients operated on for colorectal cancer as IBD-free controls. Although the non-tumoral tissue was histologically analyzed and showed no inflammatory alterations^6^, these patients may still exhibit microbial dysbiosis^29^. Importantly, key conclusions were driven within-CD comparisons. Temporal heterogeneity in patient recruitment and treatment strategies was mitigated by validation in a more recent cohort. We also identified challenges at the OTU level due to limited counts per sample, reducing interpretability and statistical power. Analyses at the bacterial family level provided clearer, more consistent results, facilitating biological interpretation. Notably, families such as Xanthomonadaceae and Porphyromonadaceae comprised multiple OTUs representing the same species, supporting the rationale for family-level analysis. This reinforces the use of higher taxonomic resolution for integrating microbial and host transcriptomic data in complex human tissues.

Overall, this work applies a stepwise, hypothesis-driven multi-omic integration framework to uncover host–microbiota interactions relevant to POR. While *ITGAM* may represent a marker of a pro-inflammatory immunity associated with POR, the most consistent and biologically coherent finding is the depletion of Porphyromonadaceae and its association with impaired epithelial barrier function through reduced SCFA availability and diminished TFF1 signaling.

These results provide a mechanistic foundation for the development of microbiome-informed prognostic strategies and support the exploration of targeted microbial or metabolite-based interventions to prevent post-operative lesions in CD.

## Data availability

Gene profiling data from the inception cohort were deposited in NCBI’s Gene GEO and are accessible through the GEO Series accession number GSE83448 (https://www.ncbi.nlm.nih.gov/geo/query/acc.cgi?acc=GSE83448). The microbiota data reported in this paper have been deposited NCBI SRA database with BioProject accession number PRJNA1241201 (http://www.ncbi.nlm.nih.gov/bioproject/1241201). The other datasets generated during and/or analyzed during the current study are available from the corresponding author on reasonable request.

## Acknowledgements

We acknowledge the patients and the IGTP BioBank integrated for its collaboration in sample collection and processing. We wish to thank the participation of Raquel Pluvinet and Helena Raurell from the High Content Genomics and Bioinformatics Unit in the IGTP for the RNA quality control and the help with metagenomic library processing. We also thank Albert Boronat Toscano, Mariona Llaves and Carla Bernal for the help in qRT-PCR experiments. We thank Adrià Aterido and Toni Julià from the Rheumatology Research Group from Vall d’Hebron Research Institute for the work on co-expression modules that was finally not used in this paper. We would also like to acknowledge the participation of Maria Pilar Armengol and Irina Pey from the IGTP Translational Genomics Unit for the 16S metagenomic sequencing analysis. We thank Mireia Coma and Cristina Segú from the Anaxomics Biotech S.L. for the transcript-bacteria correlation analyses and their guidance on the implementation of the integrative multi-omic approach. We also acknowledge the participation of Joan Carles Domingo Pedrol and Begoña Cordobilla in the SCFA experiments. We finally acknowledge Mireya Jimeno, Micaella Elizabeth Aquino and Karol Stephany Matute from Department of Pathology of HUGTP for histopathological evaluations of the Goblet cells which were eventually not used for this article.

## Author contributions

R.S., M.C., M.R. and V.L. conducted the main experiments and worked on pathway enrichment analyses and data multi-omic integration. E.D., Y.Z., G.P. and M.M. contributed to the selection of the study population, informed consent, the processing of human samples and the prospective recording of clinical and endoscopic data. E.D. and M.M. carried out the endoscopic assessments of POR, as well as the clinical evaluation. N.BH. worked on the gene profiling experiments. R.S. and M.C. did the gene profiling statistical and bioinformatic analysis. R.S., M.R., L.C., D.M. and I.G. worked on the validation experiments. M.LS. and M.MM. did the metagenomic sequencing experiments of the inception cohort and helped on implementing the sequencing of the second cohort. R.S. and M.R. worked on the sequencing experiment of the validation cohort. R.BF., J.FS. and L.S. helped on the performance of the 16S sequencing and carried out the bioinformatic and statistical analyses. J.M., C.S., L.S., E.D. and M.M. conceptualized the study. R.S., M.C., M.R. and J.M. worked on data interpretation and wrote the first draft. J.M. and C.S are guarantors of this work.

