## Supplementary Material for "Unveiling the Porphyromonadaceae-*TFF1* Interaction and *ITGAM* as Critical Factors in Post-operative Recurrence of Crohn’s Disease"

**Supplementary Methods**

***Methods S1.*** *RNA extraction, quality control, DNase treatment and cDNA synthesis.*

Total RNA extraction of all the inflamed and uninflamed intestinal mucosa was performed using the miRNeasy Mini Kit (Qiagen, Germany) as described in Yamile et al^1^. Briefly, 50 mg of tissue per sample were mixed with 700 μL of Qiazol, using GentleMacs Dissociator (Miltenyi Biotec, Germany), following the program RNA_2. RNA was isolated using phase separation, ethanol precipitation, and purification through columns. RNA was eluted in 50 μL of RNase-free water and quantified with Nanodrop. RNA integrity was evaluated using the RNA 6000 Nano Kit (Agilent Technologies, Spain) on the Agilent Bioanalyzer. Only the extractions with a RIN ≥ 6.5 were considered. Next, samples were DNAse-treated using the manufacturer’s protocol of the DNase I (RNase-free kit; Thermo Fisher Scientific, Waltham, MA). Finally, cDNA synthesis was performed using the PrimeScriptTM RT reagent (Takara, Japan) following manufacturer’s instructions.

***Methods S2.*** *Ileocolonic gene expression profiling.*

Gene expression profiling was performed as described in Yamile et al^1^. Briefly, the RNA was amplified and biotin-labeled using the MessageAmp II-Biotin enhanced kit (Ambion, Thermo FisherScientific, Waltham, MA). Samples were checked for high purity and integrity using Experion system (Bio-Rad, Hercules, CA) before hybridization them for microarray analysis. Whole transcriptome was conducted using the Human Whole Genome Microarray (Applied Microarrays CodeLink, Tempe, AZ) in the TrayMix Hybridization Station (Arrayit, Sunnyvale, CA), and the fluorescence signal detection by InnoSan 770 scanner (Innopsys, France). Probe annotation was updated with NCBI RefSeq database^2^, identifying 20,902 unique probes from 53,485 transcripts included in microarrays. Normalized and corrected data were deposited in NCBI’s Gene Expression Omnibus (GEO)^3,4^ under accession number GSE83448 in compliance with Minimum Information About a Microarray Experiment (MIAME) standards^1,5,6^.

***Methods S3.*** *Mucosa adhered-bacteriome sequencing.*

Total DNA was extracted from 25 mg of patient’s intestinal tissue sample of both inception and validation cohorts, and the controls using the Nucleospin® tissue kit (Machery-Nagel). Samples were lysed with lysozyme and proteinase K, followed by RNase A treatment. DNA was purified through column binding and ethanol precipitation. Elution was performed in 100 μL of DNase/RNase-free water. DNA was quantified with NanoDrop and integrity verified by 0.8% agarose gel electrophoresis.

Bacterial 16S rDNA hypervariable region V4 sequencing was performed using a modified version of Illumina’s 16S Metagenomic Sequencing Library Preparation protocol^7^. Dual index primers were used for single-step PCR amplification. Amplicon quality was assessed with the Agilent D1000 ScreenTape System (Agilent Technologies, Barcelona, Spain). Two different bands were amplified around 300 and 380 bp for mitochondrial MT-RNR2 gene and bacterial 16S rDNA, respectively. Only, the 380bp band was purified by low-melting 2% agarose gel extraction and cleaned using the Nucleospin gel and PCR clean-up Mini-kit (Macherey-Nagel, Germany). Samples were pooled equimolarly, concentrated, and sequenced on the Illumina MiSeq System with 8 pM loading concentration and 15% PhiX control. Run parameters included: cluster density 700-800k/mm^2^; >85% clusters passing filter; 15% aligned (amount of PhiX); no spikes in corrected intensity plot; all indices identified following index reads; and final >Q30 score of >70%.

***Methods S4.*** *Bioinformatic metagenomic analysis.*

Sequencing data from sample batches were proposed independently and then jointly analyzed. Batch effects were assessed using the MBECS package in R^8^, and Principal Component Analysis (PCA) using Ampvis2 package. Both analyses indicated that no significant batch effects were present in the data, allowing unified analysis.

Raw sequences were trimmed with TrimGalore and quality-checked using MultiQC. Data were imported into QIIME 2 for processing following standard workflows^9^. Quality control was performed using DADA2 and then, a feature table was generated to quantify the unique sequence variants, often referred to as operational taxonomic units (OTUs), across samples. Taxonomic assignment was performed using a pre-trained classifier on the SILVA 138-99 database.

Diversity analyses included both alpha (Shannon & Simpson indices) and beta (Bray-Curtis, Jaccard) diversity, visualized via Principal Coordinates Analysis (PCoA) to identify patterns and clustering of samples that may correlate with specific clinical factors or environmental variables. To rule out potential technical biases, we performed a correlation analysis between sequencing depth (raw read pairs) and observed species richness (Figure S1). Although a weak positive correlation was observed (R² = 0.1), the limited explanatory power indicates that sequencing depth alone does not account for diversity differences, which are likely influenced by biological and clinical variables.

Further analysis was conducted using Phyloseq and Ampvis2 in R for microbial community profiling and visualization. Differential abundance analysis between groups was carried out using DESeq2, applying a cutoff of adjusted *p* ≤ 0.05 and |log2FC| ≥ 1.5, base mean > 30. Unclassified bacteria at family level were included in the calculations but not in heatmaps.

***Methods S5.*** *qRT-PCR for of transcripts in the validation cohort.*

PCR thermal cycling included initial denaturing at 95 °C for 10 s, 40 cycles of 95 °C for 15 s, and 60 °C for one minute (LightCycler480 system; Roche Diagnostics). 2^−ΔCt^ method was used^10^ in the R software 4.2.2 version. Results were normalized with both *GAPDH* and *RPLP0* housekeeping genes. Data from the inception cohort was correlated to the values of its correspondent transcripts from the gene profiling analysis in order to validate Taqman primers and probes (data not shown).

**Supplementary Figures**

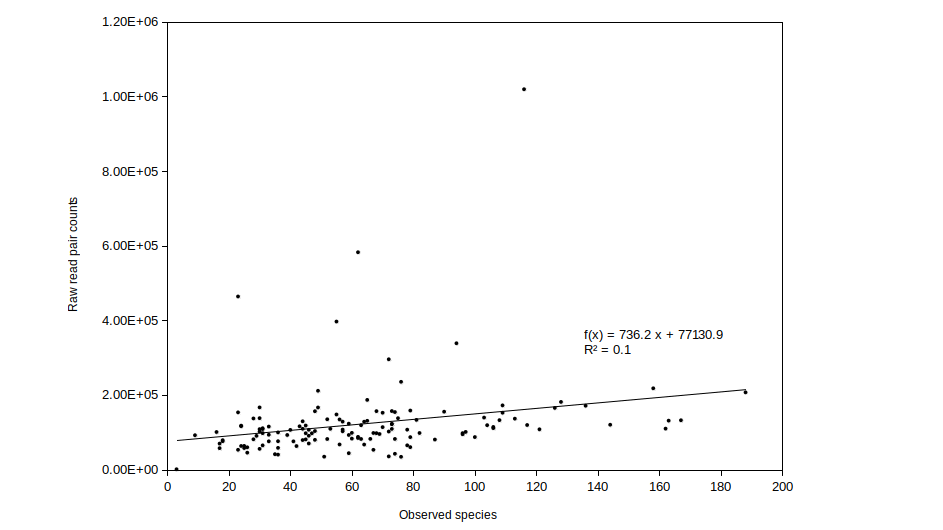

***Figure S1.*** *Scatter plot depicting the relationship between the number of observed species and the raw read pair counts across all samples.* The x-axis indicates the number of observed species (species richness) and the y-axis represents the total raw read pair counts obtained from sequencing. The fitted linear regression line illustrates the correlation between both variables.

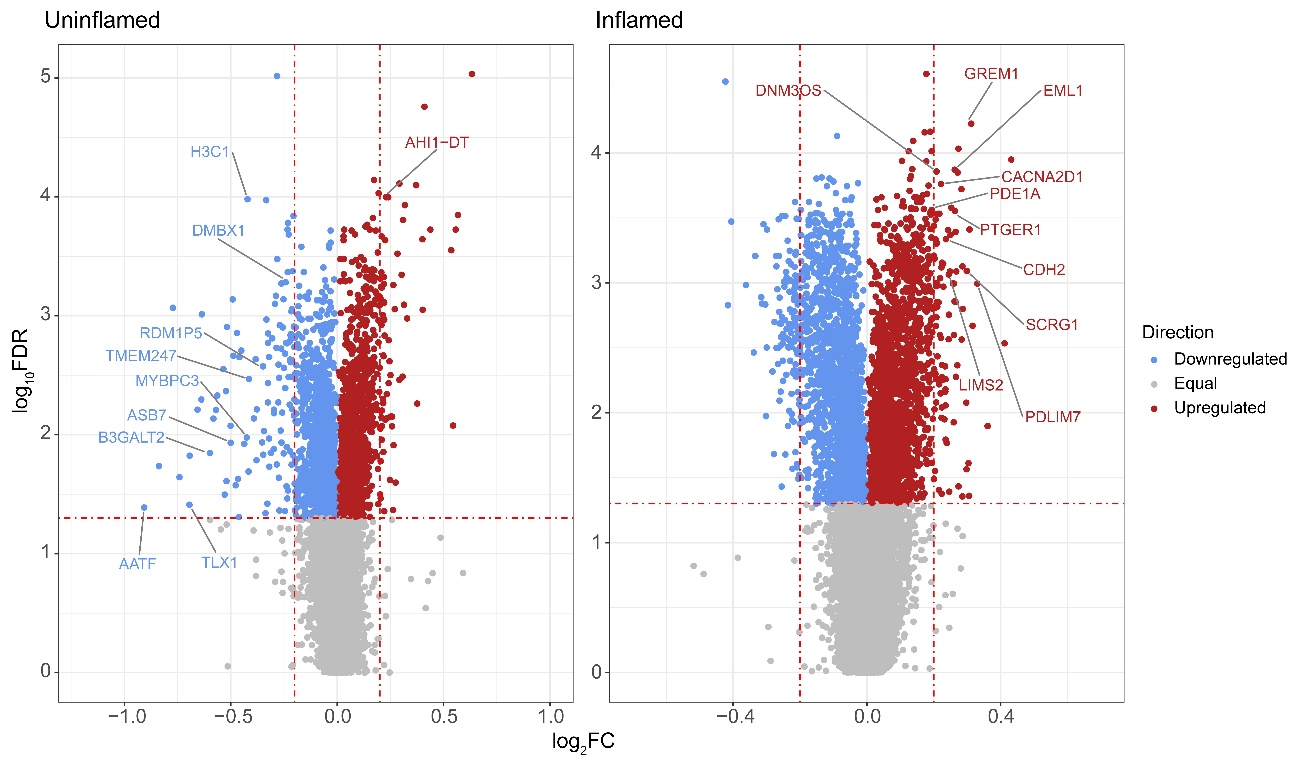

***Figure S2.*** *Volcano plots depicting the top 10 differentially expressed transcripts in the non-POR compared to the IBD-free controls in both inflammatory statuses.* The y-axis represents the log_10_FDR while the x-axis depicts the log_2_FC. Red dotted lines mark the significance thresholds (FDR ≤ 0.05 and |FC| ≥ 0.2). Red dots are for downregulated while red ones are for upregulated. Only the top 10 scoring transcripts for each condition are labeled with their gene symbol. FC, fold-change; FDR, false discovery rate; POR, post-operative recurrence.

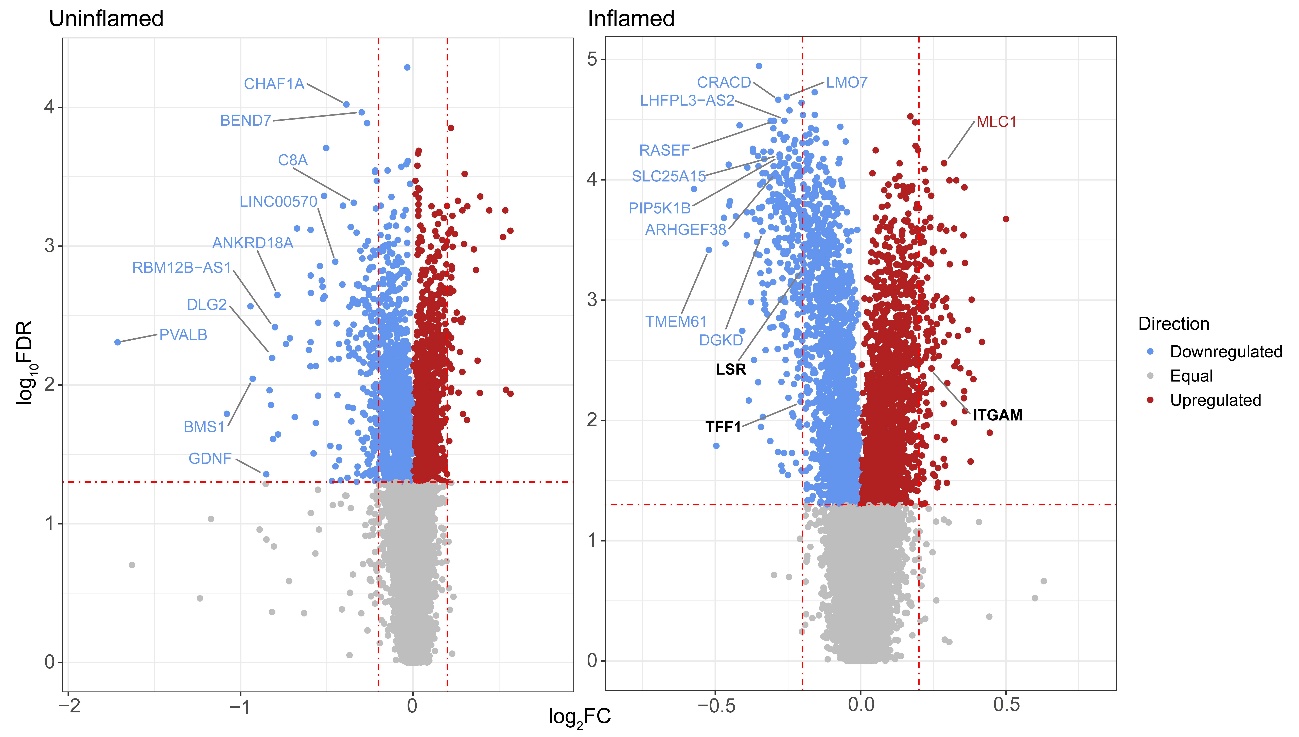

***Figure S3.*** *Volcano plots depicting the top 10 differentially expressed transcripts and the important genes for the bacteriome-transcriptome integrative analysis in POR compared to the IBD-free controls in both inflammatory statuses.* The y-axis represents the log_10_FDR while the x-axis depicts the log_2_FC. Red dotted lines mark the significance thresholds (FDR ≤ 0.05 and |FC| ≥ 0.2). Red dots are for downregulated while red ones are for upregulated. The top 10 scoring transcripts for each condition are labeled with their gene symbol. Genes important for the integration analysis are marked in bold. FC, fold-change; FDR, false discovery rate; POR, post-operative recurrence.

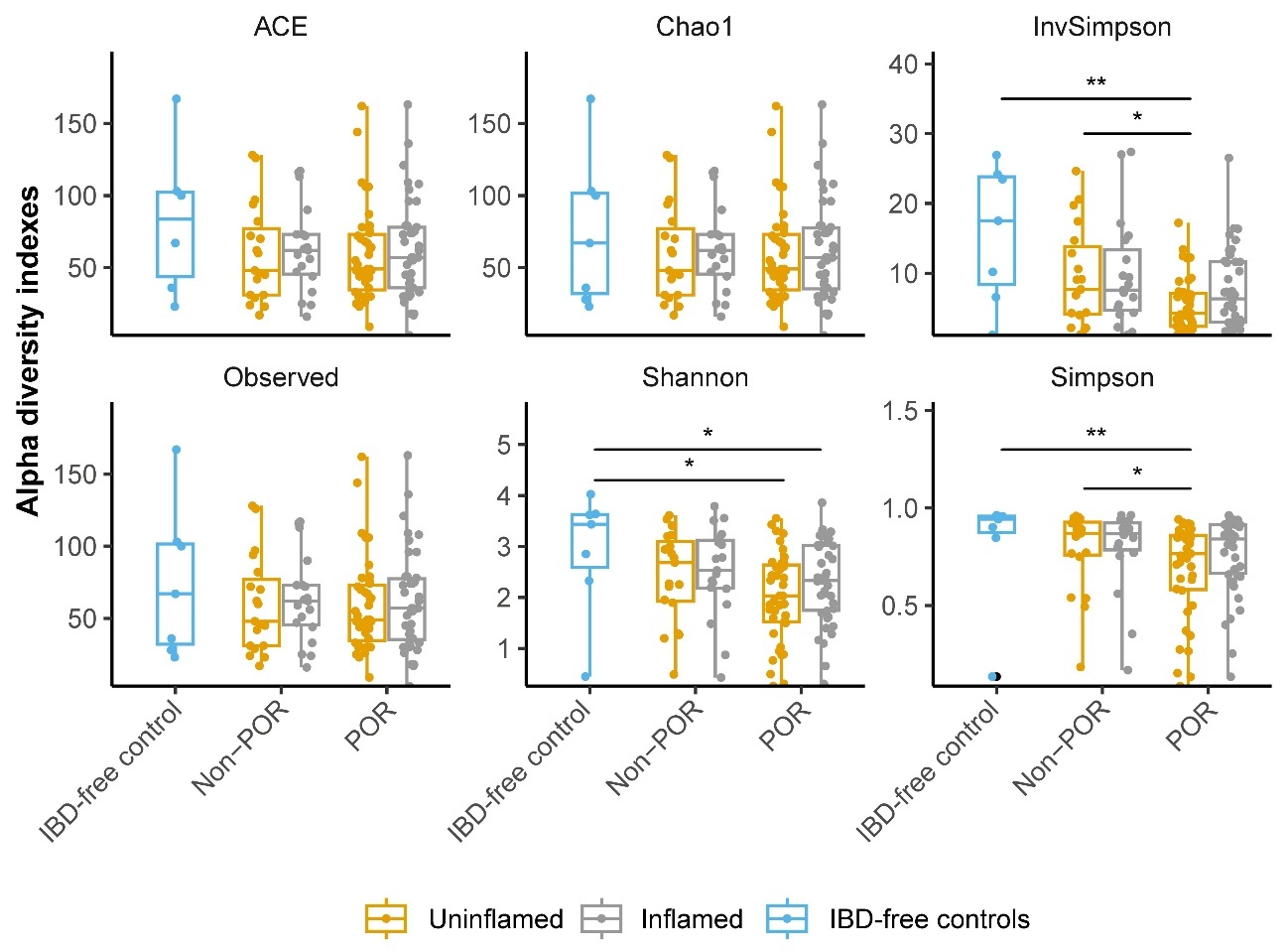

***Figure S4.*** *Boxplots depicting the alpha diversity metrics between the IBD-free controls and POR groups.* The x-axis represents the phenotypes, and the y-axis illustrates each alpha diversity metric. Data are represented as points summarized in boxplots, indicating IQR, and bars, indicating maximum and minimum. The median point is depicted as a bar inside the box. Treatment groups are differentiated using colors. Asterisks indicate significant differences between groups, as determined by the Kruskal-Wallis test with post-hoc Dunn’s test. Significance levels are: * P-value ≤ 0.05; ** P-value ≤ 0.01. ACE, abundance-based coverage estimator; IBD, inflammatory bowel disease; IQR, interquartile range; POR, post-operative recurrence.

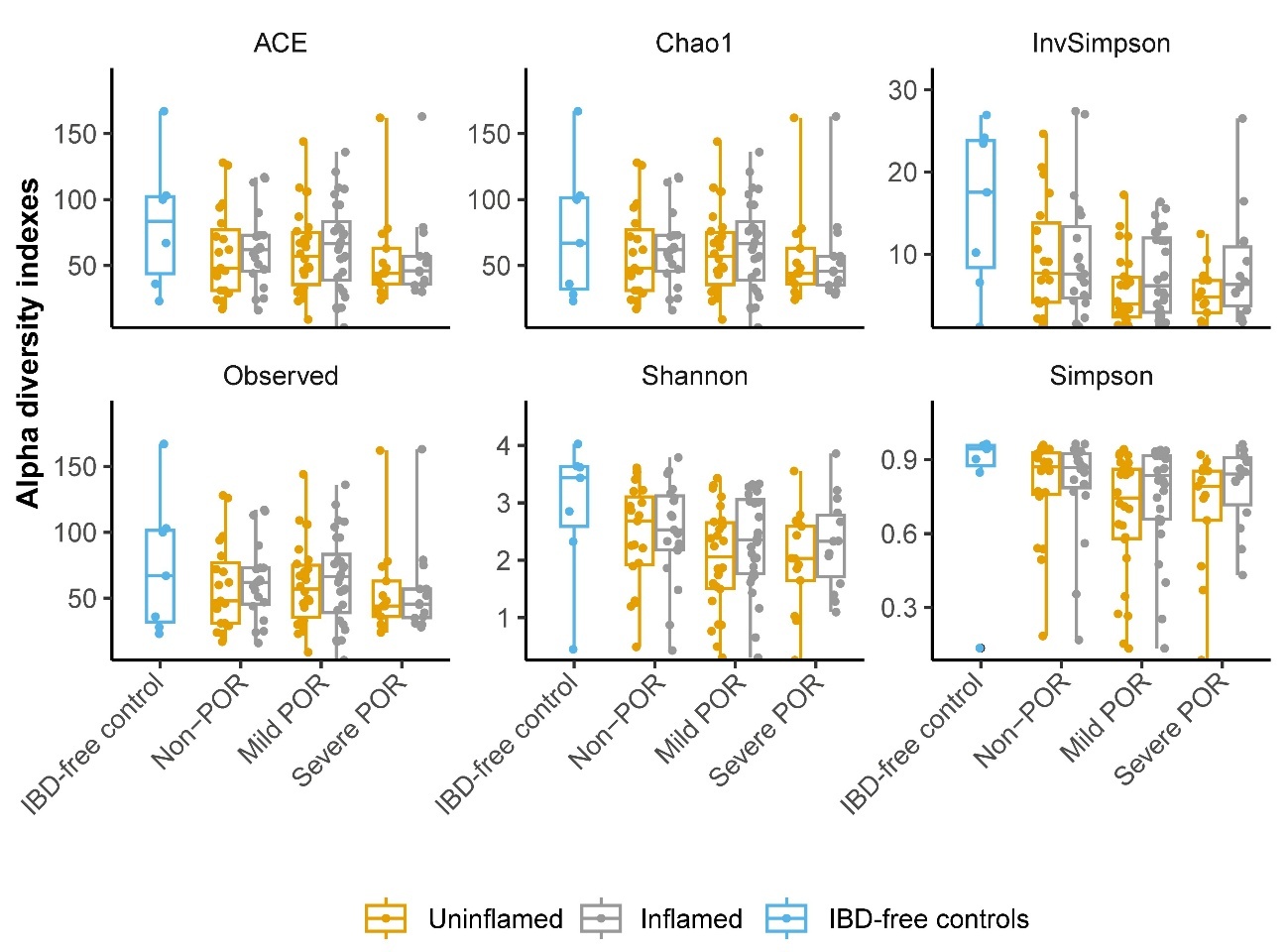

***Figure S5.*** *Boxplots depicting the alpha diversity metrics between POR groups and IBD-free controls stratifying POR in mild and severe.* The x-axis represents the phenotypes, and the y-axis illustrates each alpha diversity metric. Data are represented as points summarized in boxplots, indicating IQR, and bars, indicating maximum and minimum. The median point is depicted as a bar inside the box. Treatment groups are differentiated using colors. ACE, abundance-based coverage estimator; IBD, inflammatory bowel disease; IQR, interquartile range; POR, post-operative recurrence.

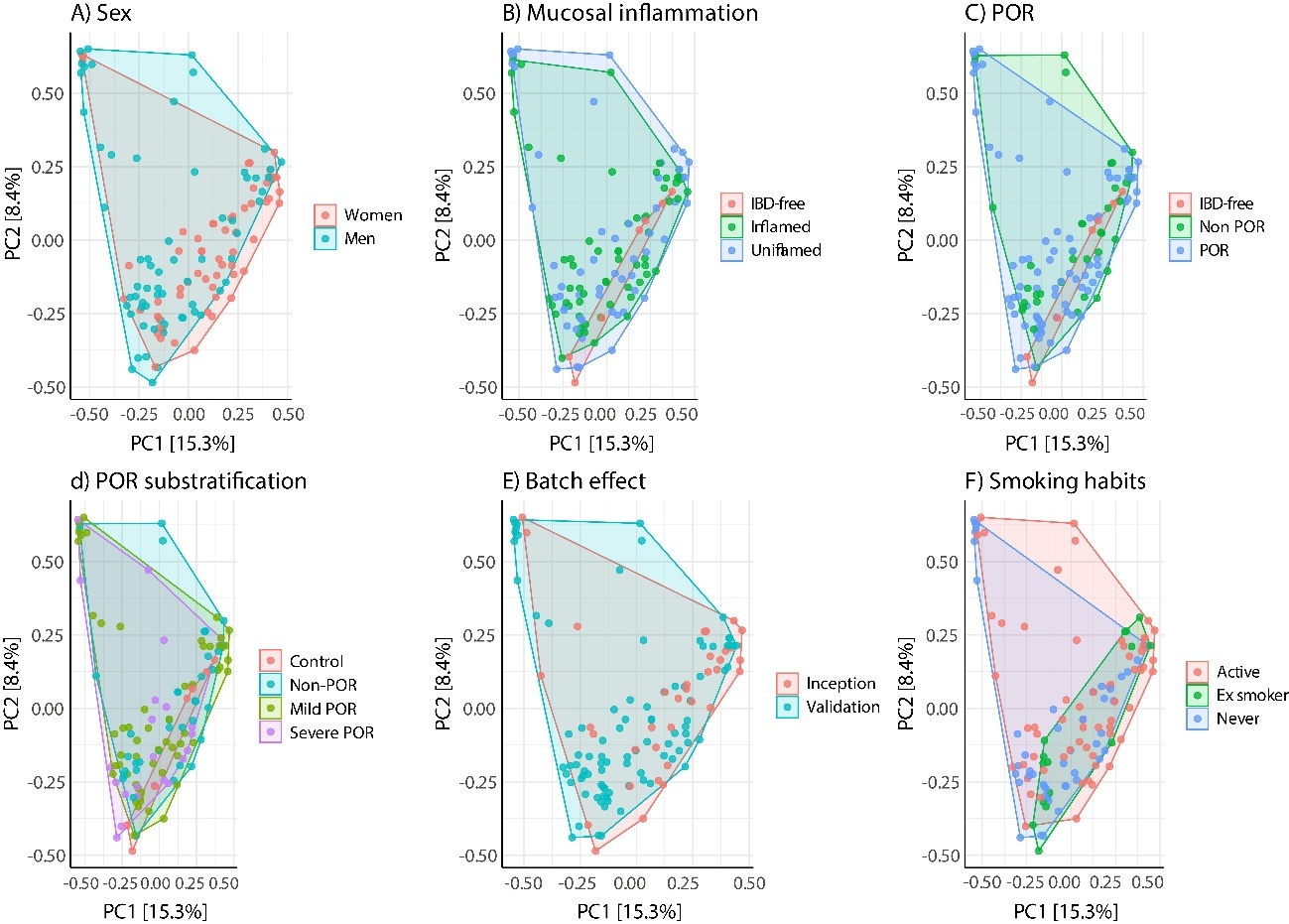
***Figure S6.*** *PCA plots of microbial community composition across different sample variables.* The variable corresponds to (a) sex, (b) mucosal inflammation, (c) POR status, (d) POR sub-stratification, (e) batch and (f) smoking habits. The first two PC, PC1 and PC2, explain 15.3% and 8.4% of the variance, respectively. Each point represents a sample, with clustering indicating similarity in microbial community composition based on the associated variable. PC, principal component; PCA, PC analysis; POR, post-operative recurrence.

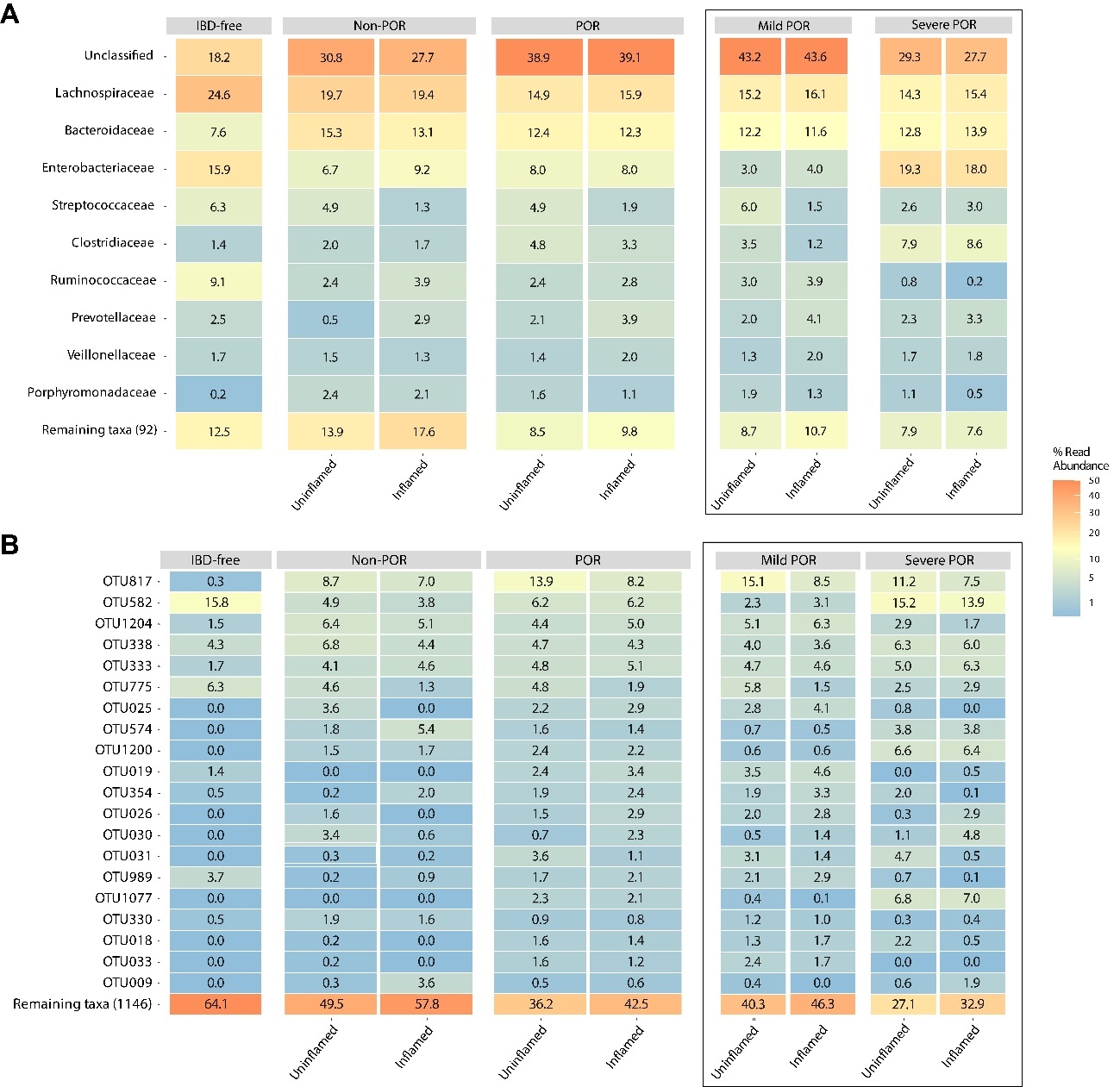
**Figure S7*.*** *Alterations in bacterial family and OTU level abundance associated with POR in CD (with unclassified bacteria).* (A) Heatmap showing the relative abundances (%) of the top 10 most prevalent bacterial families and (B) the top 20 most prevalent OTUs across all samples, stratified by POR and mucosal inflammation status. CD, Crohn’s disease; FC; fold-change; GO, gene ontology; OTU, operational taxonomic unit; ORA, over-representation analysis; PEA, pathway enrichment analysis; POR, post-operative recurrence.

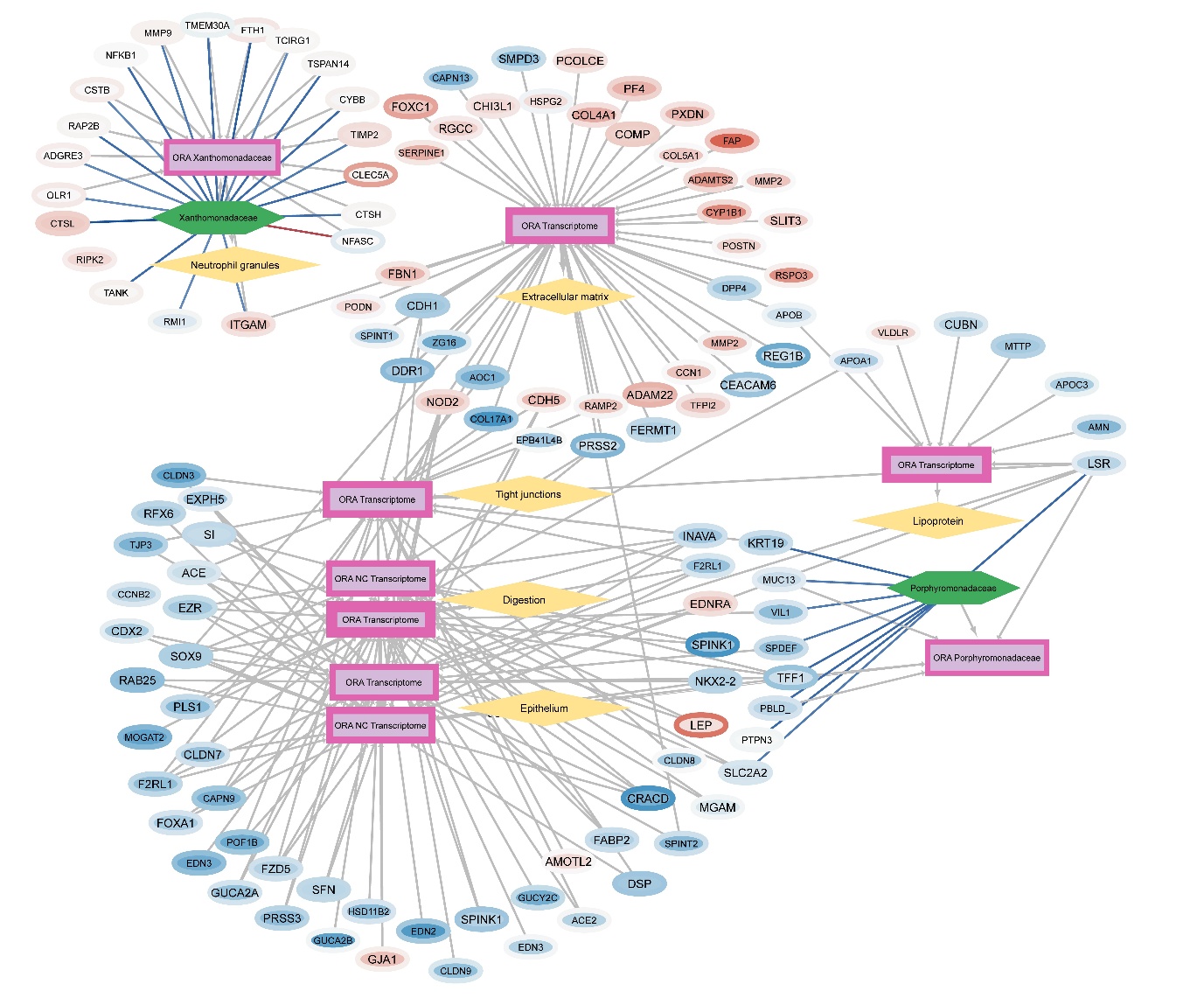

**Figure S8*.*** *Whole integrative transcriptome/microbiome network of the inflamed mucosa in patients with POR.* (A) Graphical representation of the transcriptome/microbiome integration by Cytoscape. Gene symbols are given for each transcript and intensity of the color (red for under-expressed, blue for over-expressed) is a value calculated using both the FC and the FDR. Inner color refers to the comparison POR vs IBD-free controls, and border color refers to the comparison between POR vs non-POR. Font size is bigger for non-coincident transcripts. PEA aggrupation nodes border width corresponds to the highest -log_2_(q-value) of each aggrupation. Link line color between transcripts and bacteria indicate the intensity of the correlation (red for negative correlation, blue for positive). Whole representation can be found in <https://doi.org/10.18119/N9731C>. FC; fold-change; FDR, false discovery rate; GO, gene ontology; ORA, over-representation analysis; PEA, pathway enrichment analysis; POR, post-operative recurrence.

**Supplementary tables**

***Table S1.*** *TaqMan®Assays used in the qRT-PCR experiments.*

| **Gene symbol** | **Taqman® Assay** |
| --- | --- |
| *ITGAM* | Hs00167304_m1 |
| *LSR* | Hs00210880_m1 |
| *TFF1* | Hs00907239_m1 |
| *CLND1* | Hs00221623_m1 |
| *CLND2* | Hs01549234_m1 |
| *MUC2* | Hs00159374_m1 |
| *TJP1* | Hs01551861_m1 |
| *OCLN* | Hs01049883_m1 |
| *GAPDH* | Hs02758991_g1 |
| *RPLP0* | Hs99999902_m1 |

***Table S2.*** *Demographic and epidemiological features of the inception cohort in the POR stratification.*

|  | **Non-POR** | **POR** | **P-Value** |
| --- | --- | --- | --- |
| **n** | 12 | 8 |  |
| **Age, years** | 31 (26-44) | 36 (34-38) | ns |
| **Female sex; n (%)** | 6 (50) | 3 (38) | ns |
| **BMI** | 22 (19-23) | 20 (18-23) | ns |
| **Active smoking; n (%)** |  |  |  |
| Never | 3 (25) | 0 | ns |
| Ex-smoker | 0 | 0 |  |
| Active | 9 (75) | 8 (100) |  |
| **CD location; n (%)** |  |  |  |
| Ileal | 7 (58) | 7 (88) | ns |
| Colic | 0 | 0 |  |
| Ileocolic | 5 (42) | 1 (13) |  |
| **Perianal disease; n (%)** | 3 (25) | 2 (25) | ns |
| **Family background; n (%)** | 5 (41) | 0 | ns |
| **Time from CD diagnosis to surgery, months** | 51 (32-94) | 77 (26-99) | ns |
| **Treatment before surgery; n (%)** |  |  |  |
| Nothing or aminosalicylates | 2 (17) | 1 (13) | ns |
| Immunomodulators | 7 (58) | 4 (50) |  |
| Anti-TNF-α | 0 | 1 (12.5) |  |
| Anti-TNF-α + immunomodulators | 3 (25) | 2 (25) |  |
| Undetermined |  |  |  |
| **Surgical indication; n (%)** |  |  |  |
| Stricturing disease | 8 (67) | 3 (38) | ns |
| Penetrating disease | 4 (33) | 5 (63) |  |
| Inflammatory phenotype | 0 | 0 |  |

Data is given as median (IQR) if not indicated. BMI, body mass index; CD, Crohn’s disease; IQR, inter-quartile range; ns, non-significant; POR, post-operative recurrence.

***Table S3.*** *Demographic and epidemiological features of the validation cohort in the POR stratification.*

|  | **Non-POR** | **POR** | **P-Value** |
| --- | --- | --- | --- |
| **n** | 12 | 37 |  |
| **Age, years** | 35 (31-44) | 36 (24-49) | ns |
| **Female sex; n (%)** | 6 (50) | 15 (41) | ns |
| **BMI** | 22 (19–23) | 23 (19–25) | ns |
| **Active smoking; n (%)** |  |  |  |
| Never | 6 (50) | 18 (49) | ns |
| Ex-smoker | 2 (17) | 5 (13) |  |
| Active | 4 (33) | 14 (38) |  |
| **CD location; n (%)** |  |  |  |
| Ileal | 8 (67) | 22 (60) | ns |
| Colic | 0 | 1 (3) |  |
| Ileocolic | 4 (33) | 14 (38) |  |
| **Perianal disease; n (%)** | 3 (25) | 14 (38) | ns |
| **Family background; n (%)** | 3 (25) | 5 (14) | ns |
| **Time from CD diagnosis to surgery, months** | 85 (23–142) | 57 (21–92) | ns |
| **Treatment before surgery; n (%)** |  |  |  |
| Nothing or aminosalicylates | 2 (17) | 8 (22) | ns |
| Immunomodulators | 5 (42) | 11 (30) |  |
| Anti-TNF-α | 2 (17) | 5 (14) |  |
| Anti-TNF-α + immunomodulators | 3 (25) | 1 (27) |  |
| Undetermined | 0 | 3 (8) |  |
| **Surgical indication; n (%)** |  |  |  |
| Stricturing disease | 6 (50) | 15 (41) | ns |
| Penetrating disease | 6 (50) | 18 (49) |  |
| Inflammatory phenotype | 0 | 3 (8) |  |
| Undetermined | 0 | 1 (3) |  |

Data is given as median (IQR) if not indicated. BMI, body mass index; CD, Crohn’s disease; IQR, inter-quartile range; ns, non-significant; POR, post-operative recurrence.

***Table S4.*** *Demographic and epidemiological features of the validation cohort in the POR substratification.*

|  | **Non-POR^[1]^** | **Mild POR^[2]^** | **[1] vs [2] (P-Value)** | **Severe POR^[3]^** | **[1] vs [3] (P-Value)** | **[2] vs [3] (P-Value)** |
| --- | --- | --- | --- | --- | --- | --- |
| **n** | 12 | 25 |  | 12 |  |  |
| **Age, years** | 35 (31-44) | 36 (24-50) | ns | 36 (26-43) | ns | ns |
| **Female sex; n (%)** | 6 (50) | 10 (40) | ns | 5 (42) | ns | ns |
| **BMI** | 26 (23–28) | 24 (19–25) | ns | 19 (18–20) | ns | ns |
| **Active smoking; n (%)** |  |  |  |  |  |  |
| Never | 6 (50) | 13 (52) | ns | 5 (42) | ns | ns |
| Ex-smoker | 2 (17) | 5 (20) |  | 0 |  |  |
| Active | 4 (33) | 7 (28) |  | 7 (58) |  |  |
| **CD location; n (%)** |  |  |  |  |  |  |
| Ileal | 8 (67) | 14 (56) | ns | 8 (67) | ns | ns |
| Colic | 0 | 1 (4) |  | 0 |  |  |
| Ileocolic | 4 (33) | 10 (40) |  | 4 (33) |  |  |
| **Perianal disease; n (%)** | 3 (25) | 10 (40) | ns | 4 (33) | ns | ns |
| **Family background; n (%)** | 3 (25) | 3 (12) | ns | 2 (17) | ns | ns |
| **Time from CD diagnosis to surgery, months** | 85 (23–142) | 63 (41–119) | ns | 37 (5–89) | ns | ns |
| **Treatment before surgery; n (%)** |  |  |  |  |  |  |
| Nothing or aminosalicylates | 2 (17) | 4 (16) | ns | 4 (33) | ns | ≤0.05 |
| Immunomodulators | 5 (42) | 9 (36) |  | 2 (17) |  |  |
| Anti-TNF-α | 2 (17) | 2 (8) |  | 3 (25) |  |  |
| Anti-TNF-α + immunomodulators | 3 (25) | 7 (28) |  | 3 (25) |  |  |
| Undetermined | 0 | 3 (12) |  | 0 |  |  |
| **Surgical indication; n (%)** |  |  |  |  |  |  |
| Stricturing disease | 6 (50) | 11 (44) | ns | 4 (33) | ns | ns |
| Penetrating disease | 6 (50) | 10 (40) |  | 8 (67) |  |  |
| Inflammatory phenotype | 0 | 3 (12) |  | 0 |  |  |
| Undetermined | 0 | 1 (4) |  | 0 |  |  |

Data is given as median (IQR) if not indicated. BMI, body mass index; CD, Crohn’s disease; IQR, inter-quartile range; ns, non-significant; POR, post-operative recurrence.

***Table S5.*** *Differentially expressed transcripts from the non-POR vs IBD-free controls comparison.*

|  | **GEO** | **Refseq/GenBank** | **Symbol** | **FDR** | **log_2_FC** |
| --- | --- | --- | --- | --- | --- |
| **Uninflamed** | GE811888 | BC030770 | *MYOCD-AS1* | 0.043 | 0.26 |
|  | GE55019 | NM_005451 | *PDLIM7* | 0.007 | 0.26 |
|  | GE53986 | NM_000095 | *COMP* | 0.007 | 0.25 |
|  | GE55251 | NM_007281 | *SCRG1* | 0.024 | 0.25 |
|  | GE60216 | NM_000474 | *TWIST1* | 0.002 | 0.25 |
|  | GE81510 | NM_005442 | *EOMES* | 0.002 | 0.24 |
|  | GE80070 | NM_001736 | *C5AR1* | 0.004 | 0.24 |
|  | GE506809 | BC040979 | *AHI1-DT* | 1.01x10^-04^ | 0.23 |
|  | GE60482 | NM_003383 | *VLDLR* | 0.007 | 0.23 |
|  | GE58749 | NM_017436 | *A4GALT* | 0.044 | 0.22 |
|  | GE58065 | NM_001608 | *ACADL* | 0.001 | 0.22 |
|  | GE59872 | NM_001957 | *EDNRA* | 0.001 | 0.22 |
|  | GE59684 | NM_000163 | *GHR* | 0.001 | 0.22 |
|  | GE55370 | NM_017680 | *ASPN* | 0.002 | 0.22 |
|  | GE79355 | NM_144617 | *HSPB6* | 0.014 | 0.22 |
|  | GE79242 | NM_001442 | *FABP4* | 0.006 | 0.21 |
|  | GE80040 | NM_013343 | *LINC00312* | 0.001 | 0.21 |
|  | GE518683 | NM_002427 | *MMP13* | 0.011 | 0.21 |
|  | GE57918 | NM_002736 | *PRKAR2B* | 2.15x10^-04^ | 0.21 |
|  | GE79622 | NM_012098 | *ANGPTL2* | 0.001 | 0.21 |
|  | GE80890 | NM_000093 | *COL5A1* | 0.001 | 0.20 |
|  | GE59850 | NM_001185 | *AZGP1* | 0.034 | 0.20 |
|  | GE80907 | BC008431 | *SGCD* | 0.017 | 0.20 |
|  | GE57343 | NM_006867 | *RBPMS* | 0.009 | 0.20 |
|  | GE86041 | NM_001001971 | *FAM13C* | 4.87x10^-04^ | 0.20 |
|  | GE865339 | NM_203411 | *TMEM88* | 0.032 | 0.20 |
|  | GE79347 | NM_002275 | *KRT15* | 0.002 | -0.20 |
|  | GE54360 | NM_003657 | *BCAS1* | 0.015 | -0.21 |
|  | GE657769 | BX098030 | *XG* | 4.22x10^-04^ | -0.21 |
|  | GE471595 | NM_178552 | *TEX33* | 0.001 | -0.21 |
|  | GE55306 | NM_017589 | *BTG4* | 0.002 | -0.22 |
|  | GE471268 | AL080161 | *LOC101927446* | 0.001 | -0.22 |
|  | GE55402 | NM_017711 | *GDPD2* | 0.002 | -0.22 |
|  | GE581319 | AI334015 | *ABCB5* | 0.029 | -0.23 |
|  | GE521362 | NM_172225 | *DMBX1* | 0.001 | -0.24 |
|  | GE56076 | NM_004415 | *DSP* | 0.014 | -0.25 |
|  | GE54985 | NM_019599 | *TAS2R1* | 0.001 | -0.27 |
|  | GE486855 | BC047055 | *LOC102606466* | 0.023 | -0.27 |
|  | GE605592 | CD359799 | *ANKRD50* | 0.004 | -0.33 |
|  | GE63019 | NM_007148 | *RNF112* | 0.046 | -0.34 |
|  | GE471788 | AK095115 | *RDM1P5* | 0.003 | -0.35 |
|  | GE477634 | BC038426 | *LOC101928009* | 0.012 | -0.35 |
|  | GE80202 | NM_014790 | *JAKMIP2* | 0.016 | -0.38 |
|  | GE897729 | AA431175 | *TMEM247* | 0.003 | -0.41 |
|  | GE61638 | NM_003529 | *H3C1* | 1.05x10^-04^ | -0.42 |
|  | GE62642 | NM_000256 | *MYBPC3* | 0.011 | -0.42 |
|  | GE82906 | NM_024708 | *ASB7* | 0.012 | -0.50 |
|  | GE62637 | NM_003783 | *B3GALT2* | 0.014 | -0.60 |
|  | GE545331 | NM_005521 | *TLX1* | 0.039 | -0.69 |
|  | GE55294 | NM_012138 | *AATF* | 0.041 | -0.91 |
| **Inflamed** | GE55019 | NM_005451 | *PDLIM7* | 0.001 | 0.33 |
|  | GE79962 | AF154054 | *GREM1* | 5.96x10^-05^ | 0.31 |
|  | GE60027 | NM_000867 | *HTR2B* | 0.024 | 0.30 |
|  | GE55251 | NM_007281 | *SCRG1* | 0.001 | 0.30 |
|  | GE79421 | NM_000955 | *PTGER1* | 2.80x10^-04^ | 0.26 |
|  | GE59602 | NM_004434 | *EML1* | 1.35x10^-04^ | 0.26 |
|  | GE55370 | NM_017680 | *ASPN* | 0.001 | 0.26 |
|  | GE475956 | NM_017980 | *LIMS2* | 0.001 | 0.24 |
|  | GE62436 | NM_001792 | *CDH2* | 4.61x10^-04^ | 0.24 |
|  | GE81059 | NM_001704 | *ADGRB3* | 0.007 | 0.23 |
|  | GE59898 | NM_001379 | *DNMT1* | 0.007 | 0.23 |
|  | GE62584 | NM_201432 | *GAS7* | 0.001 | 0.23 |
|  | GE82453 | NM_018934 | *PCDHB14* | 0.005 | 0.22 |
|  | GE53881 | NM_004801 | *NRXN1* | 0.004 | 0.22 |
|  | GE620213 | AK092048 | *CACNA2D1* | 1.73x10^-04^ | 0.22 |
|  | GE80221 | NM_181506 | *LRRC70* | 0.001 | 0.22 |
|  | GE79163 | NM_020239 | *CDC42SE1* | 0.003 | 0.22 |
|  | GE56445 | NM_005019 | *PDE1A* | 2.94x10^-04^ | 0.22 |
|  | GE55823 | NM_018176 | *LGI2* | 0.001 | 0.21 |
|  | GE806856 | BC042431 | *SHC3* | 0.004 | 0.21 |
|  | GE62417 | NM_031935 | *HMCN1* | 2.95x10^-04^ | 0.21 |
|  | GE83133 | NM_003617 | *RGS5* | 0.001 | 0.21 |
|  | GE56749 | NM_194303 | *JAKMIP3* | 0.001 | 0.21 |
|  | GE890536 | AK021543 | *DNM3OS* | 1.39x10^-04^ | 0.21 |
|  | GE80354 | NM_139284 | *LGI4* | 3.26x10^-04^ | 0.21 |
|  | GE82569 | NM_020957 | *PCDHB16* | 0.002 | 0.20 |
|  | GE53842 | AB033060 | *PDCD6-AHRR* | 0.009 | 0.20 |
|  | GE62312 | NM_000963 | *PTGS2* | 0.005 | 0.20 |
|  | GE514839 | NM_000856 | *GUCY1A1* | 0.001 | 0.20 |
|  | GE57547 | NM_000355 | *TCN2* | 0.001 | -0.21 |
|  | GE57561 | NM_002591 | *PCK1* | 0.004 | -0.21 |
|  | GE79410 | NM_194284 | *CLDN23* | 0.003 | -0.21 |
|  | GE60179 | NM_005257 | *GATA6* | 0.001 | -0.22 |
|  | GE58450 | NM_007289 | *MME* | 0.027 | -0.22 |
|  | GE61410 | NM_020406 | *CD177* | 0.013 | -0.24 |
|  | GE766119 | NM_031895 | *CACNG8* | 0.037 | -0.25 |

FC, fold change; FDR, false discovery rate; GEO, gene expression omnibus; IBD, inflammatory bowel disease; POR, post-operative recurrence.

***Table S6.*** *Differentially expressed transcripts from the POR vs IBD-free controls comparison.*

|  | **GEO** | **Refseq/GenBank** | **Symbol** | **FDR** | **log_2_FC** |
| --- | --- | --- | --- | --- | --- |
| **Uninflamed** | GE477882 | AL832235 | *CENPW* | 0.004 | 0.28 |
|  | GE79972 | NM_174950 | *FGF7P6* | 0.001 | 0.24 |
|  | GE663759 | AK098543 | *PDE10A* | 0.002 | 0.22 |
|  | GE88768 | NM_018124 | *RFWD3* | 0.001 | 0.22 |
|  | GE62064 | NM_004126 | *GNG11* | 0.002 | 0.22 |
|  | GE57073 | NM_006475 | *POSTN* | 0.007 | 0.22 |
|  | GE60207 | NM_001035 | *RYR2* | 0.007 | 0.21 |
|  | GE562001 | AK129955 | *CCDC178* | 0.004 | 0.21 |
|  | GE57493 | NM_000632 | *ITGAM* | 0.015 | 0.20 |
|  | GE62291 | NM_001307 | *CLDN7* | 0.006 | -0.20 |
|  | GE537535 | BX103495 | *CDH23-AS1* | 0.004 | -0.20 |
|  | GE58780 | NM_153742 | *CTH* | 0.026 | -0.20 |
|  | GE762376 | BC029465 | *MGC32805* | 0.028 | -0.20 |
|  | GE86989 | NM_176813 | *AGR3* | 0.002 | -0.20 |
|  | GE614549 | NM_173632 | *ZNF776* | 0.010 | -0.20 |
|  | GE81159 | NM_002470 | *MYH3* | 0.006 | -0.20 |
|  | GE55185 | NM_033013 | *NR1I2* | 0.010 | -0.20 |
|  | GE569237 | NM_003710 | *SPINT1* | 0.003 | -0.20 |
|  | GE61335 | NM_000582 | *SPP1* | 0.007 | -0.21 |
|  | GE87105 | NM_017770 | *ELOVL2* | 0.013 | -0.21 |
|  | GE906249 | NM_145032 | *FBXL13* | 0.041 | -0.21 |
|  | GE62169 | NM_138393 | *REEP6* | 0.022 | -0.21 |
|  | GE895359 | AK023198 | *OPRK1* | 0.005 | -0.21 |
|  | GE54715 | NM_004205 | *USP2* | 0.047 | -0.21 |
|  | GE58527 | NM_013366 | *ANAPC2* | 3.38x10^-04^ | -0.21 |
|  | GE81488 | NM_005295 | *GPR22* | 0.024 | -0.21 |
|  | GE900096 | BC003524 | *LINC00589* | 0.040 | -0.21 |
|  | GE81930 | NM_014474 | *SMPDL3B* | 0.029 | -0.21 |
|  | GE87131 | NM_017655 | *GIPC2* | 0.012 | -0.21 |
|  | GE79288 | NM_006498 | *LGALS2* | 0.045 | -0.21 |
|  | GE56228 | NM_015658 | *NOC2L* | 0.001 | -0.21 |
|  | GE59182 | NM_014422 | *INPP5J* | 0.025 | -0.22 |
|  | GE773863 | AK125951 | *ARHGEF38* | 0.010 | -0.22 |
|  | GE79404 | NM_012391 | *SPDEF* | 0.014 | -0.22 |
|  | GE54290 | NM_003712 | *PLPP2* | 0.001 | -0.22 |
|  | GE82911 | NM_024719 | *GRTP1* | 2.85x10^-04^ | -0.22 |
|  | GE59707 | NM_007127 | *VIL1* | 0.014 | -0.22 |
|  | GE899734 | NM_198571 | *NAT16* | 2.92x10^-04^ | -0.22 |
|  | GE56928 | NM_024599 | *RHBDF2* | 0.010 | -0.22 |
|  | GE83118 | BC050096 | *SLC35F5* | 0.017 | -0.22 |
|  | GE479489 | BC024191 | *OPALIN* | 0.006 | -0.22 |
|  | GE593235 | AK026327 | *POLR2F* | 0.041 | -0.22 |
|  | GE57617 | NM_001977 | *ENPEP* | 0.044 | -0.22 |
|  | GE662502 | NM_173533 | *TDRD5* | 0.003 | -0.23 |
|  | GE54549 | NM_006574 | *CSPG5* | 0.004 | -0.23 |
|  | GE81381 | NM_004306 | *ANXA13* | 0.010 | -0.23 |
|  | GE80061 | NM_002273 | *KRT8* | 0.030 | -0.23 |
|  | GE516052 | NM_153699 | *GSTA5* | 0.041 | -0.23 |
|  | GE867997 | BU608190 | *LRRIQ1* | 0.002 | -0.23 |
|  | GE82444 | NM_018725 | *IL17RB* | 0.003 | -0.23 |
|  | GE80605 | NM_020633 | *VN1R1* | 0.005 | -0.23 |
|  | GE55847 | NM_018204 | *CKAP2* | 0.001 | -0.23 |
|  | GE62674 | NM_031457 | *MS4A8* | 0.022 | -0.23 |
|  | GE60154 | NM_004563 | *PCK2* | 0.007 | -0.23 |
|  | GE85843 | NM_030793 | *FBXO38* | 0.001 | -0.23 |
|  | GE475763 | BC034724 | *LOC101928797* | 0.005 | -0.23 |
|  | GE56272 | NM_020425 | *SMIM8* | 0.007 | -0.23 |
|  | GE62004 | NM_033128 | *SCIN* | 0.003 | -0.24 |
|  | GE640715 | NM_173661 | *LINC00955* | 0.013 | -0.24 |
|  | GE505673 | NM_173853 | *KRTCAP3* | 0.021 | -0.24 |
|  | GE62376 | NM_022449 | *RAB17* | 0.014 | -0.24 |
|  | GE81661 | NM_006615 | *CAPN9* | 0.003 | -0.24 |
|  | GE674996 | AK128431 | *MORN1* | 0.004 | -0.24 |
|  | GE79278 | NM_000790 | *DDC* | 0.013 | -0.24 |
|  | GE82721 | NM_022772 | *EPS8L2* | 0.001 | -0.25 |
|  | GE79293 | NM_001306 | *CLDN3* | 0.018 | -0.25 |
|  | GE79517 | NM_003284 | *TNP1* | 0.007 | -0.25 |
|  | GE569462 | NM_053039 | *UGT2B28* | 0.041 | -0.25 |
|  | GE53902 | NM_020771 | *HACE1* | 0.045 | -0.25 |
|  | GE470208 | BC027906 | *LINC01007* | 0.008 | -0.25 |
|  | GE79182 | NM_000422 | *KRT17* | 0.014 | -0.25 |
|  | GE59041 | NM_000150 | *FUT6* | 0.004 | -0.25 |
|  | GE81191 | NM_002771 | *PRSS3* | 0.006 | -0.25 |
|  | GE59037 | NM_000196 | *HSD11B2* | 0.006 | -0.25 |
|  | GE61493 | NM_018424 | *EPB41L4B* | 0.002 | -0.26 |
|  | GE80998 | NM_001265 | *CDX2* | 0.037 | -0.26 |
|  | GE899053 | NM_017662 | *TRPM6* | 0.003 | -0.26 |
|  | GE61466 | NM_002381 | *MATN3* | 0.043 | -0.26 |
|  | GE57367 | NM_001001550 | *GRB10* | 0.036 | -0.26 |
|  | GE566577 | NM_198151 | *HEPACAM2* | 0.005 | -0.26 |
|  | GE786703 | AK026416 | *C3orf85* | 0.031 | -0.26 |
|  | GE555183 | U53531 | *DYNC2H1* | 0.011 | -0.26 |
|  | GE82735 | NM_022901 | *LRRC19* | 0.024 | -0.27 |
|  | GE61903 | NM_025052 | *MAP3K19* | 1.30x10^-04^ | -0.27 |
|  | GE81754 | NM_012114 | *CASP14* | 0.004 | -0.27 |
|  | GE544889 | AW513823 | *TRAF3IP2* | 0.008 | -0.27 |
|  | GE54006 | NM_014428 | *TJP3* | 0.043 | -0.27 |
|  | GE56110 | NM_207034 | *EDN3* | 0.024 | -0.27 |
|  | GE81808 | NM_013250 | *ZNF215* | 0.001 | -0.28 |
|  | GE753147 | BX095052 | *NEURL1-AS1* | 0.003 | -0.28 |
|  | GE79848 | NM_144575 | *CAPN13* | 0.016 | -0.28 |
|  | GE79279 | NM_004963 | *GUCY2C* | 0.026 | -0.29 |
|  | GE80684 | NM_014406 | *CCT8L2* | 0.016 | -0.29 |
|  | GE82535 | NM_020387 | *RAB25* | 0.045 | -0.29 |
|  | GE484420 | BC031618 | *BEND7* | 0.000 | -0.30 |
|  | GE87886 | NM_181723 | *MICU3* | 0.025 | -0.30 |
|  | GE769224 | BC052596 | *TMEM238* | 0.004 | -0.30 |
|  | GE60082 | NM_004059 | *KYAT1* | 0.008 | -0.30 |
|  | GE898026 | AL832375 | *XIRP2* | 0.001 | -0.30 |
|  | GE574557 | AJ417849 | *LRATD2* | 0.002 | -0.32 |
|  | GE538595 | AY040089 | *LINC00112* | 0.002 | -0.32 |
|  | GE537800 | AK057198 | *STK4-DT* | 0.003 | -0.32 |
|  | GE897460 | NM_058244 | *WNT8A* | 0.050 | -0.33 |
|  | GE54724 | NM_003504 | *CDC45* | 0.001 | -0.33 |
|  | GE57850 | NM_000562 | *C8A* | 4.86x10^-04^ | -0.34 |
|  | GE54196 | NM_006863 | *LILRA1* | 0.004 | -0.34 |
|  | GE749628 | NM_147191 | *MMP21* | 0.015 | -0.35 |
|  | GE61547 | NM_005927 | *MFAP3* | 0.002 | -0.35 |
|  | GE83286 | NM_031952 | *SPATA9* | 0.006 | -0.36 |
|  | GE901404 | AW592188 | *LINC00254* | 0.003 | -0.37 |
|  | GE503821 | NM_145865 | *ANKS4B* | 0.005 | -0.37 |
|  | GE57519 | NM_000777 | *CYP3A5* | 0.014 | -0.38 |
|  | GE59726 | NM_000242 | *MBL2* | 0.047 | -0.38 |
|  | GE81519 | NM_005483 | *CHAF1A* | 0.000 | -0.39 |
|  | GE61627 | NM_139211 | *HOPX* | 0.002 | -0.41 |
|  | GE555486 | AL117582 | *MYRF-AS1* | 0.035 | -0.43 |
|  | GE83431 | AB044556 | *PARD6G* | 0.028 | -0.43 |
|  | GE808206 | BX102688 | *LINC00570* | 0.001 | -0.45 |
|  | GE872937 | BQ189302 | *LINC01229* | 0.027 | -0.48 |
|  | GE882846 | AB095935 | *ANKRD18A* | 0.002 | -0.79 |
|  | GE555851 | NM_018608 | *RBM12B-AS1* | 0.004 | -0.80 |
|  | GE81014 | NM_001364 | *DLG2* | 0.006 | -0.82 |
|  | GE892634 | NM_199234 | *GDNF* | 0.044 | -0.85 |
|  | GE62813 | NM_014753 | *BMS1* | 0.009 | -0.93 |
|  | GE486546 | NM_002854 | *PVALB* | 0.005 | -1.71 |
| **Inflamed** | GE477882 | AL832235 | *CENPW* | 0.002 | 0.42 |
|  | GE687057 | BC034570 | *LINC00452* | 0.004 | 0.37 |
|  | GE80143 | NM_015714 | *G0S2* | 0.008 | 0.36 |
|  | GE58336 | NM_004466 | *GPC5* | 0.006 | 0.36 |
|  | GE56425 | X52332 | *ZNF10* | 0.010 | 0.32 |
|  | GE80257 | NM_020142 | *NDUFA4L2* | 0.005 | 0.30 |
|  | GE478868 | NM_001890 | *CSN1S1* | 0.033 | 0.30 |
|  | GE82459 | NM_018961 | *UBASH3A* | 0.001 | 0.29 |
|  | GE53986 | NM_000095 | *COMP* | 0.024 | 0.29 |
|  | GE56063 | NM_015166 | *MLC1* | 7.28x10^-05^ | 0.29 |
|  | GE53207 | NM_014787 | *DNAJC6* | 0.021 | 0.29 |
|  | GE60216 | NM_000474 | *TWIST1* | 0.001 | 0.28 |
|  | GE562001 | AK129955 | *CCDC178* | 0.033 | 0.27 |
|  | GE612661 | AL137270 | *LINC00939* | 0.036 | 0.26 |
|  | GE531062 | BC046439 | *ZNF252P-AS1* | 0.014 | 0.26 |
|  | GE62917 | NM_000570 | *FCGR3B* | 0.002 | 0.26 |
|  | GE80047 | NM_004194 | *ADAM22* | 0.002 | 0.26 |
|  | GE59684 | NM_000163 | *GHR* | 0.029 | 0.26 |
|  | GE57503 | NM_000570 | *FCGR3B* | 0.003 | 0.25 |
|  | GE85870 | NM_001276 | *CHI3L1* | 0.028 | 0.25 |
|  | GE56492 | NM_181690 | *AKT3* | 3.73x10^-04^ | 0.25 |
|  | GE57356 | NM_004362 | *CLGN* | 0.023 | 0.25 |
|  | GE604079 | AK096708 | *EBF1* | 0.002 | 0.25 |
|  | GE57381 | AF200348 | *PXDN* | 4.02x10^-04^ | 0.25 |
|  | GE753662 | BC031660 | *C3orf80* | 0.003 | 0.25 |
|  | GE81474 | NM_005121 | *MED13* | 0.004 | 0.24 |
|  | GE82313 | NM_018166 | *EVA1B* | 0.002 | 0.24 |
|  | GE57493 | NM_000632 | *ITGAM* | 0.004 | 0.24 |
|  | GE79587 | NM_001845 | *COL4A1* | 1.36x10^-04^ | 0.24 |
|  | GE59892 | NM_000138 | *FBN1* | 0.001 | 0.24 |
|  | GE54222 | NM_014459 | *PCDH17* | 2.07x10^-04^ | 0.24 |
|  | GE57889 | NM_002619 | *PF4* | 2.48x10^-04^ | 0.24 |
|  | GE614935 | X89067 | *TRPC2* | 0.009 | 0.24 |
|  | GE80442 | NM_001453 | *FOXC1* | 0.002 | 0.23 |
|  | GE81575 | NM_005924 | *MEOX2* | 0.002 | 0.23 |
|  | GE80405 | NM_152718 | *VWCE* | 0.002 | 0.23 |
|  | GE58895 | NM_006169 | *NNMT* | 1.22x10^-04^ | 0.23 |
|  | GE55352 | AF533709 | *HAUS6* | 0.007 | 0.23 |
|  | GE62778 | NM_001385 | *DPYS* | 0.003 | 0.23 |
|  | GE61925 | NM_021972 | *SPHK1* | 0.006 | 0.22 |
|  | GE62178 | NM_032609 | *COX4I2* | 0.016 | 0.22 |
|  | GE476683 | NM_138453 | *RAB3C* | 0.001 | 0.22 |
|  | GE79611 | NM_138440 | *VASN* | 0.001 | 0.22 |
|  | GE83553 | NM_032849 | *MEDAG* | 1.04x10^-04^ | 0.22 |
|  | GE53467 | NM_015234 | *ADGRF5* | 2.23x10^-04^ | 0.22 |
|  | GE62586 | NM_002593 | *PCOLCE* | 0.003 | 0.22 |
|  | GE87262 | NM_000230 | *LEP* | 0.049 | 0.22 |
|  | GE559599 | AL832955 | *STEAP4* | 8.12x10^-05^ | 0.22 |
|  | GE753217 | AF107456 | *LINC00244* | 0.005 | 0.22 |
|  | GE80191 | AK127826 | *FNDC3B* | 0.001 | 0.22 |
|  | GE60059 | NM_001795 | *CDH5* | 3.28x10^-04^ | 0.22 |
|  | GE87323 | NM_024560 | *ACSS3* | 2.54x10^-04^ | 0.22 |
|  | GE87899 | NM_144647 | *CAPSL* | 0.027 | 0.22 |
|  | GE81856 | NM_014059 | *RGCC* | 0.004 | 0.21 |
|  | GE53877 | NM_016242 | *EMCN* | 0.001 | 0.21 |
|  | GE553808 | NM_022162 | *NOD2* | 0.001 | 0.21 |
|  | GE52967 | NM_014906 | *PPM1E* | 0.005 | 0.21 |
|  | GE53641 | NM_016201 | *AMOTL2* | 0.020 | 0.21 |
|  | GE54995 | NM_016580 | *PCDH12* | 2.97x10^-04^ | 0.21 |
|  | GE79111 | NM_002934 | *RNASE2* | 0.050 | 0.21 |
|  | GE87894 | BC025243 | *CCDC8* | 0.001 | 0.21 |
|  | GE56612 | AL133118 | *EMCN* | 0.001 | 0.21 |
|  | GE59872 | NM_001957 | *EDNRA* | 0.001 | 0.21 |
|  | GE58126 | NM_003069 | *SMARCA1* | 3.32x10^-04^ | 0.21 |
|  | GE57179 | NM_006207 | *PDGFRL* | 0.001 | 0.21 |
|  | GE58210 | NM_000165 | *GJA1* | 0.001 | 0.21 |
|  | GE510002 | NM_003764 | *STX11* | 0.003 | 0.21 |
|  | GE80447 | NM_002432 | *MNDA* | 0.008 | 0.20 |
|  | GE53313 | BC062365 | *SLIT3* | 0.001 | 0.20 |
|  | GE81826 | NM_013356 | *SLC16A8* | 0.003 | 0.20 |
|  | GE533132 | NM_139319 | *SLC17A8* | 0.007 | 0.20 |
|  | GE79216 | NM_000954 | *PTGDS* | 0.001 | 0.20 |
|  | GE80504 | NM_002897 | *RBMS1* | 1.72x10^-04^ | 0.20 |
|  | GE80543 | NM_205843 | *NFIC* | 0.001 | 0.20 |
|  | GE79151 | NM_000134 | *FABP2* | 2.35x10^-04^ | -0.20 |
|  | GE81271 | NM_003379 | *EZR* | 4.94x10^-05^ | -0.20 |
|  | GE542626 | NM_153226 | *SLC35G1* | 0.002 | -0.20 |
|  | GE82051 | NM_205835 | *LSR* | 0.001 | -0.20 |
|  | GE87249 | NM_145252 | *ZG16B* | 0.003 | -0.20 |
|  | GE523088 | NM_031204 | *CABP2* | 2.48x10^-04^ | -0.20 |
|  | GE62206 | NM_000403 | *GALE* | 0.001 | -0.20 |
|  | GE54140 | NM_004668 | *MGAM* | 0.027 | -0.20 |
|  | GE58135 | NM_003475 | *RASSF7* | 2.18x10^-04^ | -0.20 |
|  | GE62347 | NM_004003 | *CRAT* | 0.001 | -0.20 |
|  | GE81963 | NM_014867 | *KBTBD11* | 2.29x10^-05^ | -0.20 |
|  | GE662502 | NM_173533 | *TDRD5* | 0.026 | -0.20 |
|  | GE85545 | NM_015533 | *TKFC* | 0.001 | -0.20 |
|  | GE88138 | NM_153715 | *HOXA10* | 0.001 | -0.20 |
|  | GE55389 | NM_006714 | *SMPDL3A* | 0.001 | -0.20 |
|  | GE58157 | NM_000718 | *CACNA1B* | 0.006 | -0.20 |
|  | GE552567 | NM_207468 | *FAM177B* | 2.12x10^-04^ | -0.21 |
|  | GE82202 | NM_017671 | *FERMT1* | 1.96x10^-04^ | -0.21 |
|  | GE85517 | NM_139160 | *DEPDC7* | 8.35x10^-05^ | -0.21 |
|  | GE79161 | NM_003225 | *TFF1* | 0.007 | -0.21 |
|  | GE565507 | NM_052926 | *PNMA5* | 0.011 | -0.21 |
|  | GE82225 | NM_017791 | *FLVCR2* | 4.30x10^-04^ | -0.21 |
|  | GE81237 | NM_003122 | *SPINK1* | 4.55x10^-04^ | -0.21 |
|  | GE59894 | NM_001041 | *SI* | 0.003 | -0.21 |
|  | GE82620 | NM_021813 | *BACH2* | 0.011 | -0.21 |
|  | GE54632 | NM_030928 | *CDT1* | 9.31x10^-05^ | -0.21 |
|  | GE81111 | NM_002083 | *GPX2* | 1.77x10^-04^ | -0.21 |
|  | GE62169 | NM_138393 | *REEP6* | 0.010 | -0.21 |
|  | GE86951 | NM_024320 | *PRR15L* | 1.81x10^-04^ | -0.21 |
|  | GE58117 | NM_003318 | *TTK* | 0.003 | -0.21 |
|  | GE62217 | NM_020672 | *S100A14* | 9.31x10^-05^ | -0.21 |
|  | GE57732 | NM_004360 | *CDH1* | 6.70x10^-05^ | -0.21 |
|  | GE54262 | NM_001081 | *CUBN* | 0.001 | -0.21 |
|  | GE897986 | NM_032229 | *SLITRK6* | 0.003 | -0.21 |
|  | GE55443 | NM_014576 | *A1CF* | 1.51x10^-04^ | -0.21 |
|  | GE58542 | NM_001954 | *DDR1* | 7.69x10^-05^ | -0.21 |
|  | GE55366 | AK024850 | *FZD5* | 0.001 | -0.21 |
|  | GE484935 | NM_002409 | *MGAT3* | 0.002 | -0.21 |
|  | GE57536 | NM_000531 | *OTC* | 0.011 | -0.21 |
|  | GE79843 | NM_018414 | *ST6GALNAC1* | 0.001 | -0.21 |
|  | GE58215 | NM_000346 | *SOX9* | 2.89x10^-04^ | -0.21 |
|  | GE87002 | NM_001046 | *SLC12A2* | 8.82x10^-05^ | -0.21 |
|  | GE88450 | NM_024422 | *DSC2* | 3.20x10^-04^ | -0.22 |
|  | GE57102 | NM_006507 | *REG1B* | 0.004 | -0.22 |
|  | GE523387 | AK023445 | *ZNF503-AS1* | 0.001 | -0.22 |
|  | GE79528 | NM_006142 | *SFN* | 0.001 | -0.22 |
|  | GE81190 | NM_002770 | *PRSS2* | 0.004 | -0.22 |
|  | GE546293 | NM_004959 | *NR5A1* | 0.003 | -0.22 |
|  | GE512913 | AI125340 | *ADAM7-AS2* | 1.68x10^-04^ | -0.22 |
|  | GE62517 | NM_022128 | *RBKS* | 0.001 | -0.22 |
|  | GE57395 | NM_015027 | *PDXDC1* | 0.002 | -0.22 |
|  | GE870970 | BX647688 | *SYTL5* | 0.002 | -0.22 |
|  | GE62191 | NM_006645 | *STARD10* | 0.000 | -0.22 |
|  | GE541633 | NM_001011720 | *XKR9* | 0.002 | -0.22 |
|  | GE58733 | NM_016445 | *PLEK2* | 0.002 | -0.22 |
|  | GE60469 | NM_002509 | *NKX2-2* | 4.78x10^-04^ | -0.22 |
|  | GE62552 | NM_003121 | *SPIB* | 0.022 | -0.22 |
|  | GE528842 | BX648070 | *C2orf88* | 4.67x10^-05^ | -0.22 |
|  | GE54024 | NM_000187 | *HGD* | 5.84x10^-05^ | -0.23 |
|  | GE541008 | NM_019010 | *KRT20* | 7.69x10^-05^ | -0.23 |
|  | GE563797 | NM_000870 | *HTR4* | 1.51x10^-04^ | -0.23 |
|  | GE60498 | NM_002153 | *HSD17B2* | 7.48x10^-05^ | -0.23 |
|  | GE59130 | NM_004496 | *FOXA1* | 2.41x10^-04^ | -0.23 |
|  | GE54873 | NM_173570 | *ZDHHC23* | 5.08x10^-05^ | -0.23 |
|  | GE54673 | NM_002407 | *SCGB2A1* | 0.005 | -0.23 |
|  | GE57442 | NM_000015 | *NAT2* | 0.000 | -0.23 |
|  | GE80101 | NM_000789 | *ACE* | 0.009 | -0.23 |
|  | GE80998 | NM_001265 | *CDX2* | 2.89x10^-04^ | -0.23 |
|  | GE55905 | NM_018265 | *INAVA* | 1.22x10^-04^ | -0.23 |
|  | GE56769 | NM_030574 | *STARD5* | 0.001 | -0.23 |
|  | GE802322 | NM_032435 | *MAP3K21* | 0.001 | -0.23 |
|  | GE56076 | NM_004415 | *DSP* | 2.18x10^-04^ | -0.23 |
|  | GE56013 | NM_152695 | *ZNF449* | 2.61x10^-04^ | -0.23 |
|  | GE55951 | NM_005486 | *TOM1L1* | 0.001 | -0.23 |
|  | GE79295 | NM_004210 | *NEURL1* | 2.85x10^-04^ | -0.23 |
|  | GE81930 | NM_014474 | *SMPDL3B* | 1.59x10^-04^ | -0.23 |
|  | GE59398 | NM_005035 | *POLRMT* | 1.16x10^-04^ | -0.23 |
|  | GE62493 | NM_014270 | *SLC7A9* | 0.009 | -0.24 |
|  | GE762301 | AB041269 | *KRT19P2* | 1.39x10^-04^ | -0.24 |
|  | GE86461 | NM_181353 | *ID1* | 0.001 | -0.24 |
|  | GE62652 | NM_012128 | *CLCA4* | 0.019 | -0.24 |
|  | GE56273 | AL050204 | *EXPH5* | 1.01x10^-04^ | -0.24 |
|  | GE594353 | AK090739 | *FLVCR1* | 1.09x10^-04^ | -0.24 |
|  | GE80896 | NM_000149 | *FUT3* | 3.12x10^-04^ | -0.24 |
|  | GE81139 | NM_002276 | *KRT19* | 1.96x10^-04^ | -0.24 |
|  | GE80061 | NM_002273 | *KRT8* | 0.001 | -0.24 |
|  | GE54166 | NM_000847 | *GSTA3* | 0.002 | -0.24 |
|  | GE79099 | NM_145740 | *GSTA1* | 0.001 | -0.24 |
|  | GE85596 | NM_002054 | *GCG* | 0.001 | -0.24 |
|  | GE512985 | NM_001011719 | *ARSH* | 2.65x10^-05^ | -0.24 |
|  | GE86989 | NM_176813 | *AGR3* | 5.52x10^-05^ | -0.25 |
|  | GE841843 | NM_001012993 | *C9orf152* | 1.07x10^-04^ | -0.25 |
|  | GE489203 | NM_005193 | *CDX4* | 1.86x10^-04^ | -0.25 |
|  | GE477081 | NM_173481 | *MISP* | 6.52x10^-05^ | -0.25 |
|  | GE81755 | NM_012116 | *CBLC* | 1.32x10^-04^ | -0.25 |
|  | GE873658 | BC075833 | *ABCC6P1* | 0.001 | -0.25 |
|  | GE82487 | NM_019894 | *TMPRSS4* | 1.72x10^-04^ | -0.25 |
|  | GE58214 | NM_000340 | *SLC2A2* | 0.028 | -0.25 |
|  | GE79763 | NM_001197 | *BIK* | 3.92x10^-04^ | -0.25 |
|  | GE62291 | NM_001307 | *CLDN7* | 1.01x10^-04^ | -0.25 |
|  | GE82908 | NM_024712 | *ELMO3* | 8.12x10^-05^ | -0.25 |
|  | GE549235 | NM_153338 | *GGT6* | 0.001 | -0.25 |
|  | GE475965 | NM_013430 | *GGT1* | 0.002 | -0.25 |
|  | GE79395 | NM_033553 | *GUCA2A* | 1.09x10^-04^ | -0.25 |
|  | GE53544 | NM_005358 | *LMO7* | 2.04x10^-05^ | -0.25 |
|  | GE88582 | NM_018667 | *SMPD3* | 4.41x10^-05^ | -0.26 |
|  | GE61082 | NM_020299 | *AKR1B10* | 4.12x10^-04^ | -0.26 |
|  | GE88714 | NM_024616 | *C3orf52* | 8.58x10^-05^ | -0.26 |
|  | GE549575 | CR749811 | *LRRC66* | 1.25x10^-04^ | -0.26 |
|  | GE54213 | NM_006017 | *PROM1* | 9.31x10^-05^ | -0.26 |
|  | GE79754 | NM_033520 | *C19orf33* | 7.28x10^-05^ | -0.26 |
|  | GE56254 | AL832022 | *RMND5A* | 0.002 | -0.26 |
|  | GE525098 | BC050337 | *LHFPL3-AS2* | 3.23x10^-05^ | -0.26 |
|  | GE498144 | NM_032874 | *CCDC183* | 0.019 | -0.26 |
|  | GE86468 | BC030587 | *PIP5K1B* | 4.67x10^-05^ | -0.26 |
|  | GE58102 | NM_005588 | *MEP1A* | 2.23x10^-04^ | -0.26 |
|  | GE505673 | NM_173853 | *KRTCAP3* | 4.54x10^-05^ | -0.27 |
|  | GE82757 | AL832940 | *C1orf116* | 4.12x10^-04^ | -0.27 |
|  | GE82815 | NM_024306 | *FA2H* | 1.43x10^-04^ | -0.27 |
|  | GE79541 | NM_002483 | *CEACAM6* | 0.024 | -0.27 |
|  | GE840298 | BX648086 | *CORO2A* | 7.28x10^-05^ | -0.27 |
|  | GE81191 | NM_002771 | *PRSS3* | 1.07x10^-04^ | -0.28 |
|  | GE57667 | NM_002670 | *PLS1* | 7.48x10^-05^ | -0.28 |
|  | GE60152 | NM_003558 | *PIP5K1B* | 6.52x10^-05^ | -0.28 |
|  | GE58576 | NM_014252 | *SLC25A15* | 6.17x10^-05^ | -0.28 |
|  | GE82535 | NM_020387 | *RAB25* | 1.04x10^-04^ | -0.28 |
|  | GE514337 | NM_173560 | *RFX6* | 0.001 | -0.28 |
|  | GE57437 | NM_014465 | *SULT1B1* | 9.81x10^-05^ | -0.28 |
|  | GE59346 | NM_005242 | *F2RL1* | 1.68x10^-04^ | -0.28 |
|  | GE557257 | NM_001008494 | *ISX* | 9.57x10^-05^ | -0.28 |
|  | GE85829 | AB033037 | *CRACD* | 2.16x10^-05^ | -0.28 |
|  | GE87574 | NM_014471 | *SPINK4* | 0.018 | -0.28 |
|  | GE53321 | NM_033504 | *TMEM54* | 1.47x10^-04^ | -0.29 |
|  | GE80330 | NM_005971 | *FXYD3* | 1.51x10^-04^ | -0.29 |
|  | GE773863 | AK125951 | *ARHGEF38* | 8.82x10^-05^ | -0.29 |
|  | GE516887 | AK056882 | *RASEF* | 3.23x10^-05^ | -0.30 |
|  | GE85915 | NM_003122 | *SPINK1* | 0.001 | -0.30 |
|  | GE82187 | NM_017625 | *ITLN1* | 0.002 | -0.31 |
|  | GE54595 | NM_005727 | *TSPAN1* | 4.78x10^-04^ | -0.32 |
|  | GE61812 | NM_021969 | *NR0B2* | 0.001 | -0.33 |
|  | GE57275 | NM_003648 | *DGKD* | 2.68x10^-04^ | -0.34 |
|  | GE79812 | NM_001910 | *CTSE* | 0.016 | -0.50 |
|  | GE577124 | NM_182532 | *TMEM61* | 3.82x10^-04^ | -0.52 |

FC, forld change; FDR, false discovery rate; GEO, gene expression omnibus; IBD, inflammatory bowel disease; POR, post-operative recurrence.

***Table S7.*** *DEseq2 results: differentially expressed bacterial families when comparing the IBD-free controls towards POR in both mucosal zones.*

|  |  | **Family** | **baseMean** | **log_2_FC** | **P-value** | **P-adj** |
| --- | --- | --- | --- | --- | --- | --- |
| **Non-POR vs IBD-free controls** | **Uninflamed** | Thermaceae | 24.59 | 18.88 | 0.001 | 0.007 |
|  |  | Flavobacteriaceae | 9.77 | 19.17 | 4.51x10^-06^ | 5.69x10^-05^ |
|  |  | Pseudomonadaceae | 52.50 | -22.24 | 0.001 | 0.006 |
|  |  | Campylobacteraceae | 17.48 | 18.08 | 0.001 | 0.006 |
|  |  | Vibrionaceae | 3.40 | -17.38 | 0.007 | 0.036 |
|  |  | Methylobacteriaceae | 22.06 | 16.58 | 1.51x10^-06^ | 2.75x10^-05^ |
|  |  | Caulobacteraceae | 11.74 | 17.51 | 4.99x10^-05^ | 4.05x10^-04^ |
|  |  | Bradyrhizobiaceae | 8.69 | 26.19 | 0.001 | 0.004 |
|  |  | Rhodobacteraceae | 67.55 | -25.07 | 1.26x10^-20^ | 9.22x10^-19^ |
|  | **Inflamed** | Thermaceae | 24.59 | 16.62 | 0.004 | 0.028 |
|  |  | Cytophagaceae | 2.98 | 30.00 | 6.96x10^-05^ | 0.001 |
|  |  | Flavobacteriaceae | 9.77 | 15.88 | 1.28x10^-04^ | 0.001 |
|  |  | Pseudomonadaceae | 52.50 | 22.38 | 0.001 | 0.007 |
|  |  | Campylobacteraceae | 17.48 | 18.94 | 4.40x10^-04^ | 0.004 |
|  |  | Methylobacteriaceae | 22.06 | 21.47 | 2.36x10^-10^ | 8.59x10^-09^ |
|  |  | Caulobacteraceae | 11.74 | 18.06 | 2.30x10^-05^ | 3.13x10^-04^ |
|  |  | Rhodobacteraceae | 67.55 | 19.91 | 2.73x10^-14^ | 2.97x10^-12^ |
|  |  | Bacillaceae | 47.56 | 7.33 | 0.002 | 0.018 |
| **POR vs IBD-free controls** | **Uninflamed** | Thermaceae | 24.59 | 19.60 | 2.19x10^-04^ | 0.002 |
|  |  | Flavobacteriaceae | 9.77 | 20.98 | 2.93x10^-08^ | 4.92x10^-07^ |
|  |  | Pseudomonadaceae | 52.50 | 17.52 | 0.004 | 0.024 |
|  |  | Campylobacteraceae | 17.48 | 18.20 | 2.27x10^-04^ | 0.002 |
|  |  | Vibrionaceae | 3.40 | 17.51 | 0.003 | 0.015 |
|  |  | Methylobacteriaceae | 22.06 | 19.57 | 3.42x10^-10^ | 6.91x10^-09^ |
|  |  | Caulobacteraceae | 11.74 | -25.25 | 1.27x10^-10^ | 3.22x10^-09^ |
|  |  | Rhodobacteraceae | 67.55 | 20.13 | 7.71x10^-17^ | 4.64x10^-15^ |
|  |  | Streptococcaceae | 1806.58 | 3.24 | 0.005 | 0.027 |
|  |  | Bacillaceae | 47.56 | 6.91 | 0.002 | 0.012 |
|  | **Inflamed** | Thermaceae | 24.59 | 20.33 | 1.18x10^-04^ | 0.001 |
|  |  | Flavobacteriaceae | 9.77 | 21.10 | 2.12x10^-08^ | 2.93x10^-07^ |
|  |  | Pseudomonadaceae | 52.50 | 23.98 | 8.26x10^-05^ | 0.001 |
|  |  | Campylobacteraceae | 17.48 | 17.09 | 0.001 | 0.003 |
|  |  | Vibrionaceae | 3.40 | 18.33 | 0.002 | 0.007 |
|  |  | Methylobacteriaceae | 22.06 | 21.71 | 2.58x10^-12^ | 4.96x10^-11^ |
|  |  | Caulobacteraceae | 11.74 | 20.52 | 1.36x10^-07^ | 1.31x10^-06^ |
|  |  | Bradyrhizobiaceae | 8.69 | 22.11 | 0.001 | 0.007 |
|  |  | Rhodobacteraceae | 67.55 | 22.25 | 2.17x10^-20^ | 1.67x10^-18^ |
|  |  | Bacillaceae | 47.56 | 7.20 | 0.001 | 0.006 |
| **Mild-recurrence vs IBD-free controls** | **Uninflamed** | Nitrososphaeraceae | 2.38 | -24.13 | 0.001 | 0.004 |
|  |  | Thermaceae | 24.59 | 18.36 | 0.001 | 0.007 |
|  |  | Mycobacteriaceae | 5.94 | 18.10 | 7.91x10^-05^ | 0.001 |
|  |  | Flavobacteriaceae | 9.77 | 19.34 | 1.51x10^-06^ | 1.68x10^-05^ |
|  |  | Campylobacteraceae | 17.48 | 17.66 | 0.001 | 0.004 |
|  |  | Vibrionaceae | 10.18 | 16.90 | 2.29x10^-04^ | 0.002 |
|  |  | Methylobacteriaceae | 22.06 | 19.36 | 3.12x10^-09^ | 7.80x10^-08^ |
|  |  | Caulobacteraceae | 11.74 | -22.85 | 5.40x10^-08^ | 1.08x10^-06^ |
|  |  | Rhodobacteraceae | 67.55 | 17.34 | 2.22x10^-12^ | 7.39x10^-11^ |
|  |  | Streptococcaceae | 1806.58 | 3.54 | 0.003 | 0.014 |
|  |  | Bacillaceae | 47.56 | 6.79 | 0.003 | 0.014 |
|  | **Inflamed** | Nitrososphaeraceae | 2.38 | -25.34 | 3.20x10^-04^ | 0.002 |
|  |  | Thermaceae | 24.59 | 20.54 | 2.75x10^-04^ | 0.002 |
|  |  | Mycobacteriaceae | 5.94 | 18.85 | 3.70x10^-05^ | 3.03x10^-04^ |
|  |  | Flavobacteriaceae | 9.77 | 20.97 | 1.63x10^-07^ | 2.44x10^-06^ |
|  |  | Campylobacteraceae | 17.48 | 14.98 | 0.004 | 0.021 |
|  |  | Vibrionaceae | 10.18 | 18.00 | 8.19x10^-05^ | 0.001 |
|  |  | Enterobacteriaceae | 6567.17 | -3.14 | 0.007 | 0.036 |
|  |  | Methylobacteriaceae | 22.06 | 21.42 | 4.70x10^-11^ | 1.06x10^-09^ |
|  |  | Caulobacteraceae | 11.74 | 19.81 | 1.99x10^-06^ | 1.79x10^-05^ |
|  |  | Rhodobacteraceae | 67.55 | 17.95 | 3.11x10^-13^ | 1.40x10^-11^ |
|  |  | Bacillaceae | 47.56 | 7.11 | 0.002 | 0.009 |
| **Severe-POR vs IBD-free controls** | **Uninflamed** | Nitrososphaeraceae | 2.38 | -21.12 | 0.007 | 0.036 |
|  |  | Thermaceae | 24.59 | 16.66 | 0.008 | 0.038 |
|  |  | Mycobacteriaceae | 5.94 | 19.05 | 1.79x10^-04^ | 0.002 |
|  |  | Verrucomicrobiaceae | 132.53 | -18.01 | 0.002 | 0.010 |
|  |  | Flavobacteriaceae | 9.77 | 22.33 | 5.17x10^-07^ | 7.38x10^-06^ |
|  |  | Campylobacteraceae | 17.48 | 18.69 | 0.001 | 0.008 |
|  |  | Vibrionaceae | 10.18 | 14.42 | 0.005 | 0.026 |
|  |  | Methylobacteriaceae | 22.06 | 17.37 | 1.66x10^-06^ | 2.08x10^-05^ |
|  |  | Caulobacteraceae | 11.74 | -19.62 | 2.60x10^-05^ | 2.89x10^-04^ |
|  |  | Rhodobacteraceae | 67.55 | 16.45 | 1.46x10^-09^ | 2.92x10^-08^ |
|  |  | Peptostreptococcaceae | 0.19 | 23.17 | 0.003 | 0.020 |
|  | **Inflamed** | Nitrososphaeraceae | 2.38 | -21.78 | 0.005 | 0.028 |
|  |  | Mycobacteriaceae | 5.94 | 20.14 | 5.51x10^-05^ | 0.001 |
|  |  | Flavobacteriaceae | 9.77 | 21.26 | 1.18x10^-06^ | 1.47x10^-05^ |
|  |  | Campylobacteraceae | 17.48 | 21.95 | 1.06x10^-04^ | 0.001 |
|  |  | Vibrionaceae | 10.18 | 13.66 | 0.006 | 0.034 |
|  |  | Methylobacteriaceae | 22.06 | 21.03 | 3.12x10^-09^ | 6.23x10^-08^ |
|  |  | Caulobacteraceae | 11.74 | 20.87 | 4.56x10^-06^ | 5.07x10^-05^ |
|  |  | Rhodobacteraceae | 67.55 | 20.72 | 9.13x10^-15^ | 9.13x10^-13^ |
|  |  | Fusobacteriaceae | 1514.72 | 6.93 | 0.005 | 0.030 |

IBD, inflammatory bowel diseases; POR, post-operative recurrence.

***Table S8.*** *DEseq2 results: differentially expressed bacterial families between POR in both mucosal zones.*

|  |  | **Family** | **baseMean** | **log_2_FC** | **P-value** | **P-adj** |
| --- | --- | --- | --- | --- | --- | --- |
| **POR vs Non-POR** | **Uninflamed** | ML635J-40 | 0.31 | -22.03 | 2.46x10^-06^ | 6.40x10^-05^ |
|  |  | Pseudomonadaceae | 52.50 | 17.52 | 2.36x10^-05^ | 4.92x10^-04^ |
|  |  | Rhodobacteraceae | 67.55 | 21.96 | 5.83x10^-42^ | 6.06x10^-40^ |
|  |  | Gemellaceae | 229.63 | 3.96 | 0.001 | 0.013 |
|  |  | Spirochaetaceae | 0.73 | -23.30 | 6.25x10^-07^ | 2.17x10^-05^ |
|  | **Inflamed** | Aquificaceae | 1.10 | 22.15 | 1.73x10^-06^ | 6.29x10^-05^ |
|  |  | Cytophagaceae | 2.98 | -17.57 | 1.36x10^-04^ | 0.002 |
|  |  | Vibrionaceae | 3.40 | 26.36 | 1.19x10^-11^ | 1.30x10^-09^ |
|  |  | Lactobacillaceae | 333.03 | -6.42 | 2.83x10^-05^ | 0.001 |
| **Mild POR vs Non-POR** | **Uninflamed** | Micrococcaceae | 0.70 | 16.83 | 0.001 | 0.012 |
|  |  | Cytophagaceae | 2.98 | 13.83 | 0.006 | 0.049 |
|  |  | ML635J-40 | 0.31 | -21.69 | 1.85x10^-05^ | 3.70x10^-04^ |
|  |  | Comamonadaceae | 670.12 | -2.44 | 0.004 | 0.039 |
|  |  | Caulobacteraceae | 11.74 | -20.46 | 4.71x10^-12^ | 2.35x10^-10^ |
|  |  | Rhodobacteraceae | 67.55 | 21.47 | 3.53x10^-35^ | 3.53x10^-33^ |
|  |  | Gemellaceae | 229.63 | 4.09 | 0.001 | 0.015 |
|  |  | Spirochaetaceae | 0.73 | -23.08 | 5.20x10^-06^ | 1.30x10^-04^ |
|  |  | Peptostreptococcaceae | 0.19 | -20.58 | 4.76x10^-05^ | 0.001 |
|  |  | Fusobacteriaceae | 1514.72 | -5.07 | 0.002 | 0.019 |
|  | **Inflamed** | Vibrionaceae | 10.18 | 26.11 | 3.66x10^-16^ | 3.66x10^-14^ |
|  |  | Fusobacteriaceae | 1514.72 | -9.45 | 4.33x10^-09^ | 2.17x10^-07^ |
| **Severe POR vs Non-POR** | **Uninflamed** | Verrucomicrobiaceae | 132.53 | -27.11 | 1.86x10^-10^ | 6.19x10^-09^ |
|  |  | ML635J-40 | 0.31 | -20.32 | 0.001 | 0.008 |
|  |  | Enterobacteriaceae | 6567.17 | 3.13 | 0.001 | 0.016 |
|  |  | Caulobacteraceae | 11.74 | -21.02 | 1.10x10^-09^ | 2.76x10^-08^ |
|  |  | Rhodobacteraceae | 67.55 | 20.57 | 6.09x10^-25^ | 6.09x10^-23^ |
|  |  | Spirochaetaceae | 0.73 | -21.16 | 3.22x10^-04^ | 0.005 |
|  | **Inflamed** | Bifidobacteriaceae | 408.56 | -5.98 | 0.001 | 0.029 |
|  |  | Cytophagaceae | 2.98 | -28.44 | 7.74x10^-07^ | 2.61x10^-05^ |
|  |  | Vibrionaceae | 10.18 | 19.29 | 2.13x10^-07^ | 2.15x10^-05^ |
|  |  | Rhodospirillaceae | 2.92 | 21.90 | 1.48x10^-04^ | 0.004 |
| **Severe POR vs Mild POR** | **Uninflamed** | Micrococcaceae | 0.70 | -16.75 | 0.002 | 0.034 |
|  |  | Verrucomicrobiaceae | 132.53 | -18.45 | 4.06x10^-06^ | 8.12x10^-05^ |
|  |  | Prevotellaceae | 1643.13 | 6.03 | 0.004 | 0.049 |
|  |  | Enterobacteriaceae | 6567.17 | 3.17 | 0.001 | 0.008 |
|  |  | Caulobacteraceae | 11.74 | 27.40 | 4.10x10^-17^ | 4.10x10^-15^ |
|  |  | Peptostreptococcaceae | 0.19 | 27.63 | 5.21x10^-07^ | 1.30x10^-05^ |
|  |  | Fusobacteriaceae | 1514.72 | 5.13 | 0.004 | 0.048 |
|  | **Inflamed** | Prevotellaceae | 1643.13 | 6.75 | 0.001 | 0.011 |
|  |  | Cytophagaceae | 2.98 | -16.17 | 0.002 | 0.024 |
|  |  | Enterobacteriaceae | 6567.17 | 3.30 | 1.61x10^-04^ | 0.003 |
|  |  | Fusobacteriaceae | 1514.72 | 9.21 | 7.21x10^-08^ | 2.31x10^-06^ |

IBD, inflammatory bowel diseases; POR, post-operative recurrence.

***Table S9.*** *DEseq2 results: differentially expressed OTUs when comparing the IBD-free controls towards POR in both mucosal zones.*

|  |  | **OTU** | **Family** | **Genus** | **Species** | **baseMean** | **log_2_FC** | **P-value** | **P-adj** |
| --- | --- | --- | --- | --- | --- | --- | --- | --- | --- |
| **Non-POR vs IBD-free controls** | **Uninflamed** | OTU149 | Thermaceae | Thermus | Unclassified | 27.88 | 17.31 | 0.009 | 0.034 |
|  |  | OTU240 | Corynebacteriaceae | Corynebacterium | Unclassified | 3.43 | -21.75 | 0.001 | 0.005 |
|  |  | OTU323 | Verrucomicrobiaceae | Akkermansia | muciniphila | 103.75 | 22.40 | 1.03x10^-04^ | 0.001 |
|  |  | OTU358 | Prevotellaceae | Prevotella | nigrescens | 8.93 | 21.45 | 0.005 | 0.020 |
|  |  | OTU375 | [Paraprevotellaceae] | [Prevotella] | tannerae | 68.32 | 23.72 | 0.002 | 0.008 |
|  |  | OTU377 | Bacteroidaceae | Bacteroides | uniformis | 9.86 | 20.84 | 0.006 | 0.024 |
|  |  | OTU390 | Porphyromonadaceae | Parabacteroides | gordonii | 546.23 | 25.94 | 1.72x10^-12^ | 1.73x10^-10^ |
|  |  | OTU461 | Flavobacteriaceae | Flavobacterium | Unclassified | 6.77 | 17.84 | 0.001 | 0.004 |
|  |  | OTU529 | Pseudomonadaceae | Pseudomonas | aeruginosa | 28.26 | -29.04 | 9.94x10^-06^ | 1.37x10^-04^ |
|  |  | OTU540 | Enterobacteriaceae | Escherichia | coli | 18.38 | 13.54 | 0.004 | 0.018 |
|  |  | OTU574 | Enterobacteriaceae | Klebsiella | Unclassified | 1019.18 | 10.72 | 0.001 | 0.006 |
|  |  | OTU583 | Enterobacteriaceae | Plesiomonas | shigelloides | 8.12 | 18.57 | 0.001 | 0.004 |
|  |  | OTU593 | Comamonadaceae | Brachymonas | denitrificans | 4.44 | 30.00 | 8.46x10^-05^ | 0.001 |
|  |  | OTU594 | Comamonadaceae | Brachymonas | denitrificans | 4.34 | 22.36 | 0.003 | 0.014 |
|  |  | OTU603 | Comamonadaceae | Brachymonas | denitrificans | 1.68 | 21.30 | 0.005 | 0.020 |
|  |  | OTU606 | Comamonadaceae | Brachymonas | denitrificans | 13.62 | 23.37 | 3.71x10^-05^ | 4.31x10^-04^ |
|  |  | OTU618 | Comamonadaceae | Brachymonas | denitrificans | 21.97 | -16.20 | 1.72x10^-04^ | 0.001 |
|  |  | OTU739 | Veillonellaceae | Acidaminococcus | Unclassified | 90.79 | 21.65 | 2.10x10^-04^ | 0.001 |
|  |  | OTU772 | Streptococcaceae | Lactococcus | Unclassified | 48.94 | 14.43 | 0.006 | 0.022 |
|  |  | OTU780 | Lactobacillaceae | Lactobacillus | helveticus | 96.06 | 23.43 | 0.002 | 0.009 |
|  |  | OTU922 | Ruminococcaceae | Oscillospira | Unclassified | 1.99 | 22.54 | 0.003 | 0.013 |
|  |  | OTU930 | Ruminococcaceae | Oscillospira | Unclassified | 3.56 | 24.43 | 0.001 | 0.007 |
|  |  | OTU992 | Ruminococcaceae | Faecalibacterium | prausnitzii | 27.51 | -30.00 | 5.90x10^-06^ | 8.90x10^-05^ |
|  |  | OTU1121 | Lachnospiraceae | Coprococcus | Unclassified | 16.51 | -30.00 | 8.27x10^-05^ | 0.001 |
|  |  | OTU1152 | Fusobacteriaceae | Cetobacterium | somerae | 332.56 | 23.17 | 1.03x10^-04^ | 0.001 |
|  |  | OTU1158 | Lachnospiraceae | Epulopiscium | Unclassified | 223.82 | 25.07 | 2.26x10^-07^ | 4.55x10^-06^ |
|  |  | OTU1164 | Lachnospiraceae | Epulopiscium | Unclassified | 5.45 | -18.04 | 0.002 | 0.008 |
|  |  | OTU1176 | Lachnospiraceae | Epulopiscium | Unclassified | 79.09 | 22.26 | 1.45x10^-08^ | 5.46x10^-07^ |
|  |  | OTU1194 | Lachnospiraceae | Lachnospira | Unclassified | 5.91 | -30.00 | 1.69x10^-05^ | 2.12x10^-04^ |
|  |  | OTU1200 | Lachnospiraceae | Epulopiscium | Unclassified | 1705.47 | 26.80 | 4.97x10^-22^ | 1.50x10^-19^ |
|  |  | OTU1205 | Lachnospiraceae | Epulopiscium | Unclassified | 101.04 | 20.87 | 1.93x10^-07^ | 4.15x10^-06^ |
|  |  | OTU1218 | Lachnospiraceae | Epulopiscium | Unclassified | 2.51 | 20.29 | 0.008 | 0.030 |
|  |  | OTU1222 | Lachnospiraceae | Epulopiscium | Unclassified | 562.88 | 25.28 | 1.15x10^-16^ | 1.73x10^-14^ |
|  |  | OTU1229 | Lachnospiraceae | Epulopiscium | Unclassified | 5.88 | 15.88 | 0.010 | 0.036 |
|  |  | OTU1245 | Lachnospiraceae | Epulopiscium | Unclassified | 187.77 | 21.83 | 4.50x10^-07^ | 8.50x10^-06^ |
|  |  | OTU1250 | Lachnospiraceae | Epulopiscium | Unclassified | 45.28 | 20.63 | 6.35x10^-09^ | 3.12x10^-07^ |
|  |  | OTU1257 | Lachnospiraceae | Epulopiscium | Unclassified | 3.68 | -25.60 | 0.001 | 0.004 |
|  | **Inflamed** | OTU149 | Thermaceae | Thermus | Unclassified | 27.88 | 17.31 | 0.009 | 0.034 |
|  |  | OTU240 | Corynebacteriaceae | Corynebacterium | Unclassified | 3.43 | -21.75 | 0.001 | 0.005 |
|  |  | OTU323 | Verrucomicrobiaceae | Akkermansia | muciniphila | 103.75 | 22.40 | 1.03x10^-04^ | 0.001 |
|  |  | OTU358 | Prevotellaceae | Prevotella | nigrescens | 8.93 | 21.45 | 0.005 | 0.020 |
|  |  | OTU375 | [Paraprevotellaceae] | [Prevotella] | tannerae | 68.32 | 23.72 | 0.002 | 0.008 |
|  |  | OTU377 | Bacteroidaceae | Bacteroides | uniformis | 9.86 | 20.84 | 0.006 | 0.024 |
|  |  | OTU390 | Porphyromonadaceae | Parabacteroides | gordonii | 546.23 | 25.94 | 1.72x10^-12^ | 1.73x10^-10^ |
|  |  | OTU461 | Flavobacteriaceae | Flavobacterium | Unclassified | 6.77 | 17.84 | 0.001 | 0.004 |
|  |  | OTU529 | Pseudomonadaceae | Pseudomonas | aeruginosa | 28.26 | -29.04 | 9.94x10^-06^ | 1.37x10^-04^ |
|  |  | OTU540 | Enterobacteriaceae | Escherichia | coli | 18.38 | 13.54 | 0.004 | 0.018 |
|  |  | OTU574 | Enterobacteriaceae | Klebsiella | Unclassified | 1019.18 | 10.72 | 0.001 | 0.006 |
|  |  | OTU583 | Enterobacteriaceae | Plesiomonas | shigelloides | 8.12 | 18.57 | 0.001 | 0.004 |
|  |  | OTU593 | Comamonadaceae | Brachymonas | denitrificans | 4.44 | 30.00 | 8.46x10^-05^ | 0.001 |
|  |  | OTU594 | Comamonadaceae | Brachymonas | denitrificans | 4.34 | 22.36 | 0.003 | 0.014 |
|  |  | OTU603 | Comamonadaceae | Brachymonas | denitrificans | 1.68 | 21.30 | 0.005 | 0.020 |
|  |  | OTU606 | Comamonadaceae | Brachymonas | denitrificans | 13.62 | 23.37 | 3.71x10^-05^ | 4.31x10^-04^ |
|  |  | OTU618 | Comamonadaceae | Brachymonas | denitrificans | 21.97 | -16.20 | 1.72x10^-04^ | 0.001 |
|  |  | OTU739 | Veillonellaceae | Acidaminococcus | Unclassified | 90.79 | 21.65 | 2.10x10^-04^ | 0.001 |
|  |  | OTU772 | Streptococcaceae | Lactococcus | Unclassified | 48.94 | 14.43 | 0.006 | 0.022 |
|  |  | OTU780 | Lactobacillaceae | Lactobacillus | helveticus | 96.06 | 23.43 | 0.002 | 0.009 |
|  |  | OTU922 | Ruminococcaceae | Oscillospira | Unclassified | 1.99 | 22.54 | 0.003 | 0.013 |
|  |  | OTU930 | Ruminococcaceae | Oscillospira | Unclassified | 3.56 | 24.43 | 0.001 | 0.007 |
|  |  | OTU992 | Ruminococcaceae | Faecalibacterium | prausnitzii | 27.51 | -30.00 | 5.90x10^-06^ | 8.90x10^-05^ |
|  |  | OTU1121 | Lachnospiraceae | Coprococcus | Unclassified | 16.51 | -30.00 | 8.27x10^-05^ | 0.001 |
|  |  | OTU1152 | Fusobacteriaceae | Cetobacterium | somerae | 332.56 | 23.17 | 1.03x10^-04^ | 0.001 |
|  |  | OTU1158 | Lachnospiraceae | Epulopiscium | Unclassified | 223.82 | 25.07 | 2.26x10^-07^ | 4.55x10^-06^ |
|  |  | OTU1164 | Lachnospiraceae | Epulopiscium | Unclassified | 5.45 | -18.04 | 0.002 | 0.008 |
|  |  | OTU1176 | Lachnospiraceae | Epulopiscium | Unclassified | 79.09 | 22.26 | 1.45x10^-08^ | 5.46x10^-07^ |
|  |  | OTU1194 | Lachnospiraceae | Lachnospira | Unclassified | 5.91 | -30.00 | 1.69x10^-05^ | 2.12x10^-04^ |
|  |  | OTU1200 | Lachnospiraceae | Epulopiscium | Unclassified | 1705.47 | 26.80 | 4.97x10^-22^ | 1.50x10^-19^ |
|  |  | OTU1205 | Lachnospiraceae | Epulopiscium | Unclassified | 101.04 | 20.87 | 1.93x10^-07^ | 4.15x10^-06^ |
|  |  | OTU1218 | Lachnospiraceae | Epulopiscium | Unclassified | 2.51 | 20.29 | 0.008 | 0.030 |
|  |  | OTU1222 | Lachnospiraceae | Epulopiscium | Unclassified | 562.88 | 25.28 | 1.15x10^-16^ | 1.73x10^-14^ |
|  |  | OTU1229 | Lachnospiraceae | Epulopiscium | Unclassified | 5.88 | 15.88 | 0.010 | 0.036 |
|  |  | OTU1245 | Lachnospiraceae | Epulopiscium | Unclassified | 187.77 | 21.83 | 4.50x10^-07^ | 8.50x10^-06^ |
|  |  | OTU1250 | Lachnospiraceae | Epulopiscium | Unclassified | 45.28 | 20.63 | 6.35x10^-09^ | 3.12x10^-07^ |
|  |  | OTU1257 | Lachnospiraceae | Epulopiscium | Unclassified | 3.68 | -25.60 | 0.001 | 0.004 |
| **POR vs IBD-free controls** | **Uninflamed** | OTU149 | Thermaceae | Thermus | Unclassified | 27.88 | 19.76 | 0.001 | 0.005 |
|  |  | OTU323 | Verrucomicrobiaceae | Akkermansia | muciniphila | 103.75 | 20.46 | 8.65x10^-05^ | 0.001 |
|  |  | OTU358 | Prevotellaceae | Prevotella | nigrescens | 8.93 | 21.54 | 0.002 | 0.009 |
|  |  | OTU377 | Bacteroidaceae | Bacteroides | uniformis | 9.86 | 23.55 | 0.001 | 0.004 |
|  |  | OTU390 | Porphyromonadaceae | Parabacteroides | gordonii | 546.23 | 25.29 | 3.76x10^-14^ | 4.27x10^-12^ |
|  |  | OTU461 | Flavobacteriaceae | Flavobacterium | Unclassified | 6.77 | 18.92 | 9.53x10^-05^ | 0.001 |
|  |  | OTU516 | Xanthomonadaceae | Stenotrophomonas | maltophilia | 31.26 | -10.20 | 0.001 | 0.007 |
|  |  | OTU529 | Pseudomonadaceae | Pseudomonas | aeruginosa | 28.26 | 18.06 | 0.002 | 0.011 |
|  |  | OTU540 | Enterobacteriaceae | Escherichia | coli | 18.38 | 19.90 | 3.66x10^-06^ | 5.45x10^-05^ |
|  |  | OTU574 | Enterobacteriaceae | Klebsiella | Unclassified | 1019.18 | 11.27 | 1.66x10^-04^ | 0.001 |
|  |  | OTU583 | Enterobacteriaceae | Plesiomonas | shigelloides | 8.12 | 19.10 | 1.45x10^-04^ | 0.001 |
|  |  | OTU593 | Comamonadaceae | Brachymonas | denitrificans | 4.44 | 17.30 | 0.012 | 0.043 |
|  |  | OTU594 | Comamonadaceae | Brachymonas | denitrificans | 4.34 | 20.50 | 0.003 | 0.013 |
|  |  | OTU606 | Comamonadaceae | Brachymonas | denitrificans | 13.62 | 19.74 | 1.20x10^-04^ | 0.001 |
|  |  | OTU618 | Comamonadaceae | Brachymonas | denitrificans | 21.97 | -18.94 | 1.15x10^-06^ | 2.05x10^-05^ |
|  |  | OTU739 | Veillonellaceae | Acidaminococcus | Unclassified | 90.79 | 20.72 | 8.97x10^-05^ | 0.001 |
|  |  | OTU775 | Streptococcaceae | Streptococcus | Unclassified | 1862.03 | 3.73 | 0.002 | 0.010 |
|  |  | OTU780 | Lactobacillaceae | Lactobacillus | helveticus | 96.06 | 23.33 | 0.001 | 0.004 |
|  |  | OTU808 | Bacillaceae | Bacillus | cereus | 53.72 | 7.72 | 0.001 | 0.006 |
|  |  | OTU916 | Ruminococcaceae | Oscillospira | Unclassified | 10.65 | -19.34 | 0.005 | 0.020 |
|  |  | OTU922 | Ruminococcaceae | Oscillospira | Unclassified | 1.99 | 19.86 | 0.004 | 0.017 |
|  |  | OTU930 | Ruminococcaceae | Oscillospira | Unclassified | 3.56 | -29.80 | 1.62x10^-05^ | 1.57x10^-04^ |
|  |  | OTU992 | Ruminococcaceae | Faecalibacterium | prausnitzii | 27.51 | -15.60 | 0.009 | 0.033 |
|  |  | OTU1121 | Lachnospiraceae | Coprococcus | Unclassified | 16.51 | -30.00 | 1.29x10^-05^ | 1.37x10^-04^ |
|  |  | OTU1152 | Fusobacteriaceae | Cetobacterium | somerae | 332.56 | 21.59 | 6.40x10^-05^ | 0.001 |
|  |  | OTU1158 | Lachnospiraceae | Epulopiscium | Unclassified | 223.82 | 21.46 | 1.02x10^-06^ | 1.93x10^-05^ |
|  |  | OTU1164 | Lachnospiraceae | Epulopiscium | Unclassified | 5.45 | -14.44 | 0.005 | 0.021 |
|  |  | OTU1176 | Lachnospiraceae | Epulopiscium | Unclassified | 79.09 | 18.42 | 2.43x10^-07^ | 6.36x10^-06^ |
|  |  | OTU1194 | Lachnospiraceae | Lachnospira | Unclassified | 5.91 | -18.80 | 0.003 | 0.013 |
|  |  | OTU1200 | Lachnospiraceae | Epulopiscium | Unclassified | 1705.47 | 26.58 | 9.71x10^-26^ | 3.30x10^-23^ |
|  |  | OTU1204 | Lachnospiraceae | Epulopiscium | Unclassified | 3610.72 | 4.50 | 0.011 | 0.038 |
|  |  | OTU1205 | Lachnospiraceae | Epulopiscium | Unclassified | 101.04 | 17.16 | 2.48x10^-06^ | 4.01x10^-05^ |
|  |  | OTU1222 | Lachnospiraceae | Epulopiscium | Unclassified | 562.88 | 24.82 | 4.23x10^-19^ | 7.19x10^-17^ |
|  |  | OTU1229 | Lachnospiraceae | Epulopiscium | Unclassified | 5.88 | 19.09 | 0.001 | 0.004 |
|  |  | OTU1245 | Lachnospiraceae | Epulopiscium | Unclassified | 187.77 | 19.33 | 8.58x10^-07^ | 1.82x10^-05^ |
|  |  | OTU1250 | Lachnospiraceae | Epulopiscium | Unclassified | 45.28 | 18.55 | 9.46x10^-09^ | 3.22x10^-07^ |
|  | **Inflamed** | OTU149 | Thermaceae | Thermus | Unclassified | 27.88 | 20.76 | 0.001 | 0.003 |
|  |  | OTU323 | Verrucomicrobiaceae | Akkermansia | muciniphila | 103.75 | 23.33 | 6.87x10^-06^ | 1.04x10^-04^ |
|  |  | OTU358 | Prevotellaceae | Prevotella | nigrescens | 8.93 | 20.69 | 0.003 | 0.015 |
|  |  | OTU375 | [Paraprevotellaceae] | [Prevotella] | tannerae | 68.32 | 24.16 | 4.40x10^-04^ | 0.003 |
|  |  | OTU377 | Bacteroidaceae | Bacteroides | uniformis | 9.86 | -29.87 | 1.41x10^-05^ | 1.51x10^-04^ |
|  |  | OTU390 | Porphyromonadaceae | Parabacteroides | gordonii | 546.23 | 23.98 | 5.63x10^-13^ | 5.86x10^-11^ |
|  |  | OTU461 | Flavobacteriaceae | Flavobacterium | Unclassified | 6.77 | 21.87 | 5.79x10^-06^ | 9.27x10^-05^ |
|  |  | OTU514 | Xanthomonadaceae | Stenotrophomonas | maltophilia | 2.90 | 21.97 | 0.001 | 0.008 |
|  |  | OTU529 | Pseudomonadaceae | Pseudomonas | aeruginosa | 28.26 | 23.64 | 6.22x10^-05^ | 0.001 |
|  |  | OTU540 | Enterobacteriaceae | Escherichia | coli | 18.38 | 20.25 | 2.21x10^-06^ | 4.39x10^-05^ |
|  |  | OTU574 | Enterobacteriaceae | Klebsiella | Unclassified | 1019.18 | 10.08 | 0.001 | 0.005 |
|  |  | OTU583 | Enterobacteriaceae | Plesiomonas | shigelloides | 8.12 | 18.96 | 1.52x10^-04^ | 0.001 |
|  |  | OTU593 | Comamonadaceae | Brachymonas | denitrificans | 4.44 | 18.62 | 0.007 | 0.032 |
|  |  | OTU594 | Comamonadaceae | Brachymonas | denitrificans | 4.34 | 18.77 | 0.006 | 0.031 |
|  |  | OTU606 | Comamonadaceae | Brachymonas | denitrificans | 13.62 | 20.59 | 5.54x10^-05^ | 0.001 |
|  |  | OTU618 | Comamonadaceae | Brachymonas | denitrificans | 21.97 | -15.73 | 4.92x10^-05^ | 4.66x10^-04^ |
|  |  | OTU739 | Veillonellaceae | Acidaminococcus | Unclassified | 90.79 | 23.24 | 1.01x10^-05^ | 1.36x10^-04^ |
|  |  | OTU780 | Lactobacillaceae | Lactobacillus | helveticus | 96.06 | -17.89 | 0.009 | 0.043 |
|  |  | OTU808 | Bacillaceae | Bacillus | cereus | 53.72 | 8.19 | 0.001 | 0.003 |
|  |  | OTU916 | Ruminococcaceae | Oscillospira | Unclassified | 10.65 | -20.61 | 0.003 | 0.014 |
|  |  | OTU922 | Ruminococcaceae | Oscillospira | Unclassified | 1.99 | 21.05 | 0.002 | 0.013 |
|  |  | OTU930 | Ruminococcaceae | Oscillospira | Unclassified | 3.56 | 20.80 | 0.002 | 0.014 |
|  |  | OTU1121 | Lachnospiraceae | Coprococcus | Unclassified | 16.51 | -30.00 | 1.18x10^-05^ | 1.36x10^-04^ |
|  |  | OTU1152 | Fusobacteriaceae | Cetobacterium | somerae | 332.56 | 21.30 | 7.41x10^-05^ | 0.001 |
|  |  | OTU1158 | Lachnospiraceae | Epulopiscium | Unclassified | 223.82 | 20.71 | 2.16x10^-06^ | 4.39x10^-05^ |
|  |  | OTU1164 | Lachnospiraceae | Epulopiscium | Unclassified | 5.45 | -16.85 | 0.001 | 0.007 |
|  |  | OTU1176 | Lachnospiraceae | Epulopiscium | Unclassified | 79.09 | 18.25 | 2.76x10^-07^ | 8.82x10^-06^ |
|  |  | OTU1179 | Lachnospiraceae | Lachnospira | Unclassified | 11.74 | -30.00 | 3.04x10^-06^ | 5.49x10^-05^ |
|  |  | OTU1188 | Lachnospiraceae | Epulopiscium | Unclassified | 2.83 | -29.96 | 1.21x10^-05^ | 1.36x10^-04^ |
|  |  | OTU1194 | Lachnospiraceae | Lachnospira | Unclassified | 5.91 | -17.85 | 0.004 | 0.023 |
|  |  | OTU1200 | Lachnospiraceae | Epulopiscium | Unclassified | 1705.47 | 26.08 | 4.97x10^-25^ | 2.07x10^-22^ |
|  |  | OTU1205 | Lachnospiraceae | Epulopiscium | Unclassified | 101.04 | 16.77 | 3.78x10^-06^ | 6.29x10^-05^ |
|  |  | OTU1222 | Lachnospiraceae | Epulopiscium | Unclassified | 562.88 | 24.61 | 5.85x10^-19^ | 1.22x10^-16^ |
|  |  | OTU1229 | Lachnospiraceae | Epulopiscium | Unclassified | 5.88 | 22.63 | 4.31x10^-05^ | 4.27x10^-04^ |
|  |  | OTU1245 | Lachnospiraceae | Epulopiscium | Unclassified | 187.77 | 19.43 | 6.67x10^-07^ | 1.63x10^-05^ |
|  |  | OTU1250 | Lachnospiraceae | Epulopiscium | Unclassified | 45.28 | 18.70 | 6.14x10^-09^ | 2.84x10^-07^ |
|  |  | OTU1257 | Lachnospiraceae | Epulopiscium | Unclassified | 3.68 | -23.87 | 0.001 | 0.003 |
| **Mild POR vs IBD-free controls** | **Uninflamed** | OTU128 | Nitrososphaeraceae | Candidatus Nitrososphaera | gargensis | 2.10 | -24.60 | 0.001 | 0.003 |
|  |  | OTU149 | Thermaceae | Thermus | Unclassified | 27.88 | 18.81 | 0.004 | 0.015 |
|  |  | OTU249 | Mycobacteriaceae | Mycobacterium | Unclassified | 3.29 | 18.55 | 0.003 | 0.012 |
|  |  | OTU323 | Verrucomicrobiaceae | Akkermansia | muciniphila | 109.17 | 20.30 | 1.91x10^-04^ | 0.001 |
|  |  | OTU347 | [Paraprevotellaceae] | Paraprevotella | Unclassified | 54.57 | 17.62 | 0.013 | 0.044 |
|  |  | OTU390 | Porphyromonadaceae | Parabacteroides | gordonii | 546.23 | 24.31 | 2.40x10^-12^ | 2.51x10^-10^ |
|  |  | OTU461 | Flavobacteriaceae | Flavobacterium | Unclassified | 8.31 | 19.04 | 1.73x10^-04^ | 0.001 |
|  |  | OTU516 | Xanthomonadaceae | Stenotrophomonas | maltophilia | 31.26 | -11.28 | 0.001 | 0.003 |
|  |  | OTU540 | Enterobacteriaceae | Escherichia | coli | 18.38 | 19.61 | 1.74x10^-05^ | 2.27x10^-04^ |
|  |  | OTU574 | Enterobacteriaceae | Klebsiella | Unclassified | 1019.18 | 10.00 | 0.001 | 0.006 |
|  |  | OTU592 | Comamonadaceae | Comamonas | Unclassified | 9.63 | 17.81 | 1.46x10^-06^ | 3.28x10^-05^ |
|  |  | OTU594 | Comamonadaceae | Brachymonas | denitrificans | 4.34 | 18.21 | 0.010 | 0.036 |
|  |  | OTU603 | Comamonadaceae | Brachymonas | denitrificans | 1.68 | -18.31 | 0.010 | 0.036 |
|  |  | OTU606 | Comamonadaceae | Brachymonas | denitrificans | 14.66 | 17.64 | 0.001 | 0.005 |
|  |  | OTU618 | Comamonadaceae | Brachymonas | denitrificans | 21.97 | -17.23 | 2.37x10^-05^ | 2.27x10^-04^ |
|  |  | OTU739 | Veillonellaceae | Acidaminococcus | Unclassified | 81.16 | -13.67 | 0.015 | 0.049 |
|  |  | OTU775 | Streptococcaceae | Streptococcus | Unclassified | 1862.03 | 4.03 | 0.001 | 0.006 |
|  |  | OTU808 | Bacillaceae | Bacillus | cereus | 53.72 | 7.35 | 0.002 | 0.010 |
|  |  | OTU916 | Ruminococcaceae | Oscillospira | Unclassified | 10.65 | -29.10 | 3.82x10^-05^ | 3.43x10^-04^ |
|  |  | OTU922 | Ruminococcaceae | Oscillospira | Unclassified | 1.99 | 21.43 | 0.003 | 0.011 |
|  |  | OTU930 | Ruminococcaceae | Oscillospira | Unclassified | 3.56 | -22.30 | 0.002 | 0.008 |
|  |  | OTU1121 | Lachnospiraceae | Coprococcus | Unclassified | 16.51 | -30.00 | 2.28x10^-05^ | 2.27x10^-04^ |
|  |  | OTU1152 | Fusobacteriaceae | Cetobacterium | somerae | 332.56 | 20.63 | 9.35x10^-05^ | 0.001 |
|  |  | OTU1161 | Lachnospiraceae | Epulopiscium | Unclassified | 229.86 | 24.52 | 1.16x10^-05^ | 1.66x10^-04^ |
|  |  | OTU1164 | Lachnospiraceae | Epulopiscium | Unclassified | 5.45 | -14.90 | 0.008 | 0.029 |
|  |  | OTU1176 | Lachnospiraceae | Epulopiscium | Unclassified | 79.09 | 18.22 | 6.54x10^-07^ | 1.71x10^-05^ |
|  |  | OTU1188 | Lachnospiraceae | Epulopiscium | Unclassified | 2.83 | -23.01 | 0.001 | 0.006 |
|  |  | OTU1194 | Lachnospiraceae | Lachnospira | Unclassified | 5.91 | -19.67 | 0.005 | 0.021 |
|  |  | OTU1200 | Lachnospiraceae | Epulopiscium | Unclassified | 1705.47 | 22.76 | 9.99x10^-20^ | 3.14x10^-17^ |
|  |  | OTU1205 | Lachnospiraceae | Epulopiscium | Unclassified | 101.04 | 16.20 | 9.10x10^-06^ | 1.39x10^-04^ |
|  |  | OTU1222 | Lachnospiraceae | Epulopiscium | Unclassified | 562.88 | 24.52 | 1.91x10^-17^ | 2.99x10^-15^ |
|  |  | OTU1229 | Lachnospiraceae | Epulopiscium | Unclassified | 5.88 | 19.60 | 0.001 | 0.005 |
|  |  | OTU1245 | Lachnospiraceae | Epulopiscium | Unclassified | 187.77 | 19.98 | 1.01x10^-06^ | 2.44x10^-05^ |
|  |  | OTU1250 | Lachnospiraceae | Epulopiscium | Unclassified | 45.28 | 18.15 | 5.49x10^-08^ | 1.91x10^-06^ |
|  | **Inflamed** | OTU128 | Nitrososphaeraceae | Candidatus Nitrososphaera | gargensis | 2.10 | -25.01 | 3.83x10^-04^ | 0.002 |
|  |  | OTU149 | Thermaceae | Thermus | Unclassified | 27.88 | 21.52 | 0.001 | 0.004 |
|  |  | OTU249 | Mycobacteriaceae | Mycobacterium | Unclassified | 3.29 | 19.83 | 0.001 | 0.006 |
|  |  | OTU323 | Verrucomicrobiaceae | Akkermansia | muciniphila | 109.17 | 21.12 | 9.78x10^-05^ | 0.001 |
|  |  | OTU390 | Porphyromonadaceae | Parabacteroides | gordonii | 546.23 | 23.81 | 5.67x10^-12^ | 4.20x10^-10^ |
|  |  | OTU461 | Flavobacteriaceae | Flavobacterium | Unclassified | 8.31 | 19.76 | 9.19x10^-05^ | 0.001 |
|  |  | OTU514 | Xanthomonadaceae | Stenotrophomonas | maltophilia | 6.88 | 23.87 | 0.001 | 0.004 |
|  |  | OTU540 | Enterobacteriaceae | Escherichia | coli | 18.38 | 20.48 | 6.77x10^-06^ | 1.05x10^-04^ |
|  |  | OTU574 | Enterobacteriaceae | Klebsiella | Unclassified | 1019.18 | 8.50 | 0.006 | 0.021 |
|  |  | OTU592 | Comamonadaceae | Comamonas | Unclassified | 9.63 | 19.35 | 1.48x10^-07^ | 3.99x10^-06^ |
|  |  | OTU594 | Comamonadaceae | Brachymonas | denitrificans | 4.34 | 18.54 | 0.009 | 0.032 |
|  |  | OTU603 | Comamonadaceae | Brachymonas | denitrificans | 1.68 | -19.74 | 0.005 | 0.021 |
|  |  | OTU606 | Comamonadaceae | Brachymonas | denitrificans | 14.66 | 21.06 | 7.04x10^-05^ | 0.001 |
|  |  | OTU618 | Comamonadaceae | Brachymonas | denitrificans | 21.97 | -16.82 | 3.44x10^-05^ | 2.75x10^-04^ |
|  |  | OTU739 | Veillonellaceae | Acidaminococcus | Unclassified | 81.16 | 22.28 | 7.15x10^-05^ | 0.001 |
|  |  | OTU808 | Bacillaceae | Bacillus | cereus | 53.72 | 7.85 | 0.001 | 0.005 |
|  |  | OTU916 | Ruminococcaceae | Oscillospira | Unclassified | 10.65 | -30.00 | 2.04x10^-05^ | 1.85x10^-04^ |
|  |  | OTU922 | Ruminococcaceae | Oscillospira | Unclassified | 1.99 | 22.58 | 0.001 | 0.007 |
|  |  | OTU930 | Ruminococcaceae | Oscillospira | Unclassified | 3.56 | 24.27 | 0.001 | 0.003 |
|  |  | OTU1121 | Lachnospiraceae | Coprococcus | Unclassified | 16.51 | -30.00 | 2.13x10^-05^ | 1.85x10^-04^ |
|  |  | OTU1157 | Lachnospiraceae | Epulopiscium | Unclassified | 11.93 | -20.01 | 0.004 | 0.017 |
|  |  | OTU1161 | Lachnospiraceae | Epulopiscium | Unclassified | 229.86 | 24.26 | 1.35x10^-05^ | 1.54x10^-04^ |
|  |  | OTU1164 | Lachnospiraceae | Epulopiscium | Unclassified | 5.45 | -19.86 | 3.89x10^-04^ | 0.002 |
|  |  | OTU1176 | Lachnospiraceae | Epulopiscium | Unclassified | 79.09 | 18.11 | 7.00x10^-07^ | 1.38x10^-05^ |
|  |  | OTU1188 | Lachnospiraceae | Epulopiscium | Unclassified | 2.83 | -23.54 | 0.001 | 0.004 |
|  |  | OTU1194 | Lachnospiraceae | Lachnospira | Unclassified | 5.91 | -17.21 | 0.014 | 0.049 |
|  |  | OTU1200 | Lachnospiraceae | Epulopiscium | Unclassified | 1705.47 | 22.94 | 3.87x10^-20^ | 1.15x10^-17^ |
|  |  | OTU1205 | Lachnospiraceae | Epulopiscium | Unclassified | 101.04 | 16.08 | 9.83x10^-06^ | 1.21x10^-04^ |
|  |  | OTU1222 | Lachnospiraceae | Epulopiscium | Unclassified | 562.88 | 23.85 | 1.09x10^-16^ | 1.62x10^-14^ |
|  |  | OTU1229 | Lachnospiraceae | Epulopiscium | Unclassified | 5.88 | 23.94 | 4.17x10^-05^ | 3.25x10^-04^ |
|  |  | OTU1245 | Lachnospiraceae | Epulopiscium | Unclassified | 187.77 | 20.90 | 2.77x10^-07^ | 6.30x10^-06^ |
|  |  | OTU1250 | Lachnospiraceae | Epulopiscium | Unclassified | 45.28 | 18.46 | 2.89x10^-08^ | 9.51x10^-07^ |
| **Severe POR vs IBD-free controls** | **Uninflamed** | OTU128 | Nitrososphaeraceae | Candidatus Nitrososphaera | gargensis | 2.10 | -21.94 | 0.005 | 0.021 |
|  |  | OTU149 | Thermaceae | Thermus | Unclassified | 27.88 | 18.07 | 0.012 | 0.042 |
|  |  | OTU240 | Corynebacteriaceae | Corynebacterium | Unclassified | 3.43 | -18.19 | 0.011 | 0.042 |
|  |  | OTU249 | Mycobacteriaceae | Mycobacterium | Unclassified | 3.29 | -21.81 | 0.002 | 0.008 |
|  |  | OTU323 | Verrucomicrobiaceae | Akkermansia | muciniphila | 109.17 | -18.81 | 0.002 | 0.009 |
|  |  | OTU347 | [Paraprevotellaceae] | Paraprevotella | Unclassified | 54.57 | 29.83 | 1.54x10^-04^ | 0.001 |
|  |  | OTU390 | Porphyromonadaceae | Parabacteroides | gordonii | 546.23 | 26.72 | 3.08x10^-12^ | 3.34x10^-10^ |
|  |  | OTU461 | Flavobacteriaceae | Flavobacterium | Unclassified | 8.31 | 21.51 | 1.30x10^-04^ | 0.001 |
|  |  | OTU516 | Xanthomonadaceae | Stenotrophomonas | maltophilia | 31.26 | -10.97 | 0.003 | 0.011 |
|  |  | OTU540 | Enterobacteriaceae | Escherichia | coli | 18.38 | 18.91 | 1.86x10^-04^ | 0.001 |
|  |  | OTU574 | Enterobacteriaceae | Klebsiella | Unclassified | 1019.18 | 12.42 | 2.69x10^-04^ | 0.002 |
|  |  | OTU592 | Comamonadaceae | Comamonas | Unclassified | 9.63 | 16.86 | 3.85x10^-05^ | 4.83x10^-04^ |
|  |  | OTU593 | Comamonadaceae | Brachymonas | denitrificans | 4.44 | 19.31 | 0.014 | 0.048 |
|  |  | OTU594 | Comamonadaceae | Brachymonas | denitrificans | 4.34 | 24.91 | 0.002 | 0.008 |
|  |  | OTU606 | Comamonadaceae | Brachymonas | denitrificans | 14.66 | 23.96 | 4.83x10^-05^ | 0.001 |
|  |  | OTU618 | Comamonadaceae | Brachymonas | denitrificans | 21.97 | -30.00 | 4.29x10^-11^ | 2.33x10^-09^ |
|  |  | OTU739 | Veillonellaceae | Acidaminococcus | Unclassified | 81.16 | 22.42 | 3.29x10^-04^ | 0.002 |
|  |  | OTU922 | Ruminococcaceae | Oscillospira | Unclassified | 1.99 | -27.05 | 0.001 | 0.003 |
|  |  | OTU992 | Ruminococcaceae | Faecalibacterium | prausnitzii | 27.51 | -30.00 | 2.14x10^-05^ | 3.31x10^-04^ |
|  |  | OTU1121 | Lachnospiraceae | Coprococcus | Unclassified | 16.51 | -30.00 | 1.39x10^-04^ | 0.001 |
|  |  | OTU1152 | Fusobacteriaceae | Cetobacterium | somerae | 332.56 | 24.80 | 2.26x10^-05^ | 3.35x10^-04^ |
|  |  | OTU1164 | Lachnospiraceae | Epulopiscium | Unclassified | 5.45 | -19.83 | 0.002 | 0.008 |
|  |  | OTU1176 | Lachnospiraceae | Epulopiscium | Unclassified | 79.09 | 15.00 | 0.000 | 0.001 |
|  |  | OTU1194 | Lachnospiraceae | Lachnospira | Unclassified | 5.91 | -19.43 | 0.013 | 0.046 |
|  |  | OTU1196 | Lachnospiraceae | Lachnospira | Unclassified | 2.68 | -30.00 | 1.43x10^-04^ | 0.001 |
|  |  | OTU1200 | Lachnospiraceae | Epulopiscium | Unclassified | 1705.47 | 27.89 | 3.87x10^-24^ | 1.26x10^-21^ |
|  |  | OTU1204 | Lachnospiraceae | Epulopiscium | Unclassified | 3610.72 | 5.00 | 0.014 | 0.046 |
|  |  | OTU1205 | Lachnospiraceae | Epulopiscium | Unclassified | 101.04 | -27.72 | 9.48x10^-12^ | 7.72x10^-10^ |
|  |  | OTU1222 | Lachnospiraceae | Epulopiscium | Unclassified | 562.88 | 24.06 | 3.83x10^-14^ | 6.24x10^-12^ |
|  |  | OTU1229 | Lachnospiraceae | Epulopiscium | Unclassified | 5.88 | -29.82 | 4.93x10^-06^ | 8.93x10^-05^ |
|  |  | OTU1245 | Lachnospiraceae | Epulopiscium | Unclassified | 187.77 | 15.62 | 0.001 | 0.003 |
|  |  | OTU1250 | Lachnospiraceae | Epulopiscium | Unclassified | 45.28 | 15.33 | 3.56x10^-05^ | 4.83x10^-04^ |
|  | **Inflamed** | OTU128 | Nitrososphaeraceae | Candidatus Nitrososphaera | gargensis | 2.10 | -22.20 | 0.004 | 0.016 |
|  |  | OTU240 | Corynebacteriaceae | Corynebacterium | Unclassified | 3.43 | -19.11 | 0.007 | 0.023 |
|  |  | OTU249 | Mycobacteriaceae | Mycobacterium | Unclassified | 3.29 | 18.45 | 0.006 | 0.022 |
|  |  | OTU323 | Verrucomicrobiaceae | Akkermansia | muciniphila | 109.17 | 22.29 | 1.74x10^-04^ | 0.001 |
|  |  | OTU347 | [Paraprevotellaceae] | Paraprevotella | Unclassified | 54.57 | 30.00 | 1.08x10^-04^ | 0.001 |
|  |  | OTU390 | Porphyromonadaceae | Parabacteroides | gordonii | 546.23 | 24.59 | 6.92x10^-11^ | 5.12x10^-09^ |
|  |  | OTU461 | Flavobacteriaceae | Flavobacterium | Unclassified | 8.31 | 20.04 | 2.88x10^-04^ | 0.002 |
|  |  | OTU514 | Xanthomonadaceae | Stenotrophomonas | maltophilia | 6.88 | 21.90 | 0.005 | 0.018 |
|  |  | OTU516 | Xanthomonadaceae | Stenotrophomonas | maltophilia | 31.26 | -13.54 | 1.52x10^-04^ | 0.001 |
|  |  | OTU540 | Enterobacteriaceae | Escherichia | coli | 18.38 | 17.29 | 0.001 | 0.003 |
|  |  | OTU574 | Enterobacteriaceae | Klebsiella | Unclassified | 1019.18 | 11.26 | 0.001 | 0.004 |
|  |  | OTU592 | Comamonadaceae | Comamonas | Unclassified | 9.63 | 18.76 | 3.05x10^-06^ | 6.46x10^-05^ |
|  |  | OTU594 | Comamonadaceae | Brachymonas | denitrificans | 4.34 | -29.82 | 1.23x10^-04^ | 0.001 |
|  |  | OTU603 | Comamonadaceae | Brachymonas | denitrificans | 1.68 | 27.40 | 4.13x10^-04^ | 0.002 |
|  |  | OTU606 | Comamonadaceae | Brachymonas | denitrificans | 14.66 | 22.08 | 1.40x10^-04^ | 0.001 |
|  |  | OTU618 | Comamonadaceae | Brachymonas | denitrificans | 21.97 | -14.13 | 0.002 | 0.007 |
|  |  | OTU739 | Veillonellaceae | Acidaminococcus | Unclassified | 81.16 | 24.42 | 6.91x10^-05^ | 0.001 |
|  |  | OTU745 | Veillonellaceae | Dialister | Unclassified | 79.68 | 10.27 | 0.010 | 0.032 |
|  |  | OTU922 | Ruminococcaceae | Oscillospira | Unclassified | 1.99 | -29.53 | 1.44x10^-04^ | 0.001 |
|  |  | OTU989 | Ruminococcaceae | Faecalibacterium | prausnitzii | 819.41 | -7.83 | 0.009 | 0.030 |
|  |  | OTU992 | Ruminococcaceae | Faecalibacterium | prausnitzii | 27.51 | -30.00 | 1.52x10^-05^ | 2.51x10^-04^ |
|  |  | OTU1121 | Lachnospiraceae | Coprococcus | Unclassified | 16.51 | -30.00 | 1.06x10^-04^ | 0.001 |
|  |  | OTU1152 | Fusobacteriaceae | Cetobacterium | somerae | 332.56 | 27.39 | 1.93x10^-06^ | 4.38x10^-05^ |
|  |  | OTU1161 | Lachnospiraceae | Epulopiscium | Unclassified | 229.86 | -14.97 | 0.015 | 0.042 |
|  |  | OTU1164 | Lachnospiraceae | Epulopiscium | Unclassified | 5.45 | -18.49 | 0.003 | 0.010 |
|  |  | OTU1176 | Lachnospiraceae | Epulopiscium | Unclassified | 79.09 | 14.01 | 4.66x10^-04^ | 0.003 |
|  |  | OTU1179 | Lachnospiraceae | Lachnospira | Unclassified | 13.06 | -29.99 | 2.67x10^-05^ | 3.96x10^-04^ |
|  |  | OTU1188 | Lachnospiraceae | Epulopiscium | Unclassified | 2.83 | -21.72 | 0.005 | 0.018 |
|  |  | OTU1194 | Lachnospiraceae | Lachnospira | Unclassified | 5.91 | -20.45 | 0.008 | 0.027 |
|  |  | OTU1196 | Lachnospiraceae | Lachnospira | Unclassified | 2.68 | -30.00 | 1.10x10^-04^ | 0.001 |
|  |  | OTU1200 | Lachnospiraceae | Epulopiscium | Unclassified | 1705.47 | 27.05 | 1.81x10^-23^ | 5.37x10^-21^ |
|  |  | OTU1201 | Lachnospiraceae | Epulopiscium | Unclassified | 16.30 | -13.43 | 0.008 | 0.026 |
|  |  | OTU1205 | Lachnospiraceae | Epulopiscium | Unclassified | 101.04 | 9.46 | 0.018 | 0.049 |
|  |  | OTU1222 | Lachnospiraceae | Epulopiscium | Unclassified | 562.88 | 24.72 | 2.82x10^-15^ | 4.17x10^-13^ |
|  |  | OTU1229 | Lachnospiraceae | Epulopiscium | Unclassified | 5.88 | 16.43 | 0.010 | 0.032 |
|  |  | OTU1245 | Lachnospiraceae | Epulopiscium | Unclassified | 187.77 | 15.64 | 4.56x10^-04^ | 0.002 |
|  |  | OTU1250 | Lachnospiraceae | Epulopiscium | Unclassified | 45.28 | 15.91 | 1.25x10^-05^ | 2.18x10^-04^ |

IBD, inflammatory bowel diseases; OTU, operational taxonomic unit; POR, post-operative recurrence.

***Table S10.*** *DEseq2 results: differentially expressed OTUs between POR in both mucosal zones.*

|  |  | **OTU** | **Family** | **Genus** | **Species** | **baseMean** | **log_2_FC** | **P-value** | **P-adj** |
| --- | --- | --- | --- | --- | --- | --- | --- | --- | --- |
| **POR vs Non-POR** | **Uninflamed** | OTU240 | Corynebacteriaceae | Corynebacterium | Unclassified | 3.43 | 19.62 | 1.39x10^-06^ | 1.69x10^-05^ |
|  |  | OTU329 | Bacteroidaceae | Bacteroides | uniformis | 163.89 | -11.60 | 1.41x10^-04^ | 0.001 |
|  |  | OTU356 | Prevotellaceae | Prevotella | nigrescens | 0.84 | -23.26 | 6.51x10^-07^ | 9.00x10^-06^ |
|  |  | OTU364 | Prevotellaceae | Prevotella | Unclassified | 3.92 | -23.65 | 4.22x10^-07^ | 7.14x10^-06^ |
|  |  | OTU384 | Porphyromonadaceae | Parabacteroides | gordonii | 1.76 | -23.92 | 3.14x10^-07^ | 5.73x10^-06^ |
|  |  | OTU392 | Porphyromonadaceae | Porphyromonas | Unclassified | 0.89 | -23.23 | 6.74x10^-07^ | 9.00x10^-06^ |
|  |  | OTU393 | Porphyromonadaceae | Paludibacter | Unclassified | 0.18 | -21.26 | 5.46x10^-06^ | 4.80x10^-05^ |
|  |  | OTU459 | Flavobacteriaceae | Capnocytophaga | Unclassified | 1.02 | -23.75 | 3.78x10^-07^ | 6.64x10^-06^ |
|  |  | OTU460 | Flavobacteriaceae | Capnocytophaga | ochracea | 0.52 | -22.54 | 1.43x10^-06^ | 1.69x10^-05^ |
|  |  | OTU473 | [Odoribacteraceae] | Odoribacter | Unclassified | 0.26 | -21.66 | 3.61x10^-06^ | 3.53x10^-05^ |
|  |  | OTU529 | Pseudomonadaceae | Pseudomonas | aeruginosa | 28.26 | 17.36 | 1.56x10^-05^ | 1.32x10^-04^ |
|  |  | OTU603 | Comamonadaceae | Brachymonas | denitrificans | 1.68 | -29.61 | 1.97x10^-10^ | 1.33x10^-08^ |
|  |  | OTU623 | Pasteurellaceae | Gallibacterium | Unclassified | 846.24 | 5.35 | 0.004 | 0.026 |
|  |  | OTU772 | Streptococcaceae | Lactococcus | Unclassified | 48.94 | -9.46 | 0.003 | 0.021 |
|  |  | OTU868 | Spirochaetaceae | Treponema | socranskii | 0.48 | -22.48 | 1.54x10^-06^ | 1.73x10^-05^ |
|  |  | OTU930 | Ruminococcaceae | Oscillospira | Unclassified | 3.56 | -29.83 | 1.60x10^-10^ | 1.26x10^-08^ |
|  |  | OTU989 | Ruminococcaceae | Faecalibacterium | prausnitzii | 819.41 | 5.22 | 0.005 | 0.034 |
|  |  | OTU992 | Ruminococcaceae | Faecalibacterium | prausnitzii | 27.51 | 14.40 | 4.12x10^-04^ | 0.003 |
|  |  | OTU1121 | Lachnospiraceae | Coprococcus | Unclassified | 16.51 | -15.54 | 0.001 | 0.007 |
|  |  | OTU1147 | Leptotrichiaceae | Leptotrichia | Unclassified | 1.02 | -23.60 | 4.47x10^-07^ | 7.30x10^-06^ |
|  |  | OTU1148 | Leptotrichiaceae | Leptotrichia | Unclassified | 0.40 | 16.47 | 4.32x10^-04^ | 0.003 |
|  |  | OTU1218 | Lachnospiraceae | Epulopiscium | Unclassified | 2.51 | -13.16 | 0.005 | 0.032 |
|  |  | OTU1257 | Lachnospiraceae | Epulopiscium | Unclassified | 3.68 | 21.04 | 6.72x10^-06^ | 5.79x10^-05^ |
|  | **Inflamed** | OTU246 | Corynebacteriaceae | Corynebacterium | durum | 1.92 | 18.08 | 7.00x10^-07^ | 1.54x10^-05^ |
|  |  | OTU329 | Bacteroidaceae | Bacteroides | uniformis | 163.89 | -10.04 | 0.001 | 0.009 |
|  |  | OTU364 | Prevotellaceae | Prevotella | Unclassified | 3.92 | -12.97 | 0.005 | 0.041 |
|  |  | OTU377 | Bacteroidaceae | Bacteroides | uniformis | 9.86 | -25.29 | 4.49x10^-08^ | 1.34x10^-06^ |
|  |  | OTU387 | Porphyromonadaceae | Parabacteroides | gordonii | 10.99 | 24.40 | 1.38x10^-07^ | 3.68x10^-06^ |
|  |  | OTU434 | Cytophagaceae | Dyadobacter | Unclassified | 0.39 | 20.55 | 9.11x10^-06^ | 1.18x10^-04^ |
|  |  | OTU514 | Xanthomonadaceae | Stenotrophomonas | maltophilia | 2.90 | 20.75 | 7.30x10^-06^ | 9.99x10^-05^ |
|  |  | OTU603 | Comamonadaceae | Brachymonas | denitrificans | 1.68 | 17.47 | 1.64x10^-04^ | 0.002 |
|  |  | OTU623 | Pasteurellaceae | Gallibacterium | Unclassified | 846.24 | 5.47 | 0.003 | 0.026 |
|  |  | OTU772 | Streptococcaceae | Lactococcus | Unclassified | 48.94 | 23.33 | 3.32x10^-13^ | 8.39x10^-11^ |
|  |  | OTU780 | Lactobacillaceae | Lactobacillus | helveticus | 96.06 | -28.75 | 4.89x10^-10^ | 2.25x10^-08^ |
|  |  | OTU908 | Clostridiaceae | 02d06 | Unclassified | 6.56 | -13.39 | 0.001 | 0.009 |
|  |  | OTU916 | Ruminococcaceae | Oscillospira | Unclassified | 10.65 | -20.86 | 5.88x10^-06^ | 8.26x10^-05^ |
|  |  | OTU1121 | Lachnospiraceae | Coprococcus | Unclassified | 16.51 | -16.91 | 2.59x10^-04^ | 0.003 |
|  |  | OTU1148 | Leptotrichiaceae | Leptotrichia | Unclassified | 0.40 | -24.70 | 8.44x10^-08^ | 2.37x10^-06^ |
|  |  | OTU1149 | Leptotrichiaceae | Leptotrichia | Unclassified | 3.71 | 21.27 | 4.32x10^-06^ | 6.34x10^-05^ |
|  |  | OTU1179 | Lachnospiraceae | Lachnospira | Unclassified | 11.74 | -29.31 | 1.38x10^-11^ | 1.40x10^-09^ |
|  |  | OTU1188 | Lachnospiraceae | Epulopiscium | Unclassified | 2.83 | -25.64 | 2.78x10^-08^ | 9.38x10^-07^ |
|  |  | OTU1194 | Lachnospiraceae | Lachnospira | Unclassified | 5.91 | 20.17 | 2.08x10^-06^ | 3.51x10^-05^ |
|  |  | OTU1205 | Lachnospiraceae | Epulopiscium | Unclassified | 101.04 | -6.89 | 0.004 | 0.032 |
|  |  | OTU1223 | Lachnospiraceae | Epulopiscium | Unclassified | 12.72 | 22.38 | 3.59x10^-12^ | 5.10x10^-10^ |
| **Mild POR vs Non-POR** | **Uninflamed** | OTU240 | Corynebacteriaceae | Corynebacterium | Unclassified | 3.43 | 19.39 | 2.82x10^-05^ | 2.84x10^-04^ |
|  |  | OTU329 | Bacteroidaceae | Bacteroides | uniformis | 163.89 | -10.27 | 0.002 | 0.016 |
|  |  | OTU347 | [Paraprevotellaceae] | Paraprevotella | Unclassified | 54.57 | 17.57 | 0.001 | 0.004 |
|  |  | OTU356 | Prevotellaceae | Prevotella | nigrescens | 0.84 | -22.98 | 5.72x10^-06^ | 9.15x10^-05^ |
|  |  | OTU364 | Prevotellaceae | Prevotella | Unclassified | 3.92 | -23.99 | 2.17x10^-06^ | 4.22x10^-05^ |
|  |  | OTU384 | Porphyromonadaceae | Parabacteroides | gordonii | 1.76 | -24.26 | 1.67x10^-06^ | 3.63x10^-05^ |
|  |  | OTU393 | Porphyromonadaceae | Paludibacter | Unclassified | 0.18 | -20.95 | 3.55x10^-05^ | 3.25x10^-04^ |
|  |  | OTU460 | Flavobacteriaceae | Capnocytophaga | ochracea | 0.52 | -22.33 | 1.04x10^-05^ | 1.25x10^-04^ |
|  |  | OTU473 | [Odoribacteraceae] | Odoribacter | Unclassified | 0.26 | -21.39 | 2.41x10^-05^ | 2.52x10^-04^ |
|  |  | OTU603 | Comamonadaceae | Brachymonas | denitrificans | 1.68 | -24.46 | 1.33x10^-06^ | 3.16x10^-05^ |
|  |  | OTU676 | Rhodobacteraceae | Paracoccus | marcusii | 0.45 | 16.31 | 0.001 | 0.009 |
|  |  | OTU738 | Veillonellaceae | Acidaminococcus | Unclassified | 15.09 | 16.22 | 0.001 | 0.009 |
|  |  | OTU739 | Veillonellaceae | Acidaminococcus | Unclassified | 81.16 | -25.02 | 3.63x10^-10^ | 9.88x10^-08^ |
|  |  | OTU868 | Spirochaetaceae | Treponema | socranskii | 0.48 | -22.66 | 7.72x10^-06^ | 1.10x10^-04^ |
|  |  | OTU869 | Spirochaetaceae | Treponema | socranskii | 0.45 | -22.08 | 1.31x10^-05^ | 1.55x10^-04^ |
|  |  | OTU916 | Ruminococcaceae | Oscillospira | Unclassified | 10.65 | -19.90 | 8.37x10^-05^ | 0.001 |
|  |  | OTU930 | Ruminococcaceae | Oscillospira | Unclassified | 3.56 | -25.33 | 5.50x10^-07^ | 1.76x10^-05^ |
|  |  | OTU989 | Ruminococcaceae | Faecalibacterium | prausnitzii | 819.41 | 6.92 | 4.44x10^-04^ | 0.003 |
|  |  | OTU992 | Ruminococcaceae | Faecalibacterium | prausnitzii | 27.51 | 16.24 | 3.57x10^-04^ | 0.003 |
|  |  | OTU1082 | Peptostreptococcaceae | Filifactor | Unclassified | 0.29 | -21.29 | 2.58x10^-05^ | 2.65x10^-04^ |
|  |  | OTU1188 | Lachnospiraceae | Epulopiscium | Unclassified | 2.83 | -22.79 | 6.63x10^-06^ | 1.00x10^-04^ |
|  |  | OTU1196 | Lachnospiraceae | Lachnospira | Unclassified | 2.68 | 15.13 | 0.003 | 0.019 |
|  |  | OTU1238 | Lachnospiraceae | Epulopiscium | Unclassified | 21.46 | -22.00 | 1.36x10^-05^ | 1.58x10^-04^ |
|  | **Inflamed** | OTU246 | Corynebacteriaceae | Corynebacterium | durum | 1.92 | 17.18 | 3.41x10^-05^ | 2.99x10^-04^ |
|  |  | OTU341 | Bacteroidaceae | Bacteroides | uniformis | 26.24 | 26.28 | 1.34x10^-07^ | 2.84x10^-06^ |
|  |  | OTU347 | [Paraprevotellaceae] | Paraprevotella | Unclassified | 54.57 | 23.41 | 2.60x10^-06^ | 3.46x10^-05^ |
|  |  | OTU514 | Xanthomonadaceae | Stenotrophomonas | maltophilia | 6.88 | 29.12 | 4.95x10^-09^ | 2.11x10^-07^ |
|  |  | OTU623 | Pasteurellaceae | Gallibacterium | Unclassified | 846.24 | 5.40 | 0.006 | 0.035 |
|  |  | OTU676 | Rhodobacteraceae | Paracoccus | marcusii | 0.45 | -17.04 | 0.001 | 0.004 |
|  |  | OTU738 | Veillonellaceae | Acidaminococcus | Unclassified | 15.09 | -12.93 | 0.009 | 0.048 |
|  |  | OTU772 | Streptococcaceae | Lactococcus | Unclassified | 48.94 | 22.73 | 1.94x10^-10^ | 2.32x10^-08^ |
|  |  | OTU908 | Clostridiaceae | 02d06 | Unclassified | 6.56 | -15.65 | 0.001 | 0.007 |
|  |  | OTU916 | Ruminococcaceae | Oscillospira | Unclassified | 10.65 | -28.66 | 7.74x10^-09^ | 2.78x10^-07^ |
|  |  | OTU1121 | Lachnospiraceae | Coprococcus | Unclassified | 16.51 | -16.34 | 0.001 | 0.007 |
|  |  | OTU1152 | Fusobacteriaceae | Cetobacterium | somerae | 332.56 | -18.23 | 7.25x10^-07^ | 1.13x10^-05^ |
|  |  | OTU1188 | Lachnospiraceae | Epulopiscium | Unclassified | 2.83 | -20.25 | 4.73x10^-05^ | 3.95x10^-04^ |
|  |  | OTU1194 | Lachnospiraceae | Lachnospira | Unclassified | 5.91 | 25.93 | 1.94x10^-07^ | 3.67x10^-06^ |
|  |  | OTU1196 | Lachnospiraceae | Lachnospira | Unclassified | 2.68 | 20.88 | 2.80x10^-05^ | 2.51x10^-04^ |
|  |  | OTU1223 | Lachnospiraceae | Epulopiscium | Unclassified | 12.72 | 22.92 | 2.68x10^-10^ | 2.40x10^-08^ |
| **Severe POR vs Non-POR** | **Uninflamed** | OTU240 | Corynebacteriaceae | Corynebacterium | Unclassified | 3.43 | -16.66 | 0.002 | 0.013 |
|  |  | OTU323 | Verrucomicrobiaceae | Akkermansia | muciniphila | 109.17 | -27.23 | 1.27x10^-09^ | 1.73x10^-07^ |
|  |  | OTU329 | Bacteroidaceae | Bacteroides | uniformis | 163.89 | -12.36 | 0.002 | 0.011 |
|  |  | OTU341 | Bacteroidaceae | Bacteroides | uniformis | 26.24 | -21.09 | 3.32x10^-04^ | 0.003 |
|  |  | OTU347 | [Paraprevotellaceae] | Paraprevotella | Unclassified | 54.57 | 29.83 | 3.52x10^-07^ | 1.74x10^-05^ |
|  |  | OTU356 | Prevotellaceae | Prevotella | nigrescens | 0.84 | -21.17 | 3.21x10^-04^ | 0.003 |
|  |  | OTU364 | Prevotellaceae | Prevotella | Unclassified | 3.92 | -22.20 | 1.61x10^-04^ | 0.002 |
|  |  | OTU381 | Porphyromonadaceae | Parabacteroides | Unclassified | 6.01 | -23.82 | 5.13x10^-05^ | 0.001 |
|  |  | OTU384 | Porphyromonadaceae | Parabacteroides | gordonii | 1.76 | -21.78 | 2.14x10^-04^ | 0.002 |
|  |  | OTU385 | Porphyromonadaceae | Parabacteroides | gordonii | 5.13 | 22.28 | 1.49x10^-04^ | 0.002 |
|  |  | OTU393 | Porphyromonadaceae | Paludibacter | Unclassified | 0.18 | -19.21 | 0.001 | 0.008 |
|  |  | OTU460 | Flavobacteriaceae | Capnocytophaga | ochracea | 0.52 | -20.53 | 4.82x10^-04^ | 0.004 |
|  |  | OTU473 | [Odoribacteraceae] | Odoribacter | Unclassified | 0.26 | -19.72 | 0.001 | 0.006 |
|  |  | OTU582 | Enterobacteriaceae | Escherichia | coli | 4856.97 | 2.85 | 0.006 | 0.036 |
|  |  | OTU603 | Comamonadaceae | Brachymonas | denitrificans | 1.68 | -21.36 | 2.80x10^-04^ | 0.003 |
|  |  | OTU618 | Comamonadaceae | Brachymonas | denitrificans | 21.97 | -22.33 | 6.61x10^-11^ | 1.20x10^-08^ |
|  |  | OTU623 | Pasteurellaceae | Gallibacterium | Unclassified | 846.24 | 6.69 | 0.004 | 0.024 |
|  |  | OTU868 | Spirochaetaceae | Treponema | socranskii | 0.48 | -19.70 | 0.001 | 0.006 |
|  |  | OTU869 | Spirochaetaceae | Treponema | socranskii | 0.45 | -20.39 | 0.001 | 0.005 |
|  |  | OTU922 | Ruminococcaceae | Oscillospira | Unclassified | 1.99 | -27.97 | 1.93x10^-06^ | 6.56x10^-05^ |
|  |  | OTU930 | Ruminococcaceae | Oscillospira | Unclassified | 3.56 | -22.37 | 1.42x10^-04^ | 0.002 |
|  |  | OTU1147 | Leptotrichiaceae | Leptotrichia | Unclassified | 1.82 | -21.53 | 2.53x10^-04^ | 0.003 |
|  |  | OTU1161 | Lachnospiraceae | Epulopiscium | Unclassified | 229.86 | -22.63 | 9.36x10^-07^ | 3.64x10^-05^ |
|  |  | OTU1205 | Lachnospiraceae | Epulopiscium | Unclassified | 101.04 | -30.00 | 1.10x10-23 | 5.96x10-21 |
|  |  | OTU1218 | Lachnospiraceae | Epulopiscium | Unclassified | 2.51 | -30.00 | 3.35x10^-07^ | 1.74x10^-05^ |
|  |  | OTU1229 | Lachnospiraceae | Epulopiscium | Unclassified | 5.88 | -15.07 | 0.002 | 0.013 |
|  |  | OTU1260 | Lachnospiraceae | Epulopiscium | Unclassified | 0.45 | -17.47 | 0.003 | 0.019 |
|  | **Inflamed** | OTU226 | Bifidobacteriaceae | Bifidobacterium | longum | 433.14 | -5.62 | 0.005 | 0.033 |
|  |  | OTU240 | Corynebacteriaceae | Corynebacterium | Unclassified | 3.43 | -21.72 | 3.67x10^-05^ | 4.57x10^-04^ |
|  |  | OTU246 | Corynebacteriaceae | Corynebacterium | durum | 1.92 | 18.59 | 1.07x10^-04^ | 0.001 |
|  |  | OTU329 | Bacteroidaceae | Bacteroides | uniformis | 163.89 | -11.80 | 0.002 | 0.016 |
|  |  | OTU336 | Bacteroidaceae | Bacteroides | uniformis | 553.37 | -14.64 | 7.80x10^-07^ | 2.03x10^-05^ |
|  |  | OTU341 | Bacteroidaceae | Bacteroides | uniformis | 26.24 | 15.58 | 0.007 | 0.045 |
|  |  | OTU347 | [Paraprevotellaceae] | Paraprevotella | Unclassified | 54.57 | 30.00 | 1.81x10^-07^ | 7.82x10^-06^ |
|  |  | OTU385 | Porphyromonadaceae | Parabacteroides | gordonii | 5.13 | 23.41 | 4.94x10^-05^ | 0.001 |
|  |  | OTU582 | Enterobacteriaceae | Escherichia | coli | 4856.97 | 2.96 | 0.004 | 0.026 |
|  |  | OTU594 | Comamonadaceae | Brachymonas | denitrificans | 4.34 | -23.29 | 5.25x10^-05^ | 0.001 |
|  |  | OTU603 | Comamonadaceae | Brachymonas | denitrificans | 1.68 | 29.87 | 2.24x10^-07^ | 7.94x10^-06^ |
|  |  | OTU623 | Pasteurellaceae | Gallibacterium | Unclassified | 846.24 | 7.20 | 0.002 | 0.012 |
|  |  | OTU714 | Rhodospirillaceae | Azospirillum | amazonense | 2.70 | 22.09 | 1.29x10^-04^ | 0.001 |
|  |  | OTU738 | Veillonellaceae | Acidaminococcus | Unclassified | 15.09 | -30.00 | 1.95x10^-07^ | 7.82x10^-06^ |
|  |  | OTU744 | Veillonellaceae | Megasphaera | Unclassified | 7.22 | 27.31 | 2.12x10^-06^ | 4.59x10^-05^ |
|  |  | OTU745 | Veillonellaceae | Dialister | Unclassified | 79.68 | 9.59 | 0.001 | 0.010 |
|  |  | OTU772 | Streptococcaceae | Lactococcus | Unclassified | 48.94 | 25.54 | 6.44x10^-10^ | 1.25x10^-07^ |
|  |  | OTU922 | Ruminococcaceae | Oscillospira | Unclassified | 1.99 | -26.41 | 4.59x10^-06^ | 8.53x10^-05^ |
|  |  | OTU930 | Ruminococcaceae | Oscillospira | Unclassified | 3.56 | -25.11 | 1.33x10^-05^ | 2.07x10^-04^ |
|  |  | OTU992 | Ruminococcaceae | Faecalibacterium | prausnitzii | 27.51 | -19.06 | 2.31x10^-04^ | 0.002 |
|  |  | OTU1121 | Lachnospiraceae | Coprococcus | Unclassified | 16.51 | -15.84 | 0.006 | 0.040 |
|  |  | OTU1161 | Lachnospiraceae | Epulopiscium | Unclassified | 229.86 | -23.08 | 3.44x10^-07^ | 1.12x10^-05^ |
|  |  | OTU1179 | Lachnospiraceae | Lachnospira | Unclassified | 13.06 | -25.57 | 1.55x10^-06^ | 3.56x10^-05^ |
|  |  | OTU1188 | Lachnospiraceae | Epulopiscium | Unclassified | 2.83 | -20.21 | 4.59x10^-04^ | 0.004 |
|  |  | OTU1194 | Lachnospiraceae | Lachnospira | Unclassified | 5.91 | 19.43 | 0.001 | 0.006 |
|  |  | OTU1205 | Lachnospiraceae | Epulopiscium | Unclassified | 101.04 | -12.32 | 2.25x10^-05^ | 3.14x10^-04^ |
|  |  | OTU1223 | Lachnospiraceae | Epulopiscium | Unclassified | 12.72 | 29.99 | 7.29x10^-13^ | 2.84x10^-10^ |
|  |  | OTU1260 | Lachnospiraceae | Epulopiscium | Unclassified | 0.45 | -25.04 | 1.39x10^-05^ | 2.08x10^-04^ |
| **Severe POR vs Mild POR** | **Uninflamed** | OTU231 | Actinomycetaceae | Arcanobacterium | Unclassified | 9.90 | -23.78 | 1.63x10^-10^ | 2.58x10^-08^ |
|  |  | OTU240 | Corynebacteriaceae | Corynebacterium | Unclassified | 3.43 | -16.49 | 0.001 | 0.008 |
|  |  | OTU323 | Verrucomicrobiaceae | Akkermansia | muciniphila | 109.17 | -18.40 | 1.29x10^-05^ | 2.64x10^-04^ |
|  |  | OTU336 | Bacteroidaceae | Bacteroides | uniformis | 553.37 | -10.67 | 1.60x10^-04^ | 0.002 |
|  |  | OTU341 | Bacteroidaceae | Bacteroides | uniformis | 26.24 | -21.78 | 7.78x10^-05^ | 0.001 |
|  |  | OTU364 | Prevotellaceae | Prevotella | Unclassified | 3.92 | 15.11 | 0.006 | 0.036 |
|  |  | OTU385 | Porphyromonadaceae | Parabacteroides | gordonii | 5.13 | 23.42 | 2.11x10^-05^ | 3.93x10^-04^ |
|  |  | OTU514 | Xanthomonadaceae | Stenotrophomonas | maltophilia | 6.88 | -16.29 | 0.003 | 0.021 |
|  |  | OTU582 | Enterobacteriaceae | Escherichia | coli | 4856.97 | 3.21 | 0.001 | 0.008 |
|  |  | OTU603 | Comamonadaceae | Brachymonas | denitrificans | 1.68 | 18.21 | 0.001 | 0.008 |
|  |  | OTU618 | Comamonadaceae | Brachymonas | denitrificans | 21.97 | -12.75 | 7.39x10^-05^ | 0.001 |
|  |  | OTU738 | Veillonellaceae | Acidaminococcus | Unclassified | 15.09 | -16.09 | 0.004 | 0.023 |
|  |  | OTU739 | Veillonellaceae | Acidaminococcus | Unclassified | 81.16 | 25.22 | 6.20x10^-09^ | 4.90x10^-07^ |
|  |  | OTU745 | Veillonellaceae | Dialister | Unclassified | 79.68 | 7.94 | 0.005 | 0.032 |
|  |  | OTU916 | Ruminococcaceae | Oscillospira | Unclassified | 10.65 | 23.97 | 1.30x10^-05^ | 2.64x10^-04^ |
|  |  | OTU922 | Ruminococcaceae | Oscillospira | Unclassified | 1.99 | -18.31 | 0.001 | 0.008 |
|  |  | OTU930 | Ruminococcaceae | Oscillospira | Unclassified | 3.56 | 19.02 | 0.001 | 0.006 |
|  |  | OTU992 | Ruminococcaceae | Faecalibacterium | prausnitzii | 27.51 | -15.88 | 0.001 | 0.010 |
|  |  | OTU1161 | Lachnospiraceae | Epulopiscium | Unclassified | 229.86 | -24.51 | 1.49x10^-08^ | 9.43x10^-07^ |
|  |  | OTU1188 | Lachnospiraceae | Epulopiscium | Unclassified | 2.83 | 29.11 | 1.23x10^-07^ | 4.86x10^-06^ |
|  |  | OTU1200 | Lachnospiraceae | Epulopiscium | Unclassified | 1705.47 | 5.14 | 0.005 | 0.031 |
|  |  | OTU1205 | Lachnospiraceae | Epulopiscium | Unclassified | 101.04 | -24.26 | 7.37x10^-18^ | 2.33x10^-15^ |
|  |  | OTU1229 | Lachnospiraceae | Epulopiscium | Unclassified | 5.88 | -18.89 | 3.23x10^-05^ | 0.001 |
|  |  | OTU1238 | Lachnospiraceae | Epulopiscium | Unclassified | 21.46 | 24.49 | 8.67x10^-06^ | 2.28x10^-04^ |
|  | **Inflamed** | OTU240 | Corynebacteriaceae | Corynebacterium | Unclassified | 3.43 | -17.96 | 2.09x10^-04^ | 0.002 |
|  |  | OTU336 | Bacteroidaceae | Bacteroides | uniformis | 553.37 | -11.81 | 1.50x10^-05^ | 2.40x10^-04^ |
|  |  | OTU341 | Bacteroidaceae | Bacteroides | uniformis | 26.24 | -25.29 | 1.81x10^-06^ | 4.59x10^-05^ |
|  |  | OTU385 | Porphyromonadaceae | Parabacteroides | gordonii | 5.13 | 22.35 | 2.40x10^-05^ | 3.38x10^-04^ |
|  |  | OTU516 | Xanthomonadaceae | Stenotrophomonas | maltophilia | 31.26 | -7.70 | 0.002 | 0.013 |
|  |  | OTU582 | Enterobacteriaceae | Escherichia | coli | 4856.97 | 3.44 | 2.42x10^-04^ | 0.002 |
|  |  | OTU594 | Comamonadaceae | Brachymonas | denitrificans | 4.34 | -27.81 | 1.48x10^-07^ | 6.05x10^-06^ |
|  |  | OTU603 | Comamonadaceae | Brachymonas | denitrificans | 1.68 | 27.16 | 2.79x10^-07^ | 8.96x10^-06^ |
|  |  | OTU676 | Rhodobacteraceae | Paracoccus | marcusii | 0.45 | 28.15 | 9.64x10^-08^ | 4.82x10^-06^ |
|  |  | OTU738 | Veillonellaceae | Acidaminococcus | Unclassified | 15.09 | -23.54 | 8.80x10^-06^ | 1.58x10^-04^ |
|  |  | OTU745 | Veillonellaceae | Dialister | Unclassified | 79.68 | 13.09 | 1.83x10^-06^ | 4.59x10^-05^ |
|  |  | OTU916 | Ruminococcaceae | Oscillospira | Unclassified | 10.65 | 20.36 | 1.17x10^-04^ | 0.001 |
|  |  | OTU922 | Ruminococcaceae | Oscillospira | Unclassified | 1.99 | -29.14 | 3.62x10^-08^ | 2.72x10^-06^ |
|  |  | OTU930 | Ruminococcaceae | Oscillospira | Unclassified | 3.56 | -27.63 | 1.75x10^-07^ | 6.05x10^-06^ |
|  |  | OTU989 | Ruminococcaceae | Faecalibacterium | prausnitzii | 819.41 | -7.72 | 1.79x10^-04^ | 0.002 |
|  |  | OTU992 | Ruminococcaceae | Faecalibacterium | prausnitzii | 27.51 | -24.88 | 1.72x10^-07^ | 6.05x10^-06^ |
|  |  | OTU1081 | Clostridiaceae | SMB53 | Unclassified | 8.71 | 8.75 | 0.002 | 0.013 |
|  |  | OTU1152 | Fusobacteriaceae | Cetobacterium | somerae | 332.56 | 17.05 | 1.26x10^-05^ | 2.11x10^-04^ |
|  |  | OTU1157 | Lachnospiraceae | Epulopiscium | Unclassified | 11.93 | 14.16 | 0.007 | 0.045 |
|  |  | OTU1161 | Lachnospiraceae | Epulopiscium | Unclassified | 229.86 | -26.36 | 2.32x10^-10^ | 4.17x10^-08^ |
|  |  | OTU1179 | Lachnospiraceae | Lachnospira | Unclassified | 13.06 | -18.57 | 1.50x10^-04^ | 0.001 |
|  |  | OTU1196 | Lachnospiraceae | Lachnospira | Unclassified | 2.68 | -21.28 | 5.88x10^-05^ | 0.001 |
|  |  | OTU1260 | Lachnospiraceae | Epulopiscium | Unclassified | 0.45 | -18.97 | 3.44x10^-04^ | 0.003 |

IBD, inflammatory bowel diseases; OTU, operational taxonomic unit; POR, post-operative recurrence.

***Table S11.*** *Selected transcript-bacterial group pairs.*

|  |  |  |  |  | **Significant (NC)** | **ORA (NC)** |
| --- | --- | --- | --- | --- | --- | --- |
| **POR** | **Uninflamed** | - | *-* | - | - | - |
|  | **Inflamed** | GE57493 | *ITGAM* | Xanthomonadaceae | yes | yes |
|  |  | GE82051 | *LSR* | Porphyromonadaceae | yes | yes |
|  |  | GE79161 | *TFF1* |  | yes | yes |
| **Non-POR** | **Uninflamed** | - | *-* | - | - | - |
|  | **Inflamed** | - | *-* | - | - | - |

GEO, gene expression omnibus; ME; co-expression module; NC, non-coincident; OTU, operational taxonomic unit; POR, post-operative recurrence.
